# Neuroimaging-AI endophenotypes reveal underlying mechanisms and genetic factors contributing to progression and development of four brain disorders

**DOI:** 10.1101/2023.08.16.23294179

**Authors:** Junhao Wen, Ioanna Skampardoni, Ye Ella Tian, Zhijian Yang, Yuhan Cui, Guray Erus, Gyujoon Hwang, Erdem Varol, Aleix Boquet-Pujadas, Ganesh B. Chand, Ilya Nasrallah, Theodore D. Satterthwaite, Haochang Shou, Li Shen, Arthur W. Toga, Andrew Zalesky, Christos Davatzikos

## Abstract

Recent work leveraging artificial intelligence has offered promise to dissect disease heterogeneity by identifying complex intermediate brain phenotypes, called dimensional neuroimaging endophenotypes (DNEs). We advance the argument that these DNEs capture the degree of expression of respective neuroanatomical patterns measured, offering a dimensional neuroanatomical representation for studying disease heterogeneity and similarities of neurologic and neuropsychiatric diseases. We investigate the presence of nine DNEs derived from independent yet harmonized studies on Alzheimer’s disease, autism spectrum disorder, late-life depression, and schizophrenia in the UK Biobank study. Phenome-wide associations align with genome-wide associations, revealing 31 genomic loci (P-value<5×10^−8^/9) associated with the nine DNEs.The nine DNEs, along with their polygenic risk scores, significantly enhanced the predictive accuracy for 14 systemic disease categories, particularly for conditions related to mental health and the central nervous system, as well as mortality outcomes. These findings underscore the potential of the nine DNEs to capture the expression of disease-related brain phenotypes in individuals of the general population and to relate such measures with genetics, lifestyle factors, and chronic diseases.

## Main

Disease heterogeneity^1–8^ has been a significant challenge for precision medicine^9^. A new era powered by artificial intelligence (AI) and large-scale, multi-omics biomarkers may enable us to quantify individualized liability for various brain diseases^10,11^. Recent work has leveraged semi-supervised clustering methods (**Fig. 1a** and **Supplementary Method 1**) to tackle this challenge. These methods characterize disease heterogeneity by constructing a mapping or transformation from a reference group, or healthy controls, to a target group, such as patients with a specific disease. In clinical neuroscience, these methods can quantify deviation from typical brain structure measured by T1-weighted magnetic resonance imaging (MRI)^12,2,1,13^. They represent disease-related neuroanatomical heterogeneity with multiple low-dimensional categorical subtypes associated with specific patterns of brain change relative to the reference group. Instead of focusing on the categorical subtypes, we investigated their corresponding continuous phenotypes (i.e., dimensions)^14^, given that many chronic brain diseases develop along a continuous spectrum. Each neuroanatomical pattern’s level of expression, therefore, serves as a dimensional AI-derived biomarker pertinent to the respective disease.

**Figure 1:**
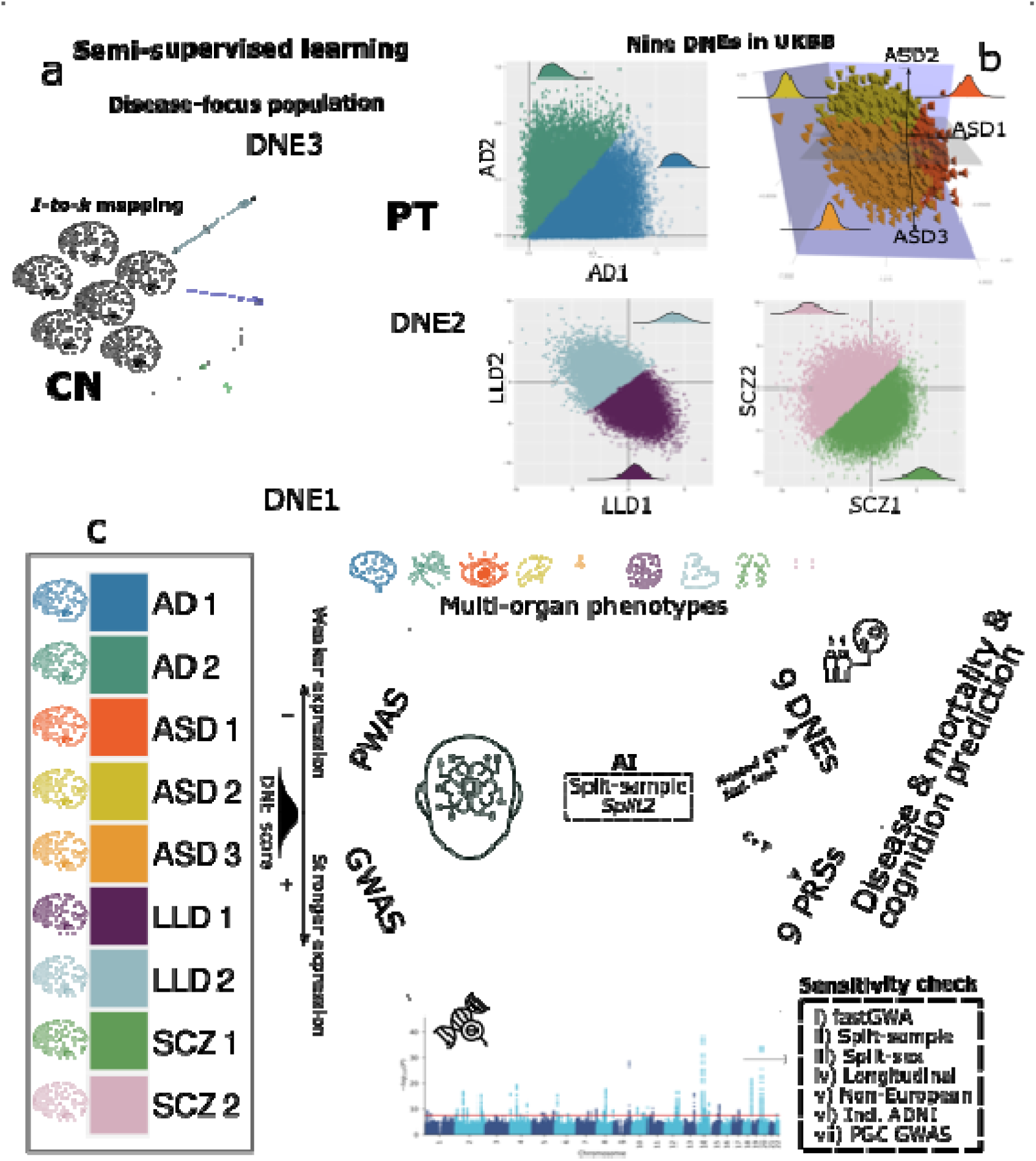
Study workflow. **a**) The concept of semi-supervised learning methods used in this study. These AI methods model the patterns and transformations from the healthy control (CN) to the patient (PT) domain, thus capturing variations related to underlying disease pathology. Nine DNEs previously published^12,2,1,13^ from four disease-focused, case-control studies were investigated. **b**) The expression of the nine DNEs in the UK Biobank (UKBB) general population. The trained models were then applied to the UKBB population to quantify the expression of the neuroanatomical patterns of the nine DNEs at individual levels; a higher DNE score indicates a greater expression (manifestation/presence) of the respective neuroanatomical pattern. For example, the blue samples express predominantly AD2, whereas the pink samples express predominantly SCZ2. The kernel density estimate for each DNE is shown. Of note, AD1-2 DNEs are from the Surreal-GAN^14^ model and others from the HYDRA^87^ model, resulting in varying DNE score ranges by modeling. Overall, lower scores imply milder imaging pattern expressions. **c**) Phenome- and genome-wide analyses were performed on the nine DENs. Phenome-wide association studies (PWAS) were conducted to associate the nine DNEs with phenotypes across nine organ systems, cognition, and lifestyle factors. Genome-wide association studies (GWAS) were performed to investigate associations between the nine DNEs and common genetic variants (SNPs). Finally, the nine DNEs and their polygenic risk scores predicted 14 disease categories (ICD-10-based), 8 cognitive scores, and mortality. CN: healthy control; PT: patient.

Previous heterogeneity research has primarily focused on within-disease heterogeneity^1–8^. However, this approach neglects the shared disease mechanisms, genetics, and clinical manifestations among different brain diseases. Conversely, while several studies have investigated the shared genetic components across various brain diseases, they have overlooked the important aspect of disease heterogeneity within each condition^15,16^. As such, a broad perspective is required to investigate disease heterogeneity simultaneously, spanning multiple neurodegenerative and neuropsychiatric disorders. This holistic approach aids in understanding the commonalities and interrelationships between these brain diseases and the multi-organ systems of humans^17,18^. Such an effort could simultaneously capture neurobiological heterogeneity within disorders and explain shared features, mechanisms, and risk factors across disorders. Ultimately, unraveling neurobiological heterogeneity within neuropsychiatric syndromes and explaining co-morbidity among them promises to accelerate more effective diagnosis, treatment, and prevention strategies.

These AI-derived biomarkers capture disease-specific neuroanatomic heterogeneity. However, whether these biomarkers are present in the general population, potentially simultaneously, remains unknown. Here, we sought to measure the presence of multiple AI-based signatures in the general population, delineate common mechanisms among them, and shed light on their relationship with other human organ systems^14,19,20^. To do this, we capitalized on nine imaging biomarkers recently derived from regional gray matter (GM) volumetrics derived from several large-scale disease-focused consortia, including ADNI^21^ for Alzheimer’s disease (AD1-2^12^), ABIDE^22^ for autism spectrum disorder (ASD1-3^1^), LLD for late-life depression older than 60 years old (LLD1-2^2^), and PHENOM^23^ for schizophrenia (SCZ1-2^23^). Using semi-supervised clustering and representation learning methodologies^24^, we obtained nine imaging biomarkers capturing the neuroanatomical diversity across the four distinct brain diseases. AD1 and AD2 illustrate distinct atrophy patterns in the overall brain and medial temporal lobe, respectively^12^. ASD1, ASD2, and ASD3 showcase reduced overall volume, expanded subcortical volume, and increased cortical volume^1^. LLD1 and LLD2^2^ exhibit augmented subcortical volume and overall brain atrophy, respectively. Finally, SCZ1 and SCZ2^23^ display global brain atrophy and expanded basal ganglia volume. We first conceptualized these biomarkers as the dimensional neuroimaging endophenotypes (DNE), seeking to test the endophenotype hypothesis in psychiatry^25–27^, which suggests that such measurable intermediate biomarkers (i.e., the endophenotypes) bridge genetics and clinical symptoms and the onset of the disease. They are thought to be more closely related to the underlying etiology and genetics than the complex clinical symptoms or the disease itself. Furthermore, the concept of dimensional representation reinforces the idea that diseases like AD progress along a continuous spectrum. As such, categorizing disease heterogeneity into distinct subtypes overlooks the presence of co-existing patterns within the same individuals (refer to **Supplementary Method 1a** for details).

We evaluated the manifestation of the nine DNEs in the general population using the neuroimaging and genetic data available in the UK Biobank study^28^ (UKBB). The four pre-trained disease-specific AI models were applied to the 39,178 UKBB participants with both brain MRI^29^ and genetic^30^ data. To delineate the phenotypic landscape of the nine DNEs, we first tested whether the neuroanatomical patterns of the nine DNEs are present in the UKBB general population. Subsequently, we conducted a phenome-wide association study (PWAS) to establish associations between the nine DNEs and 611 UKBB phenotypes, including brain imaging-derived phenotypes (IDPs), traits related to multiple human organ systems, cognition, and lifestyle factors. We performed a genome-wide association study (GWAS) linking the nine DNEs to 6,477,810 quality-checked common single nucleotide polymorphisms (SNPs) to depict their genetic architecture. Furthermore, we conducted analyses to investigate genetic correlations, colocalization, and causal relationships between the nine DNEs, nine human organ systems^17^, and several chronic diseases. Finally, we assessed the ability of the nine DNEs and their corresponding polygenic risk score (PRS) to predict 14 systemic disease categories, 8 cognitive scores, and mortality.

## Results

Our analytic framework involves computational genomics, statistical methods, and machine learning to elucidate the phenotypic landscape and genetic architecture of the nine DNEs, as illustrated in **Fig. 1**.

### All nine DNEs are evident in the general population

We tested whether the neuroanatomical patterns defined in the four disease populations could be found in the general population. We applied the DNE models pre-trained for each disease population to the UKBB general population to measure the degree of expression of each DNE at the individual level. The imaging data from the four disease populations and the UKBB general population were first statistically harmonized via the iSTAGING consortium^31^ and then linearly corrected in the AI models for common confounders like demographics to alleviate potential domain shifts^32^ (**Supplementary Text 1**).

We first summarize the neuroanatomical patterns of the nine DNEs (**Fig. 2a**). Overall, the original patterns identified in the disease populations^12,2,1,13^ manifest in the general population. AD1 exhibits a pattern of brain atrophy (i.e., negative correlation) across various brain volumes, while AD2 involves focal atrophy of the medial temporal lobe. ASD1 captures a pattern of lower GM volumes in several subcortical regions, including the pallidum, amygdala, and putamen, whereas ASD2 reflects a pattern of relatively larger GM volumes (i.e., positive correlation) in subcortical regions. ASD3, conversely, is characterized by relatively larger GM volumes in several cortical areas, including the insula. LLD1 (positive correlation) and LLD2 (negative correlation) are characterized by concomitant patterns of regional GM volumes, prominently involving the middle frontal gyrus, the insula, and the thalamus. For schizophrenia, a widespread pattern of reduced brain volumes (e.g., insula) is associated with SCZ1, whereas SCZ2 displays increased volumes of the putamen and pallidum. The details of the P-value, sample sizes, and β values of the linear regression are presented in **Supplementary File 1** and **2**.

**Figure 2:**
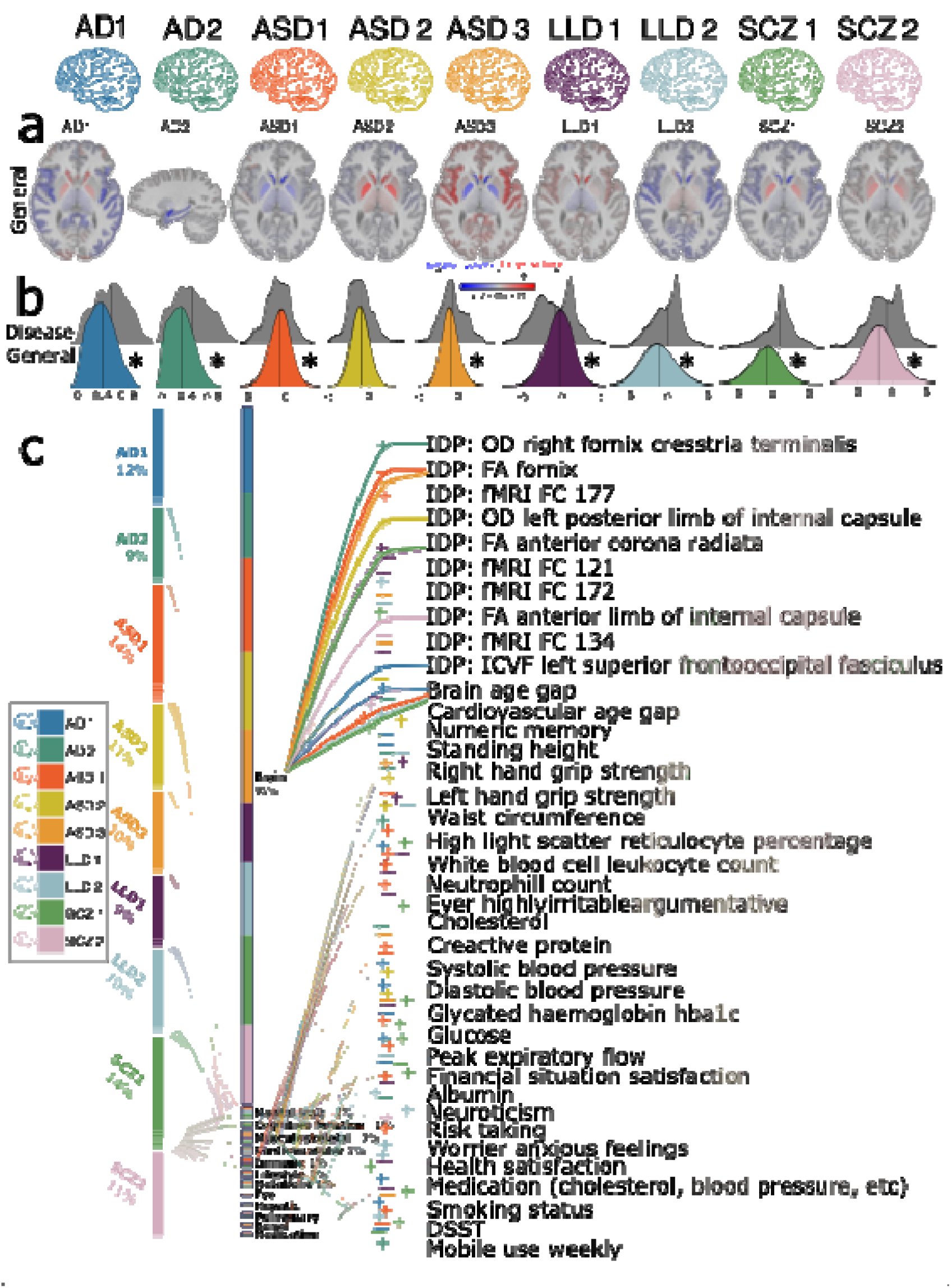
Phenome-wide associations of the nine DNEs. **a**) The neuroanatomical patterns of the nine DNEs were manifested in the UKBB general population and were concordant with the patterns initially derived from the original disease populations^12,2,1,13^. A linear regression model was applied to the 119 gray matter regions of interest (ROIs) derived from T1-weighted MRI data while accounting for various covariates. We present the *β* coefficients of the ROIs that withstood the Bonferroni correction. Positive correlations are depicted using warm reddish colors, while cold blue colors represent negative correlations. For AD2, we showed the sagittal view to visualize the hippocampus and medial temporal lobe. Sample sizes range from 40,534 to 40,981 to derive these results. **b)** The nine DNEs are over-expressed (i.e., a higher mean of the DNE score in the population) and under-expressed (i.e., a lower mean of the DNE score) in the general population compared to the disease populations. The kernel density estimates of the nine DNEs are shown for both the training dataset (gray-colored in patients) and the independent test dataset from the UK Biobank (UKBB). Significant differences that survived the Bonferroni corrections between the training and independent test datasets (two-sampled t-test) are denoted with the symbol *. Sample sizes range from 38,534 to 38,981 to derive these results in UKBB and from 307 to 1510 in the original diseased populations. **c**) Phenome-wide associations (PWAS) between the nine DNEs (left panel) and 611 phenotypes (middle panel) are dominated by brain phenotypic measures. The right panel shows representative phenotypes linked to multiple phenotype categories with the highest statistical significance after the Bonferroni correction (two-sided t-test P-value<0.05/611). A thicker colored line corresponds to a higher value of -log10(P-value). The symbols “+” and “-” represent positive and negative correlations. IDP: imaging-derived phenotype; OD: orientation dispersion; FA: fractional anisotropy; ICVF: intracellular volume fraction; FC: functional connectivity; DSST: digit symbol substitution test. Sample sizes range from 722 to 39,174 after merging the DNEs and these phenotypes. The illustration of the human anatomy is from NIH BIOART Source (https://bioart.niaid.nih.gov/).

The nine DNEs showed different strengths of expression in the general population relative to what is seen in the disease populations from which they were derived. These differences were statistically significant except for ASD2, after correcting for multiple comparisons using the Bonferroni method (**Fig. 2b**). For instance, AD1, characterized by diffuse brain atrophy in the ADNI data, showed a significant under-expression (i.e., a smaller mean of the DNE score) in the general population. Conversely, the subcortical atrophy pattern originally identified in ASD1 from the ABIDE data displayed a significant over-expression (i.e., a larger mean of the DNE score) in the participants from the general population.

We then tested whether the nine DNEs differed between the healthy control and disease groups (AD, ASD, LLD, and SCZ) in the general UKBB population. Compared to the healthy control group (*N*=6390), a small proportion of patients with the four brain diseases showed significant differences (two-sample t-test) for AD1-2 (*N*=23), LLD1-2 (*N*=1329), and SCZ1-2 (*N*=23), except for ASD1-3 (only 6 ASD patients in this population). For example, AD2, characterized by focal medial temporal lobe atrophy, had a significantly lower mean of DNE in the healthy control (0.29±0.19) compared to the AD patient groups (0.45±0.27; P-value=1.1×10^−4^). Detailed statistics for all nine DNEs are presented in **Supplementary Table 1**.

These results proved that the nine DNEs manifest in the general population and convey disease-specific information about the four brain diseases. The contrast in their expression (i.e., over-and under-expression) between the disease-specific and UKBB general populations is expected and underscores their potential relevance as sub-clinical or vulnerability quantitative indices.

### The nine DNEs exhibit phenotypic associations with traits beyond the brain

To delineate their phenotypic landscape, we associated the nine DNEs with 611 phenotypes in UKBB. To avoid circularity, the PWAS did not include the 119 GM ROIs derived from T1-weighted MRI, from which the nine DNEs were derived. Out of the 611 additional clinical traits spanning multiple organ systems, cognition, and lifestyle factors, we discovered 1818 significant associations after applying the Bonferroni correction (P-value < 0.5/611) (**Fig. 2c**, **Supplementary File 3**).

Of the 1818 significant associations, 91% were related to the brain. For example, the mean intracellular volume fraction in the superior frontal-occipital fasciculus derived from the multi-shell NODDI^33^ model was significantly associated with AD1 [*β*=-0.67±0.02, -log_10_(P-value) > 300]. Multiple DNEs were significantly associated with the biological age gap (BAG: AI-predicted age minus chronological age) of the brain [e.g., SCZ2: *β*=0.19±0.01, -log_10_(P-value)=50.10]. Furthermore, 2% of the phenotypes related to the musculoskeletal system were associated with the nine DNEs. The nine DNEs were also largely associated with many phenotypes related to mental health (1%). For example, the neuroticism score was significantly associated with LLD2 [*β*=-1.09×10^−2^±2.42×10^−3^, -log_10_(P-value)=5.20] (**Supplementary File 3**). A previous study suggests distinct neural mechanisms between older individuals with neurotic and non-neurotic depression, indicating diverse biological pathologies contributing to varying clinical presentations of LLD^34^.

We conducted two sensitivity analyses to validate the main PWAS results. We obtained high concordance rates in split-sample (98.03%) and sex-stratified analyses (93.98%). Detailed results can be found in **Supplementary Text 2** and **Supplementary File 4** and **5** for split-sample and sex-stratified analyses.

As anticipated, 91% of the significant associations were linked to the brain, given that the DNEs were derived from regional brain volumetrics in brain disease-specific populations. However, it is noteworthy that these phenotypic associations extended beyond the brain, providing evidence for the significant associations between the brain and other organ systems. This brain-body connection was consistent with previous literature on multi-organ research using imaging and genetic data^17,35,36^.

### Genome-wide associations identify 66 genomic loci

At the genome-wide significance level (P-value<5×10^−8^), GWAS using PLINK for a linear regression model identified 10, 8, 5, 21, 9, 1, 3, 3, and 6 genomic loci significantly associated with AD1, AD2, ASD1, ASD2, ASD3, LLD1, LLD2, SCZ1, and SCZ2, respectively (66 in total, **Fig. 3a**, and **Supplementary File 6**). At a more stringent significance level (P-value<5×10^−8^/9), 31 loci passed the Bonferroni correction, as annotated in **Fig. 3a (**query date: 2^nd^ June 2023, via FUMA version: v1.5.4). To support the robustness of our GWAS, we estimated the intercept of linkage disequilibrium score regression (LDSC)^37^ and obtained intercepts of 0.9997±0.0093, 1.0325±0.0091, 0.9962±0.0095, 1.0172±0.0099, 1.0117±0.0085, 1.0135±0.008, 1.0162±0.0094, 1.0101±0.0087, and 1.0124±0.0095 for the nine DNEs. All intercepts were close to 1, indicating no substantial genomic inflation in our GWASs. The Manhattan and QQ plots of the nine GWASs are presented in **Supplementary Figure 1-9** and are also publicly available on the MEDICINE knowledge portal: https://labs-laboratory.com/medicine/.

**Figure 3:**
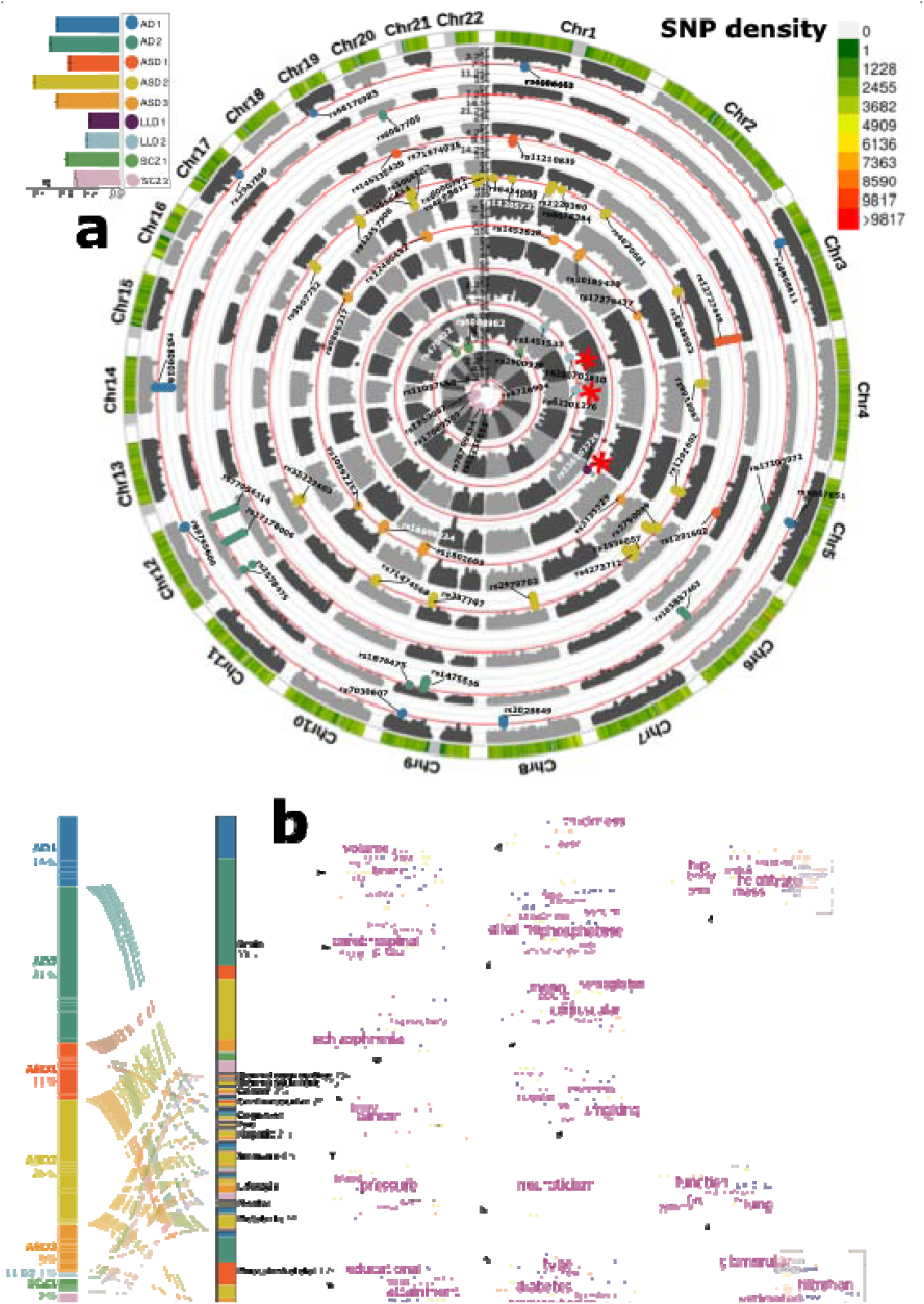
Genome-wide associations of the nine DNEs. **a**) Genome-wide associations identified 66 (10, 8, 5, 21, 9, 1, 3, 3, 6 for the nine DNEs) genomic loci (P-value<5×10^−8^) associated with the nine DNEs. Using the top lead SNP, we denoted each genomic loci linked to the 9 DNEs. Red * symbols indicate that the locus LD has not been previously associated with any trait in the EMBL-EBI GWAS catalog. The left legend indicates the significant SNP-based heritability (*h*^2^) for the nine DNEs; the right legend represents the SNP density of our genetic data throughout the human genome. GWAS was performed using the Genome Reference Consortium Human Build 37 (GRCh37). These GWASs included 31,976 participants of European ancestry. **b**) Phenome-wide association query of the previously identified genomic loci (left panel) in the EMBL-EBI GWAS Catalog (via FUMA 1.4.2) shows a brain-dominant genetic architecture. We categorized all clinical traits (middle panel) into several high-level categories linked to multiple organ systems, neurodegenerative and neuropsychiatric disorders, lifestyle factors, etc. We then show the keyword cloud plots for each category (right panel). The illustration of the human anatomy is from NIH BIOART Source (https://bioart.niaid.nih.gov/).

All DNEs are significantly heritable (0.24<*h*^2^<0.66, P-value<1×10^−10^) after Bonferroni correction (**Fig. 3a**, **Supplementary Table 2**). We employed the GCTA^38^ software to estimate *h*^2^, acknowledging that previous research has demonstrated variations in the magnitude of *h*^2^ estimates based on the choice of methods. The *h*^2^ estimates obtained through GCTA or similar methods may underestimate SNP-based heritability (i.e., missing heritability), potentially due to unaccounted factors such as gene-environment interactions, epistasis, and rare variant effects.

We further investigated the significant genomic loci by mapping them to protein-encoding genes and examining their functional implications through expression quantitative trait loci (eQTL) mapping. **Supplementary Figure 10** presents the regional Manhattan plot for the most significant genomic locus associated with each DNE. For example, we identified a locus associated with ASD2 (top lead SNP: rs3068507 at 20q11.21) and a neighboring locus associated with SCZ1 (top lead SNP: rs6088962 at 20q11.21), both of which mapped to the *MYLK2* gene (**Supplementary Figure 10d and h**). The *MYLK2* gene encodes a myosin light chain kinase primarily expressed in adult skeletal muscle.

We conducted seven sensitivity analyses to validate the main GWAS results. We obtained perfect concordant rates (100%) and similar genomic inflation factors using fastGWA^39,40^ for a generalized linear mixed model in the nine GWAS. Specifically, we excluded the related individuals (up to 2^nd^-degree) in our genetic quality check pipeline; mixed effect models like fastGWA can detect additional genetic relatedness without excluding individuals by using a sparse genomic relationship matrix, significantly reducing the computational burden for large-scale GWAS. This sensitivity check further supports that no substantial genomic inflation exists in our main GWASs. We also observed high concordance rates in split-sample, sex-stratified (63.26-92.54%), and longitudinal GWAS analyses (100%, *N*=1116), but the concordance rates were relatively low in non-European ancestry GWAS, independent ADNI whole-genome sequencing GWAS, and six case-control GWAS^41–46^ of neurodegenerative and neuropsychiatric disorders from the Psychiatric Genomics Consortium (PGC)^47^ (**Supplementary Table 3a)**. The sample sizes for the non-European (*N*=4783) and ADNI (*N*=1555) samples are small; the case-control GWAS from the PGC may overlook the heterogeneity within each disease. Detailed results are presented in **Supplementary Text 3** for the sensitivity results, **Supplementary File 7-13** for replicated SNPs/loci, and **Supplementary Figure 1-9** for Manhattan and QQ plots and the LDSC intercepts.

### The genetic associations of the nine DNEs parallel their phenotypic associations

We performed a phenome-wide look-up analysis to understand the phenotypic associations in the literature for the identified genomic loci in our GWAS.

In total, 2525 clinical traits were associated with genetic variants in our GWAS, including traits linked to multiple organ systems, cognition, and lifestyle factors (**Fig. 3b** and **Supplementary File 14**). The genomic loci were largely associated with clinical traits of the brain (53%), musculoskeletal (17%), immune system (6%), neurodegenerative (1%), and neuropsychiatric (1%) diseases. For example, AD2 genomic loci were largely associated with traits related to the brain (565 out of 781, e.g., brain IDPs), musculoskeletal (133/781, e.g., standing height), immune (17/781, e.g., reticulocyte count), cognition (15/781, e.g., cognitive performance), lifestyle factors (13/781, e.g., smoking), and neurodegenerative traits (1/781, i.e., neurofibrillary tangles).

The findings closely align with the phenotypic associations observed in **Fig. 2c**, reinforcing that the DNEs share genetic determinants linked to organs beyond the brain, lifestyle factors, and cognition.

### The genetic correlation of the nine DNEs

To understand the shared genetic underpinnings, we estimated the genetic correlation^37^ (*g_c_*) between the nine DNEs, the BAG of nine human organ systems^17^, and six brain diseases (AD, attention-deficit/hyperactivity disorder, autism spectrum disorder, bipolar, obsessive-compulsive disorder, schizophrenia; **Supplementary Table 3a**) from PGC and four lifestyle factors and cognitive scores (**Supplementary Table 3b**).

We first estimated the *g_c_* between each pair of DNEs (**Fig. 4a**). Numerous DNEs exhibited strong genetic correlations among each other. The highest positive genetic correlation was obtained between ASD2 and SCZ1 (*g_c_*=0.57±0.04); the highest negative genetic correlations were obtained between ASD2 and ASD1 (*g_c_*=-0.55±0.04), and between ASD3 and SCZ1 (*g_c_*=-0.51±0.05). We also observed a substantial alignment between the phenotypic correlation (*p_c_*) and the genetic correlation of pairwise DNEs, supporting the long-standing Cheverud’s Conjecture^48^. However, we identified two exceptions where the observed phenotypic and genetic correlations exhibited opposite directions. ASD1 and ASD3 showed a negative phenotypic (*p_c_*=-0.39±0.08) but a positive genetic correlation (*g_c_*=0.21±0.05); ASD1 and LLD1 showed a negative phenotypic (*p_c_*=-0.34±0.09) but a positive genetic correlation (*g_c_*=0.16±0.07) (**Supplementary Table 4**). This implies that non-genetic factors, such as lifestyle and environmental factors, may exert opposite influences on the two DNEs. We performed additional MAGMA gene-set analysis^49^ to test the genetic similarity between ASD2 and SCZ1. The most significant biological pathway underlying ASD2 is the negative regulation of locomotion (GO 0040013, P-value=2.27×10^−5^, β=0.22±0.02), implicated in biological processes that stop, prevent, or reduce the frequency, rate, or extent of locomotion of a cell or organism. The most significant biological pathway for SCZ1 is the negative regulation of neurotransmitter transport (GO 0051589, P-value=1.41×10^−5^, β=0.22±0.02), involved in biological processes that downregulate the directed movement of a neurotransmitter into, out of, or within a cell. In particular, the latter supports the involvement of dopamine and glutamate, two major neurotransmitters in the central nervous system, in schizophrenia^50^.

**Figure 4:**
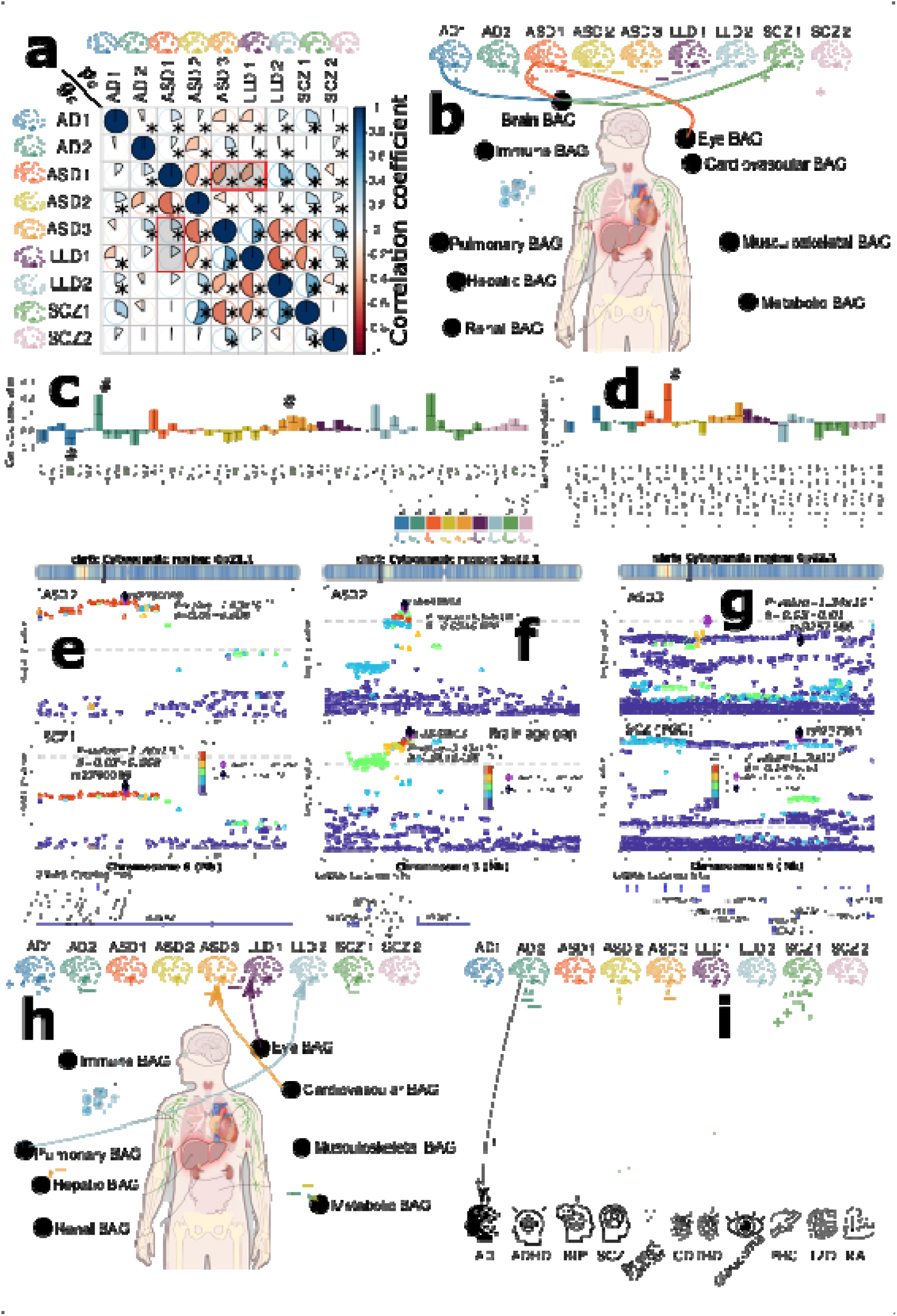
The genetic correlation, colocalization, and causal networks of the nine DNEs. **a**) The genetic correlation between two DNEs (*g_c_*, lower triangle) mirrors their phenotypic correlation (*p_c_*, upper triangle). Red-shadowed rectangles highlight two exceptions. The symbol * indicates significant results after the Benjamini-Hochberg correction. The symbol **#** indicates nominal significance. **b**) genetic correlations between the nine DNEs and nine biological age gaps (BAG) for nine human organ systems^17^. **c**) genetic correlations between the nine DNEs and six neurodegenerative and neuropsychiatric disorders. The bar plots display the estimated mean genetic correlation along with its standard error. **d**) genetic correlations between the nine DNEs and four traits related to lifestyle factors and cognition. The bar plots display the estimated mean genetic correlation along with its standard error. A two-sided p-value is used to determine statistical significance. **e**) genetic colocalization was evidenced at one locus (6p21.1) between ASD2 and SCZ1. The signed PP.H4.ABF (0.92) denotes the posterior probability (PP) of hypothesis H4, which suggests that both traits share the same causal SNP (rs2790099). A positive PP indicates concordant *β* values for both DNEs, while a negative PP implies opposite *β* values. **f**) genetic colocalization was evidenced at one locus (3p.22.1) between ASD2 and brain BAG: PP.H4.ABF=0.95 with the cause SNP rs5848503. **g**) genetic colocalization was evidenced at one locus (6p.22.1) between ASD3 and SCZ case-control GWAS^45^ from PGC (European ancestry): PP.H4.ABF=0.82 with the cause SNP rs9257566. **h**) the causal network of the nine DNEs with the eight multi-organ BAGs. Solid arrow lines (from the exposure to the outcome variables) indicate significant causal relationships after the Benjamini-Hochberg correction; dotted arrow lines show nominal significance (P-value<0.05). The symbols **+** (OR>1 and *g_c_*>0) and **–** (OR<1 and *g_c_*<0) represent a positive relationship between the two traits. **i**) the causal network of the nine DNEs with the eleven chronic diseases (e.g., AD, ADHD, BIP, and SCZ from PGC). Abbreviation: AD: Alzheimer’s disease; ADHD: Attention-deficit/hyperactivity disorder; ASD: autism spectrum disorder; BIP: bipolar disorder; SCZ: schizophrenia; OCD: Obsessive-compulsive disorder; RA: rheumatoid arthritis; CD: Crohn’s disease; T2D: type 2 diabetes; IBD: inflammatory bowel disease; PBC: Primary biliary cirrhosis. The illustration of the human anatomy is from NIH BIOART Source (https://bioart.niaid.nih.gov/).

Between the nine DNEs and the BAGs across nine human organ systems, we found significant genetic correlations between AD1 (*g_c_*=0.23±0.05), ASD1 (*g_c_*=0.44±0.05), LLD2 (*g_c_*=0.24±0.07), SCZ1 (*g_c_*=0.26±0.06), and the brain BAG, and between ASD1 and the eye BAG (*g_c_*=0.19±0.07) (**Fig. 4b** and **Supplementary Table 5**).

Finally, we also found a marginally significant genetic correlation between AD2 and AD (*g_c_*=0.22±0.12), AD1 and bipolar disorder (BIP, *g_c_*=-0.08±0.04), and ASD3 and BIP (*g_c_*=0.09±0.04) using GWAS summary statistics from PGC (**Fig. 4c** and **Supplementary Table 6a**). We observed a nominal genetic correlation signal between ASD1 and reaction time (*g_c_*=0.35±0.15; **Fig. 4d** and **Supplementary Table 6b**).

In summary, the nine DNEs demonstrate substantial genetic correlations among themselves and with organ systems beyond the brain. These findings highlight the interconnectedness of the neuroanatomical patterns and genetic determinants across multiple body systems and diseases, suggesting shared underlying disease mechanisms and potential pleiotropic effects.

### The genetic colocalization of the nine DNEs

To seek the shared causal variants between two clinical traits (e.g., AD1 vs. LLD2), we performed Approximate Bayes Factor colocalization^51^ analyses between the nine DNEs (**Fig. 4e**), with the nine BAGs (**Fig. 4f**), and the six brain disorders from PGC^47^ (**Fig. 4g**).

Among the nine DNEs, we detected 44 causal variants (SNPs) exhibiting significant colocalization signals. We showcased the shared causal variant (rs2790099 at 6p21.1) between ASD2 and SCZ2 with a PP.H4.ABF (Approximate Bayes Factor)=0.92 (**Fig. 4e**), which examines the posterior probability (PP) to evaluate the hypothesis, the presence of a single shared causal variant associated with both traits within a specific genomic locus. This causal SNP was mapped to the *RUNX2* gene. The loss of function in *RUNX2* causes a rare autosomal dominant skeletal disorder – cleidocranial dysplasia^52^, but it was implicated in ASD or SCZ in previous literature.

Between the nine DNEs and nine BAGs, we identified 13 causal variants (SNPs) exhibiting significant colocalization signals. We showcased the shared causal variant (rs5848503 at 3p22.1) between ASD2 and the brain BAG with a PP.H4.ABF=0.95 (**Fig. 4f**). One mapped gene in this locus is the *MOPB* gene, which encodes the myelin-associated oligodendrocytes basic protein and is actively involved in the structural constituent of the myelin sheath and nervous system development. This gene was previously implicated in ASD using single-cell genomics^53^, SCZ^54^, amyotrophic lateral sclerosis, and Parkinson’s disease^55^.

Between the nine DNEs and six brain diseases from PGC, we identified 6 causal variants (SNPs) exhibiting significant colocalization signals. We showcased the shared causal variant (rs9257566 at 6p22.1) between ASD3 and SCZ with a PP.H4.ABF=0.82 (**Fig. 4g**). In this locus, multiple olfactory receptor (OR) genes and the dysfunction of the olfactory system were implicated in SCZ^56,57^ and ASD^58,59^. For instance, the *OR2J2* and *OR2J3* genes are two protein-coding genes in copy number variants associated with SCZ using microRNA data^60^. The causal SNP (rs9257566) was associated with SCZ and brain IDP, such as white matter microstructural measures (**Supplementary Figure 11**). Besides those two conditions, AD has also been widely associated with early olfactory dysfunction^61,62^. The sensitivity checks on the prior probability (*p12*) for the three illustrations are shown in **Supplementary Figure 12a-c**. The causal variant, cytogenetic region, and their colocation signal direction (based on β coefficients) are presented in **Supplementary Figure 13a-c**. Detailed results are shown in **Supplementary File 15-17**.

The genetic colocalization of the nine DNEs revealed causal genetic variants, indicating that the same genomic regions may causally influence the expression of these AI-derived endophenotypes.

### The causal relationship of the nine DNEs

We applied bidirectional two-sample Mendelian randomization analyses^63^ to depict a causal network between the nine DNEs, the eight BAGs (excluding the brain BAG), and eleven chronic diseases spanning the whole-body system.

For each pair of DNEs, as the GWAS populations completely overlapped, conducting two-sample Mendelian randomization was not feasible^64^. Alternatively, the split-sample GWAS did not yield sufficient statistical power due to the limited number of instrumental variables (VI) available (< 6 SNPs).

Among the nine DNEs and eight BAGs, we found potential causal effects of the eye BAG on LLD1 [P-value=4.57×10^−3^, OR (95% CI) = 1.14 (1.04, 1.24), number of SNPs=16], the cardiovascular BAG on ASD3 [P-value=6.04×10^−3^, OR (95% CI) = 1.16 (1.04, 1.29), number of SNPs=34], and the pulmonary BAG on LLD2 [P-value=1.98×10^−3^, OR (95% CI) = 1.14 (1.04, 1.25), number of SNPs=49]. No significant causal signals persisted after the Benjamini-Hochberg correction in the inverse analyses (**Fig. 4h**). Details of the results, including all five different Mendelian randomization estimators, are shown in **Supplementary File 18**.

Between the nine DNEs and eleven chronic diseases, encompassing brain-related conditions and diseases affecting other organs, we identified a potential causal effect from AD2 to AD using the GWAS summary statistics from PGC – the largest sample size (*N*=1,126,536 individuals) in the AD case-control study [P-value=1.74×10^−4^, OR (95% CI) = 1.25 (1.11, 1.40), number of SNPs=7] (**Fig. 4i**). We didn’t detect a causal link in the opposite direction from AD to AD2. This aligns with the endophenotype hypothesis, suggesting that DNEs are situated within the causal pathway from genetics to external phenotypes, such as AD diagnosis and cognitive decline. Details of the results, including all five different Mendelian randomization estimators, are shown in **Supplementary File 19**. The results of the sensitivity check are presented in **Supplementary Figure 14-17**.

The Mendelian randomization results further emphasize the potential benefits of overall organ health for brain-related conditions. This highlights the interconnectedness between various organ systems and the brain, underscoring the significance of a holistic health and disease prevention approach.

### The nine DNEs predict14 systemic diseases and mortality

We investigated the added prediction power of the nine DNEs and their respective PRS for 14 systemic diseases based on the ICD-10 code (**Supplementary Table 7-8**), 8 cognitive scores(**Supplementary Table 9**), and mortality outcomes (i.e., the date of death). Of note, we did not perform prediction on the four brain diseases (AD, ASD, LLD, and SCZ) due to the small and highly imbalanced sample sizes in the UKBB general population. As anticipated, the prediction performance across all tasks was modest, considering that the DNEs were derived from specific disease populations.

In addition to commonly available features, such as age and sex, we found that AD1, ASD1, LLD1, SCZ1, and SCZ2 provided additional prediction power (i.e., incremental *R*^2^) for many disease categories. Across the 14 disease categories, the DNEs showed higher incremental *R*^2^ in mental and behavioral disorders (ICD-10 code: F group) and diseases linked to the central nervous system (ICD-10 code: G group) than other disease categories, proving that the nine DNEs in the general population capture disease-related effects of the four brain diseases. Combining all nine DNEs further improved the incremental *R*^2^, especially in mental and behavioral disorders (*R*^2^=1.01%, P-value=1.74×10^−5^) and diseases linked to the central nervous system (*R*^2^=0.63%, P-value=1.33×10^−5^) (**Fig. 5a**). The incremental *R*^2^ values for all tests are shown in **Supplementary Table 7**. All metrics, encompassing sample sizes and incremental R2 for both the null and alternative models, are outlined in **Supplementary File 20**. Results using only the PRS target population are presented in **Supplementary Figure 18**.

**Figure 5:**
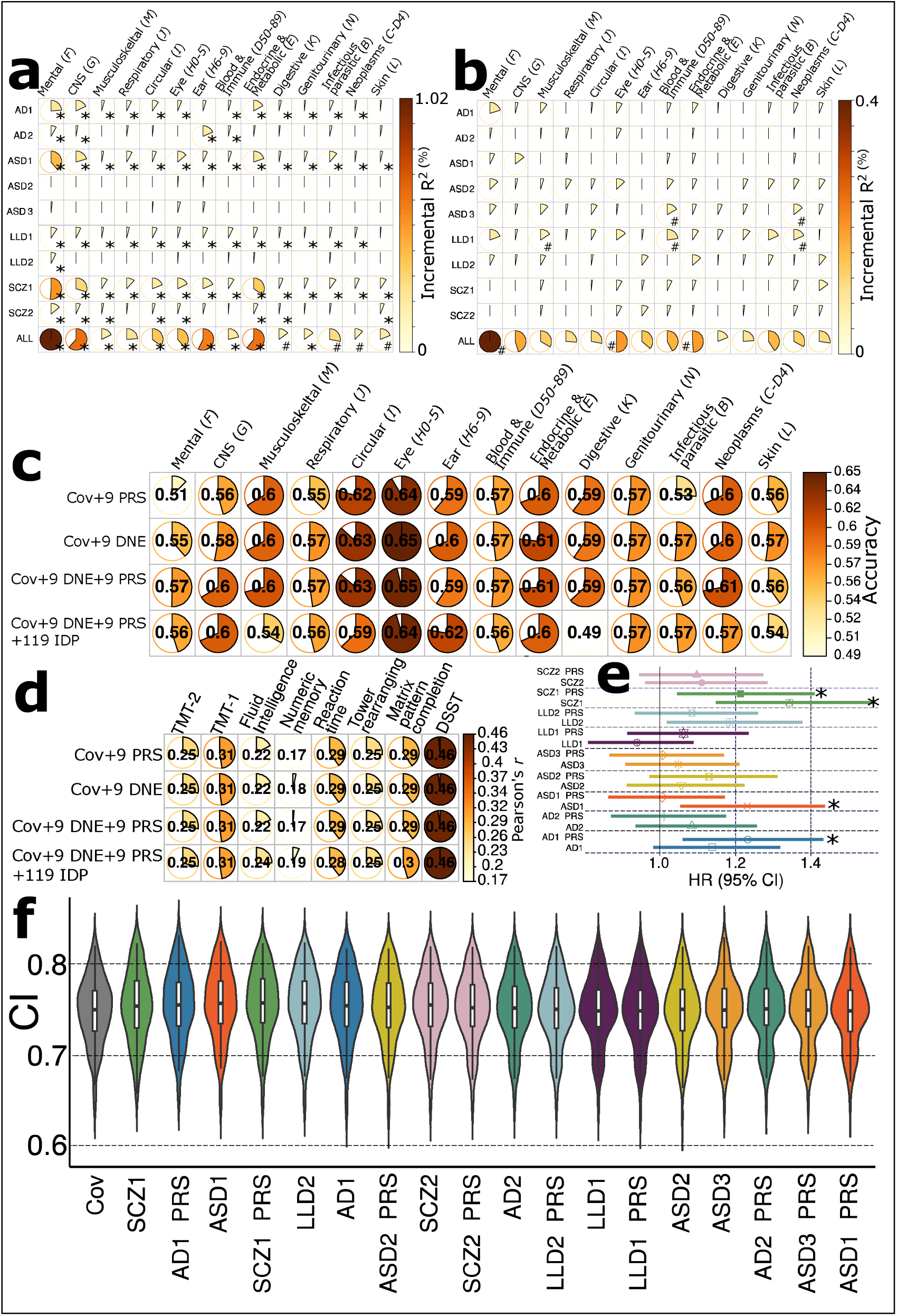
Additional prediction power of the nine DNEs and PRSs for 14 systemic diseases, cognition, and mortality outcomes. **a**) The incremental R-squared (*R*^2^) values of the nine DNEs for predicting 14 systemic disease categories were assessed using the entire UKBB sample, with *N*=39,178 participants as independent test data. The results focusing only on the PRS target population (*N*=15,891) can be found in **Supplementary Figure 18**. “ALL” indicates the incremental *R*^2^ contributed by combining the nine DNEs. **b**) The incremental *R*^2^ of the PRS of the nine DNEs to predict 14 systemic diseases based on the ICD-10 code using only the PRS target sample. **c**) In the PRS target sample, disease classification accuracy from the independently hold-out test data (*N*=5581) was assessed using nested cross-validated support vector machines in the training/validation/test data (*N*=10,000) by fitting various sets of features (Cov indicates age and sex). **d**) In the PRS target sample, cognitive score prediction accuracy (Peasrson’s *r*) from the independently hold-out test data (3632<*N*<5570) was assessed using nested cross-validated support vector regression models. **e**) The SCZ1, SCZ1-PRS, AD1-PRS, and ASD1 show significant associations with the risk of mortality in the PRS target sample. Age and sex were included as covariates in the Cox proportional hazard model. **f**) The nine DNEs and PRSs were cumulatively included as features in cross-validation for mortality risk prediction. The symbol * indicates significant results that survived the Benjamini-Hochberg correctionThe symbol **#** indicates nominal significance. All P values are two-sided. HR: hazard ratio; CI: concordance index; DSST: digit symbol substitution test; TMT: trail-making test. The box plots display the mean of the CIs, while the violin plots illustrate their distribution during cross-validation.

Compared to the nine DNEs, the nine PRSs provided smaller additional prediction power. For example, the PRS for ASD3 explained an additional 0.05% of the variance (incremental *R*^2^) in diseases associated with the blood and immune systems (P-value=0.03), as well as neoplasms (P-value=0.04). Combining all nine PRSs improves the incremental *R*^2^, particularly in mental and behavioral disorders (*R*^2^=0.3%, P-value=0.047). No results survived multiple comparisons using the Benjamini-Hochberg method (**Fig. 5b**). Detailed results are shown in **Supplementary Table 8** and **File 21**.

We assessed the prediction ability of support vector machines (SVM) at the individual level to classify the 14 disease categories. The highest performance was observed for eye diseases (ICD-codes: H0-5). Age and sex significantly enhanced the classification accuracy. Including age and sex alongside the 9 DNEs and 9 PRSs boosted the classification accuracy from 0.55 to 0.65. The inclusion of PRSs, DNEs, and the combination of both, along with age and sex as features, resulted in improved classification accuracy for mental and behavioral disorders. For example, the accuracy increased from 0.51 to 0.55 and 0.57 for features of age and sex, 9 DNEs, and 9 PRSs, incrementally (**Fig. 5c**). These findings highlight the added value of incorporating the nine DNEs and PRSs in predicting these disease categories. Detailed results are shown in **Supplementary Table 9a**. The full evaluation metrics of the cross-validated and independent test results are presented in **Supplementary File 22**.

We examined the predictive capability of these feature sets in estimating 8 cognitive scores. The digit symbol substitution test (DSST) demonstrated the highest performance, reaching a Pearson’s *r* of 0.46 (e.g., P-value=3.9×10^−189^ and MAE=3.75 for the feature set of 9 DNE plus covariates) in the independent test dataset using all different sets of features (**Fig. 5d**). The combination of DNEs and PRSs did not notably enhance prediction accuracy. However, contrary to the 14 systemic disease classifications, incorporating the 119 GM imaging-derived phenotypes alongside DNEs and PRSs resulted in a 2% increase in the prediction accuracy for fluid intelligence, measured by Pearson’s *r* (**Supplementary Table 9b**). The full evaluation metrics of the cross-validated and independent test results are presented in **Supplementary File 23.**

Finally, we evaluated the prediction power of the nine DNEs and PRSs for mortality risk prediction using the Cox regression. Among these, SCZ1, SCZ1-PRS, ASD1, and AD1-PRS were significantly associated with the risk of mortality (**Fig. 5e** and **Supplementary Table 10a**). Adding SCZ1, AD1-PRS, ASD1, and SCZ1-PRS to age and sex further improved the prediction, but the performance decreased afterward (**Fig. 5f**). Lastly, incorporating the nine DNEs from the second scan of 1348 participants into the model slightly increased the statistical significance and the HR (**Supplementary Table 10b** and **c**).

## Discussion

This study investigated the manifestation of nine disease-related brain endophenotypes – derived from four case-control studies via semi-supervised AI methods – in the general population of 39,178 participants in UKBB. We assessed commonalities and differences among the nine DNEs, their genetic correlates in the general population, their relationships with the multiple human organ systems, and their predictive capacity for 14 systemic disease categories and mortality. Our findings demonstrate the potential of the nine AI-derived DNEs in identifying high-risk individuals within the general population prone to developing the four major brain disorders.

### Shared neuroanatomical patterns and genetic determinants

Understanding the shared disease mechanism of neurodegenerative and neuropsychiatric diseases is a complex and ongoing challenge in medical research^15,16,41,43–46,65^. Our results suggest that shared underlying mechanisms and genetic factors may contribute to the development and progression of these disorders. This notion of shared disease mechanisms across the four major neurodegenerative and neuropsychiatric diseases, namely ASD, SCZ, LLD, and AD, has garnered considerable attention and reshaped our understanding of these conditions^15,16^.

Despite the inherent heterogeneity among neuroanatomical patterns observed in different brain diseases (**Fig. 2a**), a notable commonality exists regarding their manifestations, which might emanate from underlying mechanisms sharing neuropathologic characteristics and pathways. As an illustration, AD1, LLD2, and SCZ1 exhibited a negative correlation (brain atrophy) with widespread cortical volumes, such as the bilateral insula and middle frontal gyrus. This aligns with expectations, considering that the UKBB population includes individuals primarily from mid to late life (above 45 years). From an etiological standpoint, various factors can contribute to the global cortical volume reduction within the general population, with late-onset neurodegenerative and neuropsychiatric disorders and aging exerting a significant impact. Likewise, ASD2 and SCZ2 exhibited a positive association with the basal ganglia, including the globus pallidum. This could imply the existence of potential protective genetic or environmental factors that collectively contribute to the concept of “brain reserve”, which might mitigate volume loss in a portion of the general population. Notably, we previously revealed that individuals predominantly expressing SCZ2 exhibited higher levels of education^23^ and higher remission rates than those primarily influenced by SCZ1. Alternatively, these volume increases might reflect neuropathologic mechanisms, such as disrupted connectivity, which are not necessarily associated with neurodegenerative and neurodevelopmental components related to relatively lower brain volumes. Ultimately, another significant aspect to consider involves the impact of medications prescribed by clinicians, affecting both disease-specific and general populations. Research indicates that antipsychotic medications, commonly prescribed for most individuals with schizophrenia and some with ASD and AD, have been shown to delay brain volume loss in the basal ganglia^66^ and pallidum^67^.

The commonalities in neuroanatomical patterns across brain diseases can be attributed to several factors. First, shared genetic factors may influence brain structure and function^30,31^, contributing to similar neuroanatomical alterations across different diseases. Genetically, this was largely evidenced by our genetic correlation (**Fig. 4a**) and colocalization results (**Fig. 4e**). For instance, ASD2 showed prominent positive genetic overlap with SCZ1. Historically, there has been a long-standing association between ASD and SCZ, leading to the notion that autism could be a form of “childhood schizophrenia“^68^. This conceptual link between the two conditions has been debated and discussed. Previous case-control neuroimaging studies demonstrated divergent structural and functional brain patterns in individuals with ASD compared to those with SCZ^69,70^, largely ignoring the neuroanatomical heterogeneity within each condition. Genetic variants that impact key signaling pathways, synaptic function, and neuronal connectivity^71,72^ could influence multiple disease phenotypes, leading to overlapping neuroanatomical patterns.

AD1 also shared genetic similarities with LLD1, both characterized by spatially extensive brain atrophy and increased brain age. Previous studies found a higher prevalence of depressive symptoms and LLD in individuals with AD compared to the general population^73–75^. The relationship between AD and LLD likely involves multiple factors and may be bidirectional. On the one hand, LLD may increase the risk of developing AD or accelerate the progression of cognitive decline in individuals already affected by AD. On the other hand, AD-related changes in the brain, such as neuroinflammation and neurochemical imbalances, may worsen depressive symptoms in individuals with LLD.

Recognizing the shared disease mechanisms across the four brain diseases underscores the importance of a broader perspective on clinical presentations and underlying biological mechanisms. This understanding is important for developing targeted and personalized approaches to patient care, leading to more effective treatments and interventions. Future research may explore disease heterogeneity to transcend traditional classification boundaries and identify data-driven subtypes or dimensions within a transdiagnostic framework^76^.

### Beyond the brain

Our findings strongly concur with a paradigm shift in treating brain diseases. While the conventional approach has predominantly concentrated on interventions targeting the brain, emerging evidence highlights the critical importance of considering the broader systemic and environmental factors that influence disease onset and progression.

Unraveling the intricate interconnections between the brain and other organ systems is crucial in broadening our understanding of brain diseases, as demonstrated by our findings and other findings. The brain does not function in isolation but interacts with and is influenced by various physiological systems throughout the body. Our results showed a close genetic association and causality between the DNEs and the eye, cardiovascular, and pulmonary systems (**Fig. 4b, f**, and **h**). These findings parallel previous literature. For instance, eye-related pathological changes have been revealed to mirror early signs of neurological and neuropsychiatric conditions^77^. The nervous and cardiovascular systems – the heart-brain axis – are intricately linked, with brain regions controlling heart function via sympathetic and parasympathetic pathways^78^. Dysfunctions in one system can affect the other’s function, resulting in brain and cardiovascular diseases. The immune system plays a crucial role in modulating inflammation and neuroinflammation, which are implicated in many brain disorders, such as AD^79^, SCZ^80^, and depression^81^. Similarly, the gut-brain axis highlights the bidirectional communication between the gut microbiota and the brain, with emerging evidence linking alterations in the gut microbiome to brain diseases such as Parkinson’s disease^82^ and depression^83^. Understanding and targeting these systemic interactions can modulate disease processes and improve treatment outcomes.

Furthermore, considering environmental and lifestyle factors is essential in treating brain diseases^84^. Our previous work^17^ has shown that the BAGs of nine human organ systems comply with Cheverud’s Conjecture: the phenotypic correlation of two BAGs mirrors their genetic correlations. However, herein we showed that the phenotypic correlation between two DNEs (e.g., ASD1 vs. LLD1) did not reflect their underlying genetic correlation (**Fig. 4a**), indicating potentially strong environmental and lifestyle factors that exert opposite effects on the two DNEs. Furthermore, an interesting observation from our study was that the heritability estimate (*h*^2^) of early-onset diseases, such as ASD, was higher than that of late-onset diseases, such as LLD, within the three neuropsychiatric disorders. This finding suggests that genetic factors play a more prominent role in developing ASD at a younger age. In contrast, other factors, such as environmental influences, socioeconomic factors, and lifestyle choices, may have a stronger impact on developing LLD later in life. These differential heritability patterns shed light on the complex interplay between genetic and non-genetic factors in the underlying disease mechanism of neurodegenerative and neuropsychiatric disorders across different stages of life. These heritability patterns aligned with a previous study that examined multiple GWAS drawn from more than 200,000 patients for 25 brain-associated disorders and 17 phenotypes^16^.

In conclusion, going beyond the brain is crucial for understanding and treating brain diseases. By considering the connections between the brain and other organ systems, understanding the impact of environmental and lifestyle factors, and harnessing the power of advanced AI technologies, we can develop more effective and personalized approaches to prevent, diagnose, and treat brain diseases.

### AI-derived DNEs for precision diagnostics

The present study leverages cutting-edge, semi-supervised AI methods^24^ and open science advancements to enhance our understanding of disease heterogeneity in neurodegenerative and neuropsychiatric disorders. In this context, implementing these AI-derived DNEs at early disease or preclinical stages – in the general population – may facilitate the identification of individuals at risk, and the initiation of proactive interventions before the onset of noticeable symptoms, likely leading to more effective treatments and interventions and better outcomes.

The proposed AI-derived DNEs capture intricate brain structure and function variations, often subtle and spatially complex, which traditional diagnostic methods and case-control studies may overlook. By quantifying the neuroanatomical patterns associated with specific brain disorders, DNEs may offer a personalized disease vulnerability assessment, inform interventions at early preclinical stages, and potentially prevent or delay the onset of symptoms. At the individual level, integrating genetic information (i.e., PRSs) with DNEs significantly improves prediction performance for 14 systemic diseases and mortality outcomes (**Fig. 5**). In addition, our Mendelian randomization analyses supported the well-established endophenotype hypothesis in genetic psychiatry^25^ – endophenotype in psychiatric disorders resides inside the causal pathway from underlying genetics to their exo-phenotypes (i.e., the disease syndrome itself), thereby being closer to its disease mechanisms. We found that AD2, characterized by focal medial temporal lobe atrophy, exerted a causal relationship with AD. However, we did not find evidence of a reverse causal relationship, suggesting that the underlying genetics may influence the development of AD through the DNE, although it may not be the exclusive pathway contributing to the disease. This highlights the role of genetics in influencing the disease process, particularly through the identified DNE, shedding light on potential pathways and mechanisms involved in AD development.

### Limitation

The present study has several limitations. Firstly, the genetic analysis focused exclusively on common genetic variants. Future investigations should explore the contribution of rare variants to these brain diseases. Secondly, it is important to recognize that our GWAS analyses predominantly involved participants of European ancestry, limiting the generalizability of the genetic findings to other populations with different ancestral backgrounds. Further research efforts are necessary to collect more diverse genetic data and include underrepresented racial and ethnic groups to enhance the generalizability of the findings. Additionally, the validation of the nine DNEs would benefit from additional longitudinal analyses. Fortunately, ongoing efforts to collect longitudinal brain MRI data in UKBB^85^ hold promise for providing valuable insights to the scientific community and advancing the field of precision medicine. Furthermore, more advanced methodological developments will be developed and integrated within our semi-supervised clustering learning framework to alleviate potential domain shifts^86^. Finally, our future research will incorporate multimodal imaging to generate DNEs sensitive to a broader range of brain alterations, including macrostructural volumetric changes, functional disruptions, and structural network abnormalities.

### Outlook

Together, our AI-derived DNEs have emerged as novel instruments for precision medicine. By capturing the complexity and heterogeneity of brain disorders, DNEs provide a better understanding of disease pathology, facilitate personalized risk assessment, and hold promise for targeted interventions and population selection.

## Methods

### Study populations

Our previous studies used semi-supervised AI models to define the nine DNEs from four disease case-control populations. These populations consisted of 865 healthy controls (CN), 1096 individuals with mild cognitive impairment (MCI), and 414 AD patients from ADNI^21^ for AD1 and AD2, 362 typically developing controls, and 307 patients with autism spectrum disorder (ASD) from ABIDE^22^ for ASD1-3, 495 healthy controls and 501 LLD patients from the LLD study^2^ for LLD1-2, and 364 healthy controls and 307 SCZ patients from PHENOM^23^ for SCZ1-2. For more detailed information about the characteristics of the study populations, please refer to the original papers.

The trained AI models were then applied to the UKBB general population as independent data. UKBB is a population-based study of approximately 500,000 people recruited from the United Kingdom between 2006 and 2010. The UKBB study has ethical approval, and the ethics committee is detailed here: https://www.ukbiobank.ac.uk/learn-more-about-uk-biobank/governance/ethics-advisory-committee. The protocols of this study were approved by the University of Pennsylvania institutional review board. The current study analyzed 39,178 multimodal brain MRI data from UKBB. T1-weighted MRI data were locally processed at the University of Pennsylvania; imaging-derived phenotypes (IDP) from diffusion and resting-state functional MRI were downloaded from UKBB. In addition, we processed the imputed genotype data^28^ from UKBB for GWAS analyses. Last, other clinical traits were also analyzed, including phenotypes related to nine human organ systems. The current work was performed under application numbers 35148 and 60698. We detail our dataset splits in **Supplementary Text 4**.

### Semi-supervised AI methods to derive the nine DNEs

The methodologies used in the current study to derive the nine DNEs belong to the semi-supervised learning algorithms (**Fig. 1a**) pioneered by our group. Refer to a review for details of this type of modeling. In particular, the current study employed the HYDRA^87^ and Surreal-GAN^14^ models.

#### (a) Surreal-GAN

Surreal-GAN^14^ dissects underlying disease-related heterogeneity via a deep representation learning approach, instead of the discriminative SVM, under the principle of semi-supervised clustering – the “*1-to-k*” mapping. The methodological advance of this method is that Surreal-GAN models disease heterogeneity as a continuous dimensional representation, enforces monotone disease severity in each dimension, and allows the non-exclusive manifestation of all dimensions in the same participant (**Supplementary Method 1a**).

#### (b) HYDRA

HYDRA leverages a widely used discriminative method, i.e., support vector machines (SVM), to seek the “*1-to-k*” mapping. The novelty is that HYDRA extends multiple linear SVMs to the non-linear case piecewise, thereby simultaneously serving for classification and clustering. Specifically, it constructs a convex polytope by combining the hyperplane from *k* linear SVMs, separating the CN group from the *k* subpopulation of the PT (patient) group. Intuitively, each face of the convex polytope can be regarded to encode each subtype, capturing a distinct disease effect (**Supplementary Method 1b**). **Supplementary Text 5** details the original image patterns of the diseased population in our previous study.

### Imaging analyses

#### (a) T1-weighted MRI processing

All images were first corrected for magnetic field intensity inhomogeneity.^88^ A deep learning-based skull stripping algorithm was applied to remove extra-cranial material. In total, 145 IDPs were generated in gray matter (GM, 119 ROIs), white matter (WM, 20 ROIs), and ventricles (6 ROIs) using a multi atlas label fusion method from the MUSE atlas.^89^ The ROIs were fit to the four machine learning models to derive the nine DNEs. The imaging quality check is detailed in **Supplementary Method 2**.

#### (b) Neuroanatomical pattern of the nine DNEs

We assessed the neuroanatomical patterns exhibited by the nine DNEs within the general population. Since the DNEs were defined based on the 119 GM ROIs obtained from T1-weighted MRI scans, we aimed to test whether these patterns observed in the disease populations were manifested in the general population. To this end, we used a linear regression model in which each DNE was treated as the dependent variable, while the ROI, age, age-squared, sex, age x sex interaction, age-squared x sex interaction, intracranial volume, brain positions in the scanner (lateral, transverse, and longitudinal; Field ID: 25756-25758), and head motion were considered independent variables and covariates.

#### (c) PWAS for the nine DNEs

We performed PWAS to associate the nine DNEs to each of the 611 additional phenotypes. PWAS excluded the 119 GM ROIs utilized to derive the nine DNEs to prevent any potential circular effects. Instead, the analysis incorporated IDPs from other modalities, such as diffusion and resting-state functional MRI. The same linear regression models and multiple comparison corrections were employed.

To check the robustness of our PWAS results, we also performed two sensitivity checks: *i*) sex-stratified PWAS for males and females, and *ii*) split-sample PWAS by randomly dividing the entire population into two splits (sex and age did not significantly differ).

### Genetic analyses

We used the imputed genotype data for all genetic analyses, and our quality check pipeline resulted in 31,929 participants with European ancestry and 6,477,810 SNPs. First, we excluded 36,394 related individuals (up to 2^nd^-degree) from the full UKBB sample (*N*∼500k) using the KING software for family relationship inference^90^. We then removed duplicated variants from all 22 autosomal chromosomes. Individuals whose genetically identified sex did not match their self-acknowledged sex were removed. Other excluding criteria were: i) individuals with more than 3% of missing genotypes; ii) variants with minor allele frequency (MAF) of less than 1%; iii) variants with larger than 3% missing genotyping rate; iv) variants that failed the Hardy-Weinberg test at 1×10^−10^. The Hardy-Weinberg test removes the SNPs that deviate from the expected HWE frequency, ensures the integrity of the genetic data, reduces false-positive findings, and improves the overall robustness of the GWAS results. To adjust for population stratification,^91^ we derived the first 40 genetic principle components (PC) using the FlashPCA software^92^. Details of the genetic quality check protocol are described elsewhere.

#### (a) GWAS

For GWAS, we ran a linear regression using Plink^93^ for each DNE, controlling for confounders of age, age-squared, sex, age x sex interaction, age-squared x sex interaction, the first 40 genetic principal components, total intracranial volume, three brain position parameters in the scanner, and head motion were included, as suggested by a previous study^30^. We adopted the genome-wide P-value threshold (5 × 10^−8^) and annotated independent genetic signals considering linkage disequilibrium (see below).

To check the robustness of our GWAS results, we also performed several sensitivity checks: *i*) fastGWA^39,40^ for a generalized mix effect model, *ii*) sex-stratified GWAS for males and females, *iii*) split-sample GWAS by randomly dividing the entire population into two splits (sex and age did not significantly differ), *iv*) comparison of the GWAS results using the 1348 participants (i.e., 1116 European ancestry) that were collected for baseline and longitudinal scans from UKBB, *v*) non-European GWAS (*N*=4783), *vi*) independent GWAS on ADNI whole-genome sequencing data (*N*=1555) on AD1 and AD2, *vii*) concordance with six European ancestry GWAS from PGC, including AD, ADHD, ASD, BIP, OCD, and SCZ (**Supplementary Table 3a**).

#### (b) SNP-based heritability

We estimated the SNP-based heritability (*h*^2^) using GCTA^38^ with the same covariates as in GWAS. GCTA estimates the SNP-based heritability using a method called restricted maximum likelihood (REML) to quantify the proportion of phenotypic variance in a trait that the additive effects of all common SNPs can explain, which was claimed to address the “missing heritability”. The main steps involved in GCTA include constructing the genetic relationship matrix, modeling phenotypic variance, and using REML to estimate the *h*^2^.

#### (c) Annotation of genomic loci

The annotation of genomic loci and mapped genes was performed via FUMA^94^. For the annotation of genomic loci, FUMA first defined lead SNPs (correlation *r*^2^ ≤ 0.1, distance < 250 kilobases) and assigned them to a genomic locus (non-overlapping); the lead SNP with the lowest P-value (i.e., the top lead SNP) was used to represent the genomic locus. For gene mappings, three different strategies were considered. First, positional mapping assigns the SNP to its physically nearby genes (a 10 kb window by default). Second, eQTL mapping annotates SNPs to genes based on eQTL associations using the GTEx v8 data^95^. Finally, chromatin interaction mapping annotates SNPs to genes when there is a significant chromatin interaction between the disease-associated regions and nearby or distant genes^94^. The definition of top lead SNP, lead SNP, independent significant SNP, and candidate SNP can be found in **Supplementary Method 3**.

To determine if a genomic locus is new (i.e., newly discovered in our study), we used FUMA to query the candidate SNPs, independent significant SNPs, and top lead SNP in the EMBL-EBI GWAS Catalog, taking LD into account. For these new loci, we further manually queried the non-significant SNPs in LD (*r*^2^>0.1) with these SNPs directly on the GWAS Catalog platform on 29^th^ October 2024. A locus is considered new if no previous GWAS has identified associations with any of these SNPs. It is worth noting that the current definition of new loci is not completely exhaustive and objective, as new GWAS data are being collated into the GWAS Catalog, while other public data platforms (such as the GWAS Catalog and the IEU Open GWAS Project) are not considered.

#### (d) Phenome-wide association queries for the identified loci in the GWAS Catalog

We queried the candidate and significant independent SNPs, considering LD (**Supplementary Method 3**), within each locus in the EMBL-EBI GWAS Catalog (query date: 2^nd^ June 2023 and also via FUMA version: v1.5.4) to determine their previously identified associations with any other traits. For these associated traits, we further mapped them into several high-level categories for visualization purposes. We categorized these traits based on their primary affected organ systems, with examples including blood pressure under cardiovascular, brain MRI-derived measures under brain, and AD under neurodegenerative disorders. Other platforms, such as the GWAS Atlas (https://atlas.ctglab.nl/PheWAS), used a similar categorization approach; we followed a similar approach to curating the high-level categories for the trait-category mapping.

#### (e) Genetic correlation

We used the LDSC^37^ software to estimate the pairwise genetic correlation (*g_c_*) between each pair of DNEs, as well as between the nine DNEs and 9 BAGs of multiple organ systems from our previous work^17^ and 6 neurodegenerative and neuropsychiatric disorders from PGC (**Supplementary Table 3a**). We used the precomputed LD scores from the 1000 Genomes of European ancestry. To ensure the suitability of the GWAS summary statistics, we first checked that the selected study’s population was European ancestry; we then guaranteed a moderate SNP-based heritability *h*^2^ estimate. Notably, LDSC corrects for sample overlap and provides an unbiased estimate of genetic correlation^96^. Benjamini-Hochberg procedure was performed to account for multiple comparisons.

#### (f) Bayesian colocalization

We used the R package (*coloc*) to investigate the genetic colocalization signals between two traits at each genomic locus. We employed the Fully Bayesian colocalization analysis using Bayes Factors (*coloc.abf*). This method examines the posterior probability (PP.H4.ABF: Approximate Bayes Factor) to evaluate hypothesis *H4*, which suggests the presence of a single shared causal variant associated with both traits within a specific genomic locus. To determine the significance of the *H4* hypothesis, we set a threshold of PP.H4.ABF>0.8^51^. All other parameters (e.g., the prior probability of p_12_) were set as default. We also performed relevant sensitivity analyses to check the robustness of our findings. For each pair of traits, the genomic locus (*N*>100 SNPs) was defined by default from FUMA on the nine DNEs, and then the *coloc* package extracted and harmonized the GWAS summary statistics within this locus for the other trait.

#### (g) Two-sample bidirectional Mendelian randomization

We did not perform causal inference between each pair of DNEs due to the overlapped populations and low sample sizes in split-sample analyses.

We employed a bidirectional, two-sample Mendelian randomization using the TwoSampleMR package^63^ to infer the causal relationships between the nine DNEs and the eight BAGs across nine human organ systems^17^ (excluding the brain). The forward and inverse Mendelian randomization was performed between each trait pair by switching the exposure and outcome variables. We applied five different Mendelian randomization methods and reported the results of inverse variance weighted (IVW) in the main text and the four others (i.e., Egger, weighted median, simple mode, and weighted mode estimators) in the supplement.

We then performed Mendelian randomization between the nine DNEs and eleven chronic diseases spanning the whole-body system. These diseases include four diseases from PGC (the ASD and OCD GWAS summary statistics did not provide the allele frequency information) and seven diseases unbiasedly curated in our previous work^17^, which detailed a systematic procedure to choose the appropriate traits. After harmonizing their GWAS summary statistics (using the function *harmonise_data* from 2SampleMR), this resulted in 11 clinical traits with at least six valid IVs (i.e., SNPs). The clinical traits included in our Mendelian randomization are presented in **Supplementary Table 11**. Benjamini-Hochberg correction was performed for all tested traits.

We performed several sensitivity analyses. First, a heterogeneity test was performed to check for violating the IV assumptions. Horizontal pleiotropy was estimated to navigate the violation of the IV’s exclusivity assumption^97^ using a funnel plot, single-SNP Mendelian randomization approaches, and Mendelian randomization Egger estimator^98^. Moreover, the leave-one-out analysis excluded one instrument (SNP) at a time and assessed the sensitivity of the results to individual SNP.

#### (h) PRS calculation for the nine DNEs

We calculated the PRS using the GWAS results from the split-sample analyses. The weights of the PRS were defined based on split1 data (base data), and the split2 GWAS summary statistics were used as the target data for PRS calculation. The QC steps for the base data are as follows: *i*) removal of duplicated and ambiguous SNPs for the base data; *ii*) clumping the base GWAS data; *iii*) pruning to remove highly correlated SNPs in the target data; *iv*) removal of high heterozygosity samples in the target data; *v*) removal of duplicated, mismatching and ambiguous SNPs in the target data. After rigorous QC, we used PLINK to generate PRS for the split2 population by adopting the classic C+T method (clumping + thresholding). To determine the “best-fit” PRS, we performed a linear regression using the PRS calculated at different P-value thresholds (0.001, 0.05, 0.1, 0.2, 0.3, 0.4, 0.5), controlling for age, sex, intracellular volume, and the first forty genetic PCs. For each DNE-PRS, we chose the P-value threshold with the highest incremental *R*^2^ (**Supplementary Figure 19**). The comparison between PLINK and PRS-CS^99^ is presented in **Supplementary Text 6** and **Figures 20–22**.

### Disease, cognition, and mortality outcome prediction

We employed logistic regression to calculate the incremental R-squared (*R*^2^) statistics of the nine DNEs and PRSs to predict 14 disease categories (**a**), support vector machines to classify the healthy control participants from the disease groups (**b**), and Cox proportional hazard model to predict mortality outcomes (**c**). The patients for the 14 disease categories were defined based on the ICD-10 code from the UKBB website (Data field: 41270). The healthy control group included participants without any ICD-10-based disease diagnoses. The cognitive function data included 8 cogntive scores (Category: 100026), which were detailed in **Supplementary File 23**. The mortality outcome refers to the date of death: https://biobank.ndph.ox.ac.uk/ukb/field.cgi?id=40000.

#### (a) Pseudo R-squared (*R*^2^) statistics of the logistic regression

We built a null model by including age, sex, and intracranial volume as predictors and the disease as the outcome variable. The alternative model took the disease-specific DNE or PRS as one additional predictor. The incremental *R*^2^ was calculated as the difference between the pseudo *R*^2^ of the alternative model and that of the null model, implemented by the *PseudoR2* function from the *DescTools* R package (v 0.99.38). For the nine PRSs, we used the PRS target sample (*N*=15,891). For the nine DNEs, we calculated the incremental *R*^2^ using the entire UKBB sample (*N*=39,178) and the PRS target sample (*N*=15,891). We reported the incremental *R*^2^ of the DNE/PRS in the main manuscript (**Fig. 5a** and **b**). Other metrics, including *R*^2^ of the null and the alternative model, sample sizes, and the β values of other covariates, are presented in **Supplementary File 20-21**.

#### (b) Support vector machines to classify patients vs. controls

Using the PRS target sample (*N*=15,891), we used 10000 participants in a nested cross-validation (CV) procedure (i.e., CV training/validation/test datasets) to select the hyperparameter *C* in SVM. In addition, we held out 5581 participants as an independent test dataset. In **Fig. 5c**, we only presented the classification accuracy from the independent test dataset. The nested cross-validation (CV) procedure^100^ involved an outer loop repeated 50 times, where 80% of the data were randomly selected for training/validation and 20% for testing. Within each outer loop iteration, an inner loop used 80% of the training/validation data for a 10-fold training/validation split. Critically, the model trained on the training/validation/test datasets generalized to the independent test dataset (**Fig. 5c**). **Supplementary File 22** contains various metrics, including balanced accuracy, sensitivity, specificity, negative predictive value (NPV), positive predictive value (PPV), and sample sizes for the independent dataset (*Ind. accuracy*) and the CV training/validation/test datasets (*CV accuracy*). We did not statistically compare the performance between different machine learning models as this is a complex matter without a universal solution. A standard *t*-test on cross-validation results is too liberal and should not be applied, as shown by Nadeau and Bengio^101^. The corrected resampled *t*-test^101^ was proposed, but it depends on the data and the cross-validation setup.

#### (c) Support vector regression to predict cognitive scores

Using the PRS target sample (*N*=15,891), we used 10000 participants in a nested cross-validation (CV) procedure (i.e., CV training/validation/test datasets) to select the hyperparameter *C* in SVM. In addition, we held out 5581 participants (some had missing values depending on specific cognitive scores) as an independent test dataset (*Ind. r*). In **Fig. 5d**, we only presented Pearson’s *r* from the independent test dataset. The same nested CV procedure was used, as mentioned above. Likewise, **Supplementary File 23** encompasses a range of metrics, including mean absolute error, P-values, and sample sizes, pertaining to the independent dataset (*Ind. r)* and the CV training/validation/test datasets (*CV r*).

#### (d) Cox proportional hazard model to predict the date of death

To evaluate the predictive capacity of individual DNE and PRS for mortality risk, we employed a Cox proportional hazard model while adjusting for covariates such as age and sex. The hazard ratio (HR) was calculated and reported as the effect size measure that indicates the influence of each DNE or PRS on mortality risk. Furthermore, we incrementally added the most predictive DNE or PRS to the Cox model to determine when the model’s performance reached saturation. The concordance index (CI) was utilized to assess the model’s performance using a 5-fold cross-validation procedure. All survival analyses were conducted using the *lifelines* (v0.25.7) Python package.

## Data Availability

The GWAS summary statistics and pre-trained AI models from this study are publicly accessible via the MEDICINE Knowledge Portal (https://labs-laboratory.com/medicine/) and Synapse (https://www.synapse.org/Synapse:syn64923248/wiki/630992^102^). Genomic loci annotation used data from FUMA (https://fuma.ctglab.nl/). UKBB data can be requested at https://www.ukbiobank.ac.uk/. GWAS summary data from the Psychiatric Genomics Consortium (PGC) can be accessed at https://pgc.unc.edu/. MUSE atlas is generated via the pipeline at: https://github.com/CBICA/MUSE.

## Code Availability

The software and resources used in this study are all publicly available:

- *MLNI*^103^ *(HYDRA)*: https://github.com/anbai106/mlni, DNEs for ASD1-3, LLD1-2, SCZ1-2
- *Surreal-GAN*^104^: https://github.com/zhijian-yang/SurrealGAN, DNEs for AD1-2

## Acknowledgments

We want to express our sincere gratitude to the UK Biobank team for their invaluable contribution to advancing clinical research in our field. We thank the Psychiatric Genomics Consortium (PGC: https://pgc.unc.edu/) for generously sharing the GWAS summary statistics with the scientific community. This study used the UK Biobank resource under Application Numbers 35148 (CD) and 60698 (AZ). JW leads the MULTI consortium under UK Biobank Application number (647044). We also gratefully acknowledge the support of the iSTAGING consortium, funded by the National Institute on Aging through grant RF1 AG054409 at the University of Pennsylvania (CD). Additionally, we acknowledge the funding program from the Rebecca L. Cooper Foundation at the University of Melbourne (AZ). YET is supported by a National Health and Medical Research Council Investigator Grant (APP2026413)

## Author Contributions Statement

Dr. Wen has full access to all the data in the study and takes responsibility for the integrity of the data and the accuracy of the data analysis.

*Study concept and design*: Wen, Davatzikos

*Acquisition, analysis, or interpretation of data*: all authors

*Drafting of the manuscript*: Wen

*Critical revision of the manuscript for important intellectual content*: all authors

*Statistical analysis*: Wen

