## Supplementary materials for "Neuroimaging-AI endophenotypes reveal underlying mechanisms and genetic factors contributing to progression and development of four brain disorders"

**Method 1: The two semi-supervised AI methods used in the current study**
**Method 2: Image quality check for the ROIs of the MUSE atlas**
**Method 3: The definition of genomic loci, independent significant SNP, lead SNP, candidate SNP**
**Text 1: Statistical harmonization protocol in iSTAGING**
**Text 2: Sensitivity check for the main PWAS**
**Text 3: Sensitivity check for the main GWAS using the entire UKBB participants of European ancestry**
**Text 4: Dataset splits for training our PRS and machine leanring in this study**
**Text 5: Imaging patterns of the 9 DNEs in the original studies**
**Text 6: Comparison for PRS generated by PLINK and PRS-CS**
**Figure 1: The sensitivity analyses of AD1 GWAS**
**Figure 2: The sensitivity analyses of AD2 GWAS**
**Figure 3: The sensitivity analyses of ASD1 GWAS**
**Figure 4: The sensitivity analyses of ASD2 GWAS**
**Figure 5: The sensitivity analyses of ASD3 GWAS**
**Figure 6: The sensitivity analyses of LLD1 GWAS**
**Figure 7: The sensitivity analyses of LLD2 GWAS**
**Figure 8: The sensitivity analyses of SCZ1 GWAS**
**Figure 9: The sensitivity analyses of SCZ2 GWAS**
**Figure 10: Exemplary genomic locus for each DNE**
**Figure 11: Phenome-wide associations of the causal SNP (rs9257566) in the GWAS Catalog and Ensembl**
**Figure 12: Sensitivity check on the prior probability of *p12* for Genetic colocalization analyses**
**Figure 13: Genetic colocalization results between the nine DNEs, nine BAGs, and six neurodegenerative and neuropsychiatric disorders from PGC**
**Figure 14: Mendelian randomization sensitivity check for the brain BAG on LLD1**
**Figure 15: Mendelian randomization sensitivity check for the cardiovascular BAG on ASD3**
**Figure 16: Mendelian randomization sensitivity check for the pulmonary BAG on LLD2**
**Figure 17: Mendelian randomization sensitivity check for AD2 on AD**
**Figure 18: Incremental $R^2$ of the nine DNEs to predict the 14 disease categories using only the PRS target population**
**Figure 19: Incremental $R^2$ of the nine PRSs derived from PLINK to explain the nine DNEs in the PRS target population**
**Figure 20: Incremental $R^2$ of the nine PRSs derived from PRS-CS to explain the nine DNEs in the PRS target population**
**Figure 21: Scatter plot for the nine PRSs derived from PLINK and PRS-CS**
**Figure 22: Incremental $R^2$ of the nine DNEs to predict the 14 disease categories using the nine PRSs derived from PRS-CS for only the PRS target population**
**Figure 23: Model considerations and illustrations of the semi-supervised clustering approaches**
**Figure 24: Comparisons of beta coefficient and P-value between PLINK and fastGWA**

**Table 1: Descriptive statistics of the difference between the healthy control and disease groups for the 9 DNEs in the UKBB population.**
**Table 2: The SNP-based heritability estimates**
**Table 3: The six case-control GWAS on neurodegenerative and neuropsychiatric disorders from from PGC (a) and four lifestyle factors and cognitive scores (b).**
**Table 4: Genetic correlation estimates between the nine DNEs**
**Table 5: Genetic correlation estimates between the nine DNEs and the nine BAGs**
**Table 6: Table 6: Genetic correlation estimates between the nine DNEs and the six neurodegenerative and neuropsychiatric disorders from PGC (a) and four lifestyle factors and cognitive scores (b)**.
**Table 7: Incremental $R^2$ of the nine DNEs to predict the 14 disease categories**
**Table 8: Incremental $R^2$ of the nine PRSs to predict the 14 disease categories**
**Table 9: Prediction accuracy of the nine DNEs and PRSs to predict the 14 disease categories (a) and 8 cognitive scores (b)**.
**Table 10: Survival analysis results of the nine DNEs and PRSs to predict the risk of mortality**
**Table 11: The 9 BAG GWAS and 11 chronic diseases GWAS used in our Mendelian randomization analyses**

**Method 1: The two semi-supervised AI methods used in the current study**

**a) Surreal-GAN**:

Surreal-GAN[1] is a novel deep representation learning method. Unlike the other semi-supervised methods which seek a categorical disease subtype, it dissects the neuroanatomical heterogeneity of brain diseases into $k$ continuous variables. Compared to its precursor, the Smile-GAN model,[2] Surreal-GAN follows the same principle of semi-supervised clustering but solves several limitations of Smile-GAN. Semi-supervised clustering methods seek the so-called "*1-to-k*" mapping by learning distribution transformation from CN data to PT data. The following schematic figure demonstrates the principles of Smile-GAN and Surreal-GAN in the semi-supervised learning framework to disentangle AD neuroanatomical heterogeneity (**Supplementary Figure 23**).

Surreal-GAN learns one transformation function $f$, which transforms CN data $\boldsymbol{x}$ to different synthesized PT data $\boldsymbol{y}' = f(\boldsymbol{x}, \boldsymbol{z})$, with latent variable $\boldsymbol{z}$ specifying distinct mapping directions. However, compared to Smile-GAN, Surreal-GAN considers that disease heterogeneity spatially and temporally (subtype) expands along a continuum (severity), similar to Sustain[3] to a certain extent. Thus, the latent variable $\boldsymbol{z}$ is modeled as a continuous variable, allowing infinite mapping directions from CN to PT data. $\boldsymbol{z}{\sim}p_{lat}(\boldsymbol{z})$ is sampled from a multivariate uniform distribution $U(0,1)^k$, rather than a categorical distribution as in Smile-GAN. Further, Surreal-GAN aims at disentangling spatial and temporal variations independently so that each dimension of the latent variable $\boldsymbol{z}$ is correlated with the severity of one relatively homogeneous imaging pattern. In contrast, different dimensions can be associated with spatially different imaging patterns.

To achieve the goals above, the objective function of Surreal-GAN consists of one adversarial loss and other regularization terms. The adversarial loss aims at matching the distribution of synthesized PT data, $p_{syn}$, and the distribution of real PT data, $p_{PT}$. Other regularization terms serve the following purposes: 1) encouraging sparse transformations (change loss); 2) reconstructing latent variables from synthesized or real PT data through the decomposer $g_1$ (decomposition loss) and reconstruction function $g_2$ (reconstruction loss); 3) boosting spatial separation of synthesized/captured patterns (orthogonality loss); 4) enforcing positive correlations between components of $\boldsymbol{z}$ and severity of synthesized patterns (monotonicity loss and cn loss).

Specifically, with distributions of CN, real PT, synthesized PT data denoted as $p_{CU}(\boldsymbol{x})$, $p_{PT}(\boldsymbol{y})$ and $p_{syn}(\boldsymbol{y}')$ respectively, the adversarial loss is defined as:

$$L_{GAN}(D, f) = E_{\boldsymbol{y}\sim p_{PT}(\boldsymbol{y})}\left[log\left(D(\boldsymbol{y})\right)\right] + E_{\boldsymbol{z}\sim p_{Lat}(\boldsymbol{z}), \boldsymbol{x}\sim p_{CN}(\boldsymbol{x})}\left[1 - log\left(D\left(f(\boldsymbol{x}, \boldsymbol{z})\right)\right)\right]$$

$$= E_{\boldsymbol{y}\sim p_{PT}(\boldsymbol{y})}\left[log\left(D(\boldsymbol{y})\right)\right] + E_{p_{syn}(\boldsymbol{y}')}\left[1 - log\left(D(\boldsymbol{y}')\right)\right]$$

The transformation function attempts to synthesize PT data $\boldsymbol{y}'$, so that they follow similar distributions as real PT data. The discriminator, $D$, tries to distinguish the synthesized PT data from real PT data. Therefore, the discriminator is updated to maximize the adversarial loss, while the transformation function is optimized to minimize it.

Other regularization terms are introduced to further regularize the transformation function $f$. With the assumption that the disease process will not change brain anatomy dramatically and primarily only affect certain regions throughout most of the disease stages, the change loss is introduced to control sparsity and distance of transformations:

$$L_{change}(f) = E_{\boldsymbol{z}\sim p_{Lat}(\boldsymbol{z}), \boldsymbol{x}\sim p_{CU}(\boldsymbol{x})}\left[\|f(\boldsymbol{x}, \boldsymbol{z}) - \boldsymbol{x}\|_1\right]$$

The decomposer $g_1$ serves to reconstruct changes synthesized by each component: $\boldsymbol{q}_i = f(\boldsymbol{x}, \boldsymbol{a}^i) - \boldsymbol{x}$, where $\boldsymbol{a}_i^i = \boldsymbol{z}_i$ and $\boldsymbol{a}_j^i = 0$ for $j \neq i$. With $\widehat{\boldsymbol{q}}_{f(x,z)} = [\boldsymbol{q}_1^T, \boldsymbol{q}_2^T, \cdots, \boldsymbol{q}_M^T]^T$, the decomposition loss is defined as:

$$L_{decom}(f, g_1) = E_{\boldsymbol{z}\sim p_{Lat}(\boldsymbol{z}), \boldsymbol{x}\sim p_{CN}(\boldsymbol{x})}\left[\left\|g_1(f(\boldsymbol{x}, \boldsymbol{z})) - \widehat{\boldsymbol{q}}_{f(\boldsymbol{x},\boldsymbol{z})}\right\|_2\right]$$

The reconstruction function $g_2$ serves to further reconstruct each component of the sampled $\boldsymbol{z}$ variable from $g_1\big(f(\boldsymbol{x}, \boldsymbol{z})\big)$. The function $g$ is defined as a composition of $g_1$ and $g_2$, with $g\big(f(\boldsymbol{x}, \boldsymbol{z})\big) = \left[g_2\left(g_1\big(f(\boldsymbol{x}, \boldsymbol{z})\big)_{0:S}\right), \cdots, g_2\left(g_1\big(f(\boldsymbol{x}, \boldsymbol{z})\big)_{S*(M-1):S*M}\right)\right]^T$ (S = number of ROIs), and the reconstruction loss is defined as

$$L_{recons}(f, g) = L_{recons}(f, g_1, g_2) = E_{\boldsymbol{z}\sim p_{Lat}(\boldsymbol{z}), \boldsymbol{x}\sim p_{CN}(\boldsymbol{x})}\left[\left\|g\big(f(\boldsymbol{x}, \boldsymbol{z})\big) - \boldsymbol{z}\right\|_2\right]$$

The orthogonality loss aims to boost changes led each component, $\boldsymbol{q}_i$, to be relatively orthogonal to each other. For this purpose, a matrix $\boldsymbol{A}_{f(\boldsymbol{x},\boldsymbol{z})}$ is constructed with the $i_{th}$ column $A_{f(\boldsymbol{x},\boldsymbol{z})_{:,i}} = \boldsymbol{q}_i/||\boldsymbol{q}_i||_2$, and the orthogonality loss is defined as:

$$L_{ortho}(f) = E_{\boldsymbol{z}\sim p_{Lat}(z), \boldsymbol{x}\sim p_{CU}(\boldsymbol{x})}[||\boldsymbol{A}_{f(\boldsymbol{x},\boldsymbol{z})}^T \boldsymbol{A}_{f(\boldsymbol{x},\boldsymbol{z})} - I||_F]$$

To encourage a positive correlation between the severity of the synthesized pattern and the value of each component $\boldsymbol{z}_i$, another latent variable $\boldsymbol{z}'\sim p_{sev}(\boldsymbol{z}'|\boldsymbol{z})$ is sampled conditioned on previously sampled $\boldsymbol{z}$ variables, such that $\boldsymbol{z}_i' \geq \boldsymbol{z}_i$ for any $1 \leq i \leq k$. With these two sampled latent variables, the monotonicity loss is defined as:

$$L_{mono}(f) = E_{\boldsymbol{z}\sim p_{Lat}(\boldsymbol{z}), \boldsymbol{z}\sim p_{sev}(\boldsymbol{z}'|\boldsymbol{z}), \boldsymbol{x}\sim p_{CU}(\boldsymbol{x})}[||max\ (|f(\boldsymbol{x}, \boldsymbol{z}) - \boldsymbol{x}| - |f(\boldsymbol{x}, \boldsymbol{z}') - \boldsymbol{x}|, 0)||_2]$$

The cn loss is introduced to further ensure that a small $\boldsymbol{z}$ variable lead to mild patterns. By letting $p_{cn}(\boldsymbol{z}) = U(0, 0.05)^k$ to be a multivariate uniform distribution, the cn loss is defined as:

$$L_{cn}(f) = E_{\boldsymbol{z}^{cn}\sim p_{cn}(\boldsymbol{z}), \boldsymbol{x}\sim p_{CU}(\boldsymbol{x})}[\|f(\boldsymbol{x}, \boldsymbol{z}^{cn}) - \boldsymbol{x}\|_1]$$

With all loss functions introduced above, the full objective of Surreal-GAN can be written as

$$\begin{aligned} L(D, f, g_1, g_2) = L_{GAN}(D, f) + \gamma L_{change}(f) + \kappa L_{decom}(f, g_1) + \zeta L_{recon}(f, g_1, g_2) \\ + \lambda L_{ortho}(f) + \mu L_{mono}(f) + \eta L_{cn}(f) \end{aligned}$$

With $\gamma$, $\kappa$, $\zeta$, $\lambda$, $\mu$, and $\eta$ being hyperparameters that control the relative importance of each loss function during the training process. More details of parameter selections can be found in the Surreal-GAN paper.[1]

Through the training process, parametrized functions $f$ , $g_1$, and $g_2$ are updated to satisfy that:

$$f, g_1, g_2 = arg \min_{f,g} \max_{D} L(D, f, g_1, g_2)$$

More importantly, after the training process, the function g, a composition of $g_1$ and $g_2$, can be applied to unseen PT data to infer the latent variable, which is referred to as the R-indices (i.e., dimensions) of PT data.

**b) HYDRA**: HYDRA leverages multiple ($k$) support vector machines (SVM) to seek the "*1-to-k*" mapping. It extends the $k$ linear SVMs' hyperplanes to the non-linear case piecewise, thereby constructing a $k$-face polytope for classification and clustering. This polytope, therefore, separates the CN group and the $k$ subpopulation of the PT group. Intuitively, each face of the convex polytope can be regarded to encode each subtype to derive each DNE – the distance of each participant to his/her nearest face of the $k$-face polytope (**Fig. 1a**).

To solve this joint optimization problem, the convex polytope is estimated by sequentially solving each linear SVM as a sub-problem under the principle of the sample-weighted SVM. The optimization stops until the sample weights get stable, i.e., the polytope was stably established. The objective of maximizing the polytope's margin can be summarized as:

$$\min_{\{w_j,b_j\}_{(j=1)}^{k}} \sum_{j=1}^{k} \frac{\left\|w_j\right\|_2^2}{2} + \mu \sum_{i|y_i=+1} \frac{1}{k} \max\{0, 1 - w_j^T X_j^T - b_j\}$$

$$+ \mu \sum_{i|y_i=-1} S_{i,j} \max\{0, 1 + w_j^T X_j^T + b_j\}$$

where $w_j$ and $b_j$ are the weight and bias for each hyperplane, respectively. $\mu$ is a penalty parameter on the training error, and $\boldsymbol{S}$ is the subtype membership matrix of dimension $n * k$ deciding whether a patient sample $i$ belongs to subtype $j$. The DNE indices are derived as:

$$DNE = w_j^T X_j^T + b_j$$

**Method 2: Image quality check for the ROIs of the MUSE atlas**

T1-weighted MRIs were first quality checked (QC) for motion, image artifacts, or restricted field-of-view. Another QC was performed as follows: First, the images were examined manually to evaluate for pipeline failures (e.g., poor brain extraction, tissue segmentation, and registration errors via https://cbica.github.io/MRISnapshot/). Furthermore, a second step automatically flagged images based on outlying values of quantified metrics; those flagged images were re-evaluated. In general, the image scan quality from the UKBB surpassed that of other ISTAGING studies due to more stringent acquisition protocols involving a single scanner type and scanning procedure, alongside rigorous data verification and exclusion criteria. Dr. Guray Erus conducted the quality check for the T1-weighted MRI scans, and we only excluded 2 scans during this quality check procedure within UKBB.

Following the QC procedure, Dr. Junhao Wen, Dr. Gyujoon Hwang, and Mr. Zhijian Yang individually applied the trained Surreal-GAN and HYDRA models to the UKBB participants. Eventually, Dr. Junhao Wen consolidated and overlapped the populations across the nine DNEs, resulting in 39,178 UKBB individuals being included in the current study.

**Method 3: The definition of genomic loci, independent significant SNP, lead SNP, candidate SNP**

FUMA defined the significant independent SNPs, lead SNPs, candidate SNPs, and genomic risk loci as follows (https://fuma.ctglab.nl/tutorial#snp2gene):

*Independent significant SNPs*

They are defined as SNPs with $P \leq 5\times10^{-8}$ that are independent of each other at the user-defined $r^2$ (set to 0.6 in the current study). We further describe *candidate SNPs* as those in linkage disequilibrium (LD) with independent significant SNPs. FUMA then queries each candidate SNP in the GWAS Catalog to check whether any clinical traits have been reported to be associated with previous GWAS studies.

*Lead SNPs*

Lead SNPs are defined as independent significant SNPs that are also independent of each other at $r^2<0.1$. If multiple independent significant SNPs are correlated at $r^2\geq0.1$, then the one with the lowest individual *P*-value becomes the lead SNP. If $r^2$ threshold is set to 0.1 for the independent significant SNPs, then they would constitute the identical set as the lead SNPs by definition. FUMA thus advises setting $r^2$ to be 0.6 or higher.

*Genomic risk loci*

FUMA defines genomic risk loci to include all independent signals physically close or overlapping in a single locus. First, independent significant SNPs dependent on each other at $r^2\geq0.1$ are assigned to the same genomic risk locus. Then, independent significant SNPs with less than the user-defined distance (250 kilobases by default) away from one another are merged into the same genomic risk locus - the distance between two LD blocks of two independent significant SNPs is the distance between the closest points from each LD block. Each locus is represented by the SNP within the locus with the lowest *P*-value.

In our Phenome-wide look-up analysis, we used FUMA and also manually operated (to define whether a locus is new) to query all SNPs in LD with any significant SNPs with $r^2$ greater or equal to the threshold (0.1), as indicated in the original FUMA paper[4]: "In addition, independent significant SNPs and correlated SNPs are also linked to the GWAS catalog to provide insight into previously reported associations of the SNPs in the risk loci with a variety of phenotypes."

**Text 1: Statistical harmonization protocol in iSTAGING**

Leveraging the large-scale MRI data (>50,000) from the iSTAGING consortium, we statistically harmonized the MRI data in the current study using our original publication in 2020 for the ComBat-GAM method[5]. The method was encapsulated in an open-source software package at https://github.com/rpomponio/neuroHarmonize. We detail the harmonization protocol here for the low-dimensional imaging features (e.g., MUSE ROIs) using cross-sectional data in iSTAGING: *i*) we preprocessed all T1-weighted MRI data consolidated by the iSTAGING consortium that covers the entire lifespan using the RAVENS and MUSE pipeline (https://www.nitrc.org/projects/cbica_muse/); *ii*) we then defined the healthy control (CN) population in iSTAGING for these participants without any pathological diagnosis (e.g., AD and schizophrenia); *iii*) we ensured to reserve biological effects as covariates (e.g., age and sex) and alleviate site or batch effects (assign SITE as a numerical dummy variable for each study); *iv*) we applied the method to the data using the *harmonizationLearn* function, where we can specify the reference site and the age boundaries; *v*) we also provided the function *harmonizationApply* to harmonize out-of-sample data using the pre-trained model. All these procedures are also publicly available at our online NiChart platform: https://neuroimagingchart.com/components/#Harmonization.

**Text 2: Sensitivity check for the main PWAS**

**Split-sample PWAS**

In the PWAS from the first split, we found 184, 103, 191, 158, 147, 112, 120, 186, and 172 DNE-phenotype associations for the nine abovementioned DNEs (P-value < 0.05/611). Of these, 176 (162), 100 (83), 190 (176), 157 (135), 141 (127), 110 (92), 119 (117), 183 (171), and 170 (165) associations were replicated in the second split PWAS using both the nominal P-value (<0.05, 98.03% concordance rate) and the Bonferroni-corrected threshold (<0.05/611, 89.43% concordance rate). Detailed results are presented in **Supplementary eFile 4.**

**sex-stratified PWAS**

For the female-specific PWAS, we observed 178, 119, 180, 168, 135, 113, 149, 197, and 175 DNE- phenotype associations for the nine DNEs. Among these, 173 (154), 106 (75), 177 (164), 152 (127), 124 (107), 101 (81), 135 (122), 188 (161), and 173 (164) associations were replicated in the male-specific GWAS using both the nominal P-value (<0.05, 93.98% concordance rate) and the Bonferroni-corrected threshold (<0.05/611, 81.68% concordance rate). Detailed results are presented in **Supplementary eFile 5**.

### Text 3: Sensitivity check for the main GWAS using the entire UKBB participants of European ancestry

#### fastGWA for linear mixed model

In the GWAS using PLINK linear models, we found 287, 465, 233, 962, 88, 3, 3, 23, and 31 DNE-SNP associations for the nine abovementioned DNEs (P-value < $5x10^{-8}$). As expected, for fastGWA *i*) with related individuals (*N*~33k) and *ii*) without related individuals (*N*~32k), all the signals were replicated and had the same effect size directions (beta coefficient sign) at both the nominal P-value (<0.05) and the Bonferroni-corrected threshold (<0.05/*N*).

We then compared the *beta* coefficient and P-value for the three sets of GWAS: *i*) PLINK (*N*~32k), *ii*) fastGWA with related (*N*~33k), and *iii*) fastGWA without unrelated (a.k.a. to a linear model) (*N*~32k). For the -$\log_{10}$(P-value), fastGWA with related individuals, as expected, showed more significant signals compared to the other two GWASs. No statistical significance was observed regarding the effect size *beta* coefficients (**Supplementary Figure 24**). Detailed results are presented in **Supplementary eFile 7**.

#### Split-sample GWAS

In the GWAS from the first split, we found 5, 63, 87, 81, 9, 0, 0, 0, and 0 DNE-SNP associations for the nine abovementioned DNEs (P-value < $5x10^{-8}$). Of these, 5 (5), 63 (62), 87 (87), 0 (0), 0 (0), 0 (0), 0 (0), 0 (0), and 0 (0) associations were replicated in the second split GWAS using both the nominal P-value (<0.05, 63.26% concordance rate) and the Bonferroni-corrected threshold (<0.05/*N*, 63.26% concordance rate). Detailed results are presented in **Supplementary eFile 8**.

#### sex-stratified GWAS

For the female-specific GWAS, we observed 12, 180, 112, 83, 1, 0, 0, 0, and 1 DNE-SNP associations for the nine DNEs. Among these, 12 (4), 173 (173), 112 (90), 62 (54), 1 (1), 0 (0), 0 (0), 0 (0), and 0 (0) associations were replicated in the male-specific GWAS using both the nominal P-value (<0.05, 92.54% concordance rate) and the Bonferroni-corrected threshold (<0.05/*N*, 82.78% concordance rate). While we observe fewer significant loci (P-value < $5x10^{-8}$) in the sensitivity analyses than the results obtained from the full sample sizes, which may be due to reduced sample sizes, the signal peaks of the P-values remain consistent, as well as the effect directions. Detailed results are presented in **Supplementary eFile 9**.

#### Longitudinal GWAS

We examined the concordance rate of the GWAS by analyzing both the baseline data and longitudinal data from 1116 participants of European ancestry. These participants underwent a second MRI scan approximately two years after their initial scan. We found 16, 35, 32, 12, 20, 33, 18, 7, and 11 DNE-SNP associations in the baseline GWAS for the nine abovementioned DNEs (P-value < $5x10^{-6}$ due to the smaller sample size). Of these, the longitudinal GWAS can 100% replicate these associations at the nominal P-value (<0.05) and the Bonferroni-corrected threshold (<0.05/*N*). Detailed results are presented in **Supplementary eFile 10**.

#### Non-European ancestry GWAS

We assessed the concordance rate for the main GWAS findings in 4783 participants with non-European ancestries. In the main GWAS with European ancestry, we found 287, 455, 233, 962,

88, 3, 3, 23, and 31 DNE-SNP associations for the nine abovementioned DNEs (P-value < $5 \times 10^{-8}$). Of these, 70 (28), 263 (1), 204 (0), 363 (1), 39 (5), 0 (0), 1 (0), 10 (0), and 11(1) associations were replicated in non-European GWAS using both the nominal P-value (<0.05, 46.09% concordance rate) and the Bonferroni-corrected threshold (<0.05/*N*, 1.7% concordance rate). Detailed results are presented in **Supplementary eFile 11.**

**ADNI WGS GWAS**

In the independent ADNI whole-genome sequencing (WGS) data curated by the NIAGADS program, we observed low concordance rates for the AD1 and AD2 dimensions. Specifically, only 4 out of the 241 significant SNPs (P-value<$5 \times 10^{-8}$) for AD1 (1.51% concordance rate) and 28 out of the 379 significant SNPs (P-value<$5 \times 10^{-8}$) for AD2 (6.71% concordance rate) were replicated at the nominal significant level (P-value<0.05). These findings were expected due to the small sample size of ADNI (*N*=1555) and the differences between the curated research data for AD and the general population, which may impact the generalizability of the results. Detailed results are presented in **Supplementary eFile 12.**

**PGC GWAS**

In addition, we compared our DNE GWAS to 6 case-control GWAS (European ancestry) of Alzheimer's disease[6] (AD), attention deficit hyperactivity disorder[7] (ADHD), autism spectrum disorder[8] (ASD), bipolar disorder[9] (BIP, type I and type II combined), obsessive-compulsive disorder[10] (OCD), and schizophrenia[11] (SCZ) from the psychiatric genetic consortium (PGC) (**Supplementary Table 2**). After harmonizing the PGC summary statistics with our GWAS, we observed relatively low concordance rates of 10%, 14.58%, 10%, 10.31%, 14.81%, 0%, 0%, 11.11%, and 8.33% at the nominal significance threshold for the abovementioned nine DNEs (only for the top lead SNP, **Supplementary eFile 13**). These findings highlight the potential limitations of previous case-control GWAS in capturing the disease heterogeneity present in complex brain disorders. However, our DNE GWAS showed concordance to a certain extent. For instance, the association between the top lead SNP rs4858811 at 3p21.31 (P-value=$7.49 \times 10^{-9}$, $\beta$=0.046±0.008) and AD1 was marginally significant in the PGC AD case-control GWAS (P-value=0.014, $\beta$=0.024±0.01). Further evidence supporting this concordance can be found in the Bayesian colocalization analyses (**Fig. 4f**). Our approach incorporating DNEs provides valuable insights into disease heterogeneity that traditional case-control GWAS may have overlooked.

**Text 4: Dataset splits for training our PRS and machine leanring in this study**

To unbiasedly evaluate the PRS and machine learning models, we defined the following populations:

- *Disease case-control populations*: The four datasets used to train the AI models and define the nine DNEs from four brain diseases.
- *Independent UKBB general population (N*=39,178*)*: The UKBB population in which the trained AI models were applied to derive the nine DNEs.
- *PRS base/target population (split1/split2) (N*=15,968*)*: the UKBB population was divided into two splits (split1 vs. split2) in the split-sample GWAS. To derive the PRS, we used the GWAS from split1 as the base data and split2 as the target data. All disease and mortality prediction tasks involving PRS used only the PRS target population (*N*=15,968).

Within the UKBB general population, individuals with various diseases are classified under the ICD-10 code (Data field: 41270), though often in smaller numbers. Among the 39,178 participants from the UK Biobank included in our study, we outline the sample sizes and criteria for the healthy control group, as well as those with AD, ASD, LLD (encompassing a broad spectrum of depression), and SCZ, all categorized according to the ICD-10 code:

- *Healthy control*: 6390 participants without any ICD-10-based disease diagnoses from the data field: 41270 from the training/validation/test dataset.
- *AD*: 23 AD patients based on the ICD-10 code (G30).
- *ASD*: 6 ASD patients based on the ICD-10 code (F84.0).
- *LLD*: 1329 patients based on the ICD-10 code (F32).
- *SCZ*: 23 SCZ patients based on the ICD-10 code (F20).
- *The 14 systemic disease categories*: we used the ICD-10 code to define the disease groups. The exact ICD-10 code for each category is detailed in **Fig. 5a**.

**Text 5: Imaging patterns of the 9 DNEs in the original studies**

We summarized the main characteristics of the ASD1-3, LLD1-2, and SCZ1-2 derived by HYDRA in the disease-specific populations. ASD1 showed correlations with reduced brain volume, decreased cognitive function, and genetic variants related to aging. ASD2 exhibited enlarged subcortical volumes and was linked to the use of antipsychotic medication. ASD3 was identified by expanded cortical volumes and higher performance in nonverbal cognitive tasks[12]. LLD1 displayed relatively maintained brain structure without disruptions in white matter compared to healthy control individuals, whereas LLD2 exhibited extensive brain atrophy, disruptions in white matter integrity, cognitive decline, and increased severity of depression[13]. SCZ1 exhibited widespread reduction in grey matter volumes, particularly notable in the thalamus, nucleus accumbens, medial temporal, medial prefrontal/frontal, and insular cortices. SCZ2 displayed increased volume in the basal ganglia and internal capsule, while other brain volumes remained relatively normal[14]. We encapsulated the characteristics of the AD1 and AD2 derived by Surreal-GAN from the disease-specific population. AD1 represents a "diffuse-AD" dimension characterized by widespread brain atrophy, whereas AD2, described as a "medial temporal lobe-AD" dimension, exhibits focal atrophy in the medial temporal lobe (MTL)[15].

**Text 6: Comparison for PRS generated by PLINK and PRS-CS**

We also tested the PRS-CS[16] method to derive the nine PRS, which infers posterior SNP effect sizes under continuous shrinkage priors using GWAS summary statistics and an LD reference panel (i.e., UKBB reference). Compared to the PLINK method, PRS-CS obtained higher incremental $R^2$ to explain the 9 DNEs in the split2 data (1.99% vs. 0.52%, P-value=0.003 for two-sample t-test, **Supplementary Figure 20**; P-value thresholding was explicitly employed for PRS-CS to ensure a fair comparison). Second, the 9 PRSs from PLINK and PRC-CS were highly concordant and correlated with each other (Pearson's *r* correlation=0.98, P-value=0.000007, **Supplementary Figure 21**). Last but not least, the 9 PRSs from PRS-CS did not predict well for mental and behavioral disorders (ICD-10 code: F group) and diseases linked to the central nervous system (ICD-10 code: G group). That is, the highest incremental $R^2$ was obtained for ear diseases (H6-H9) using all combined 9 PRSs (**Supplementary Figure 22**). Therefore, we chose to use the 9 PRSs derived from PLINK for all results presented in **Fig. 5**.

**Figure 1: The sensitivity analyses of AD1 GWAS**

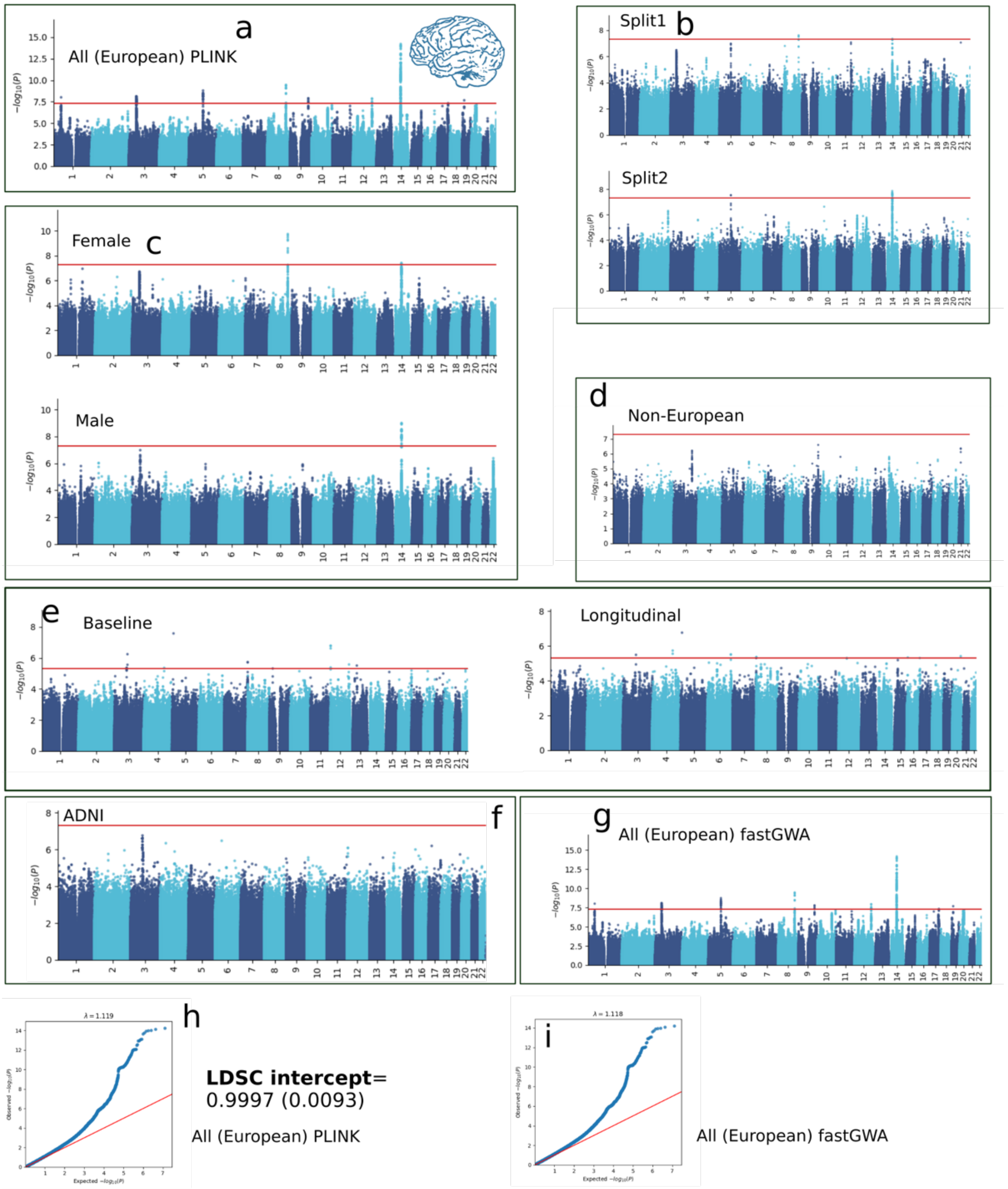


**a-g**) Manhattan plots for AD1 in all European ancestry (PLINK linear models), split1, split2, females, males, non-European ancestries, baseline and longitudinal data, independent ADNI WGS data, and European ancestry (fastGWA). **h-i**) the QQ plot, lambda, and LDSC intercept estimate for PLINK linear models and fastGWA.

**Figure 2: The sensitivity analyses of AD2 GWAS**

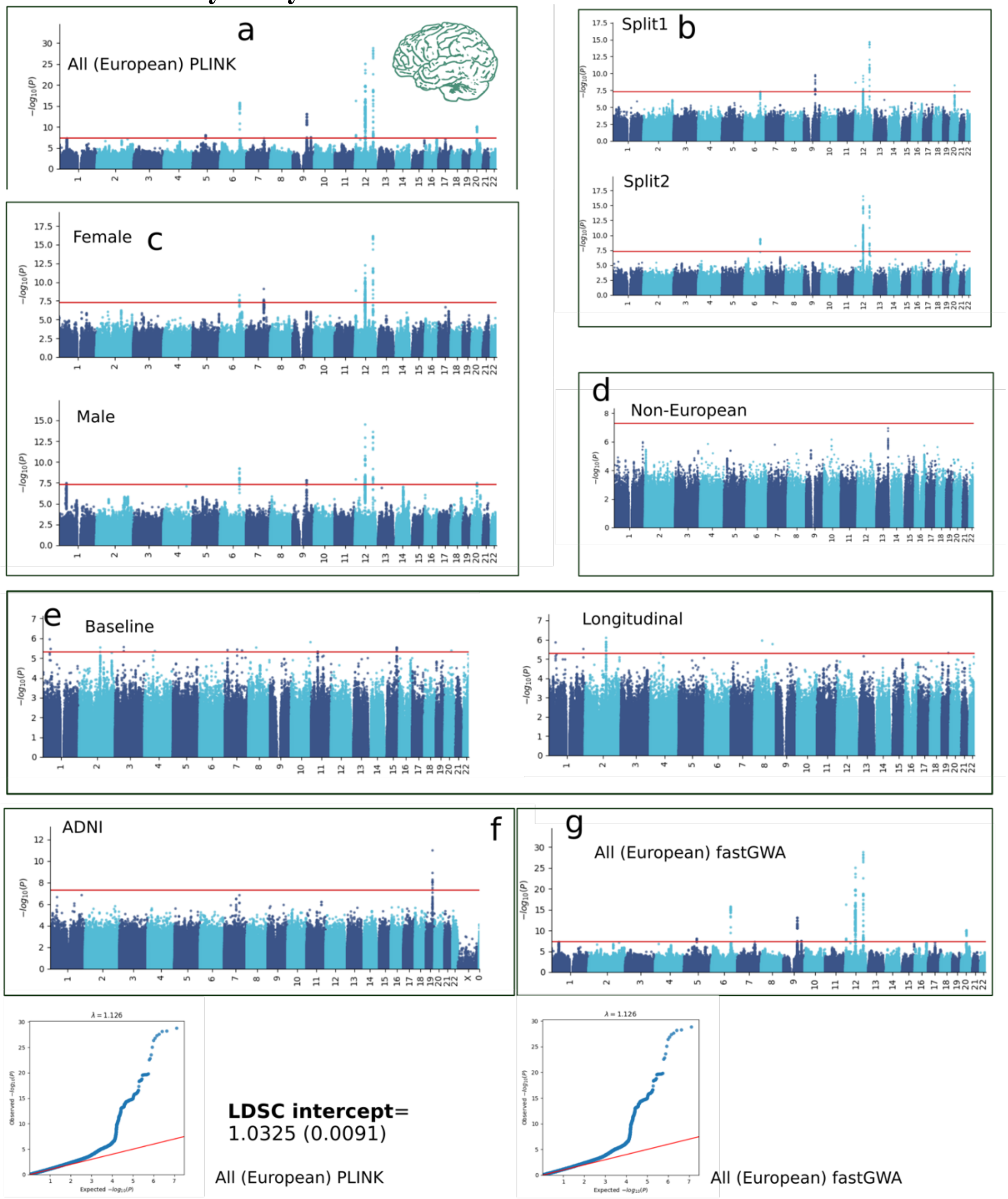


**a-g**) Manhattan plots for AD2 in all European ancestry (PLINK linear models), split1, split2, females, males, non-European ancestries, baseline and longitudinal data, independent ADNI WGS data, and European ancestry (fastGWA). **h-i**) the QQ plot, lambda, and LDSC intercept estimate for PLINK linear models and fastGWA.

**Figure 3: The sensitivity analyses of ASD1 GWAS**

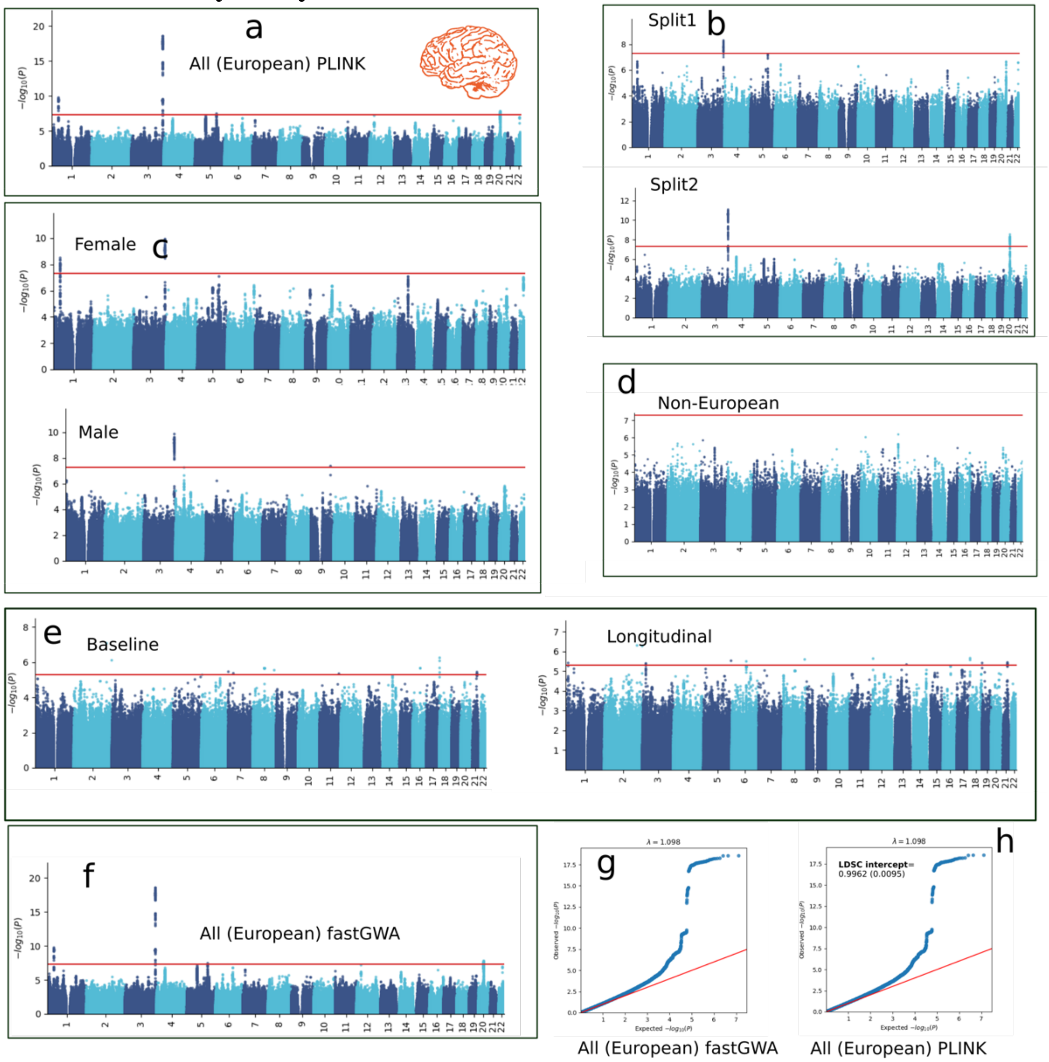


**a-f**) Manhattan plots for ASD1 in all European ancestry (PLINK linear models), split1, split2, females, males, non-European ancestries, baseline and longitudinal data, and European ancestry (fastGWA). **g-h**) the QQ plot, lambda, LDSC intercept estimate for fastGWA and PLINK linear models.

**Figure 4: The sensitivity analyses of ASD2 GWAS**

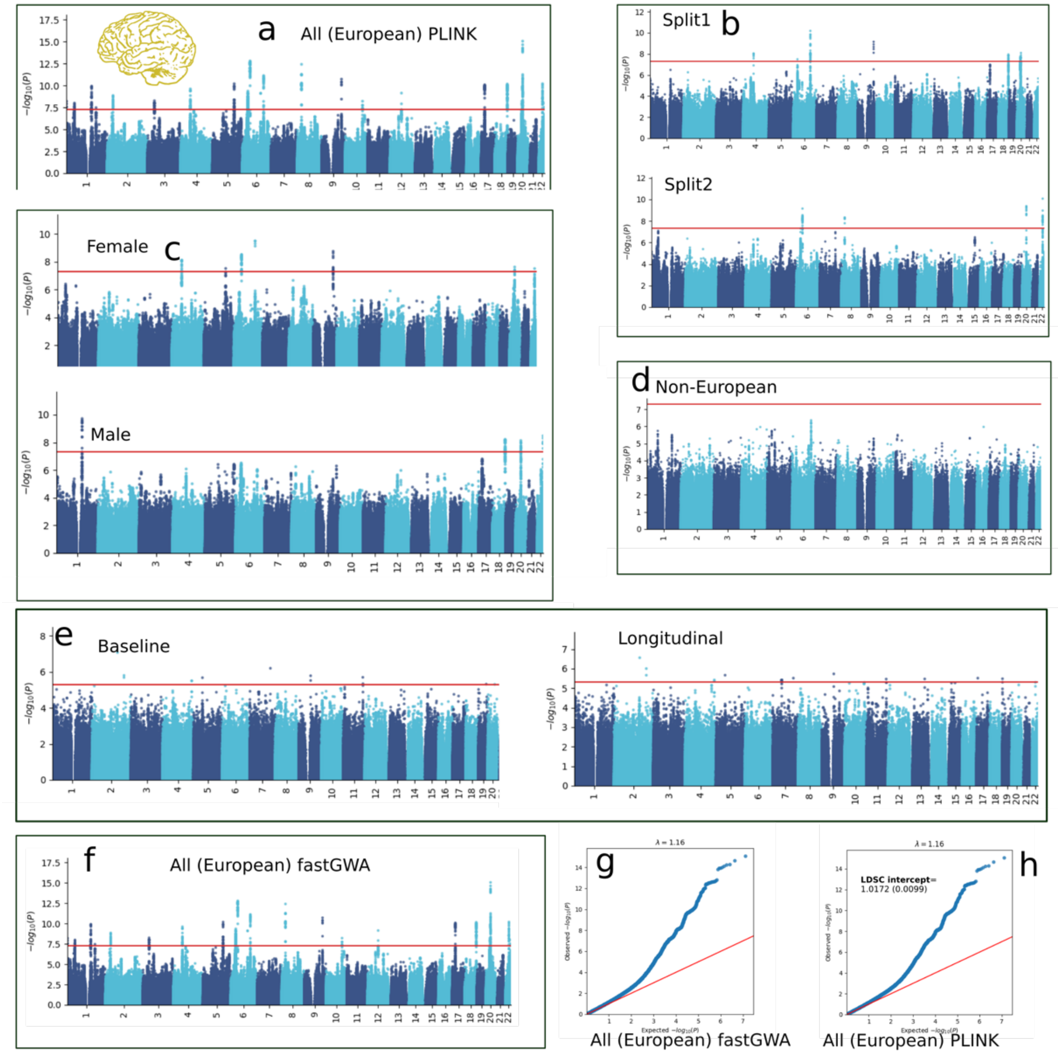


**a-f**) Manhattan plots for ASD2 in all European ancestry (PLINK linear models), split1, split2, females, males, non-European ancestries, baseline and longitudinal data, and European ancestry (fastGWA). **g-h**) the QQ plot, lambda, LDSC intercept estimate for fastGWA and PLINK linear models.

**Figure 5: The sensitivity analyses of ASD3 GWAS**

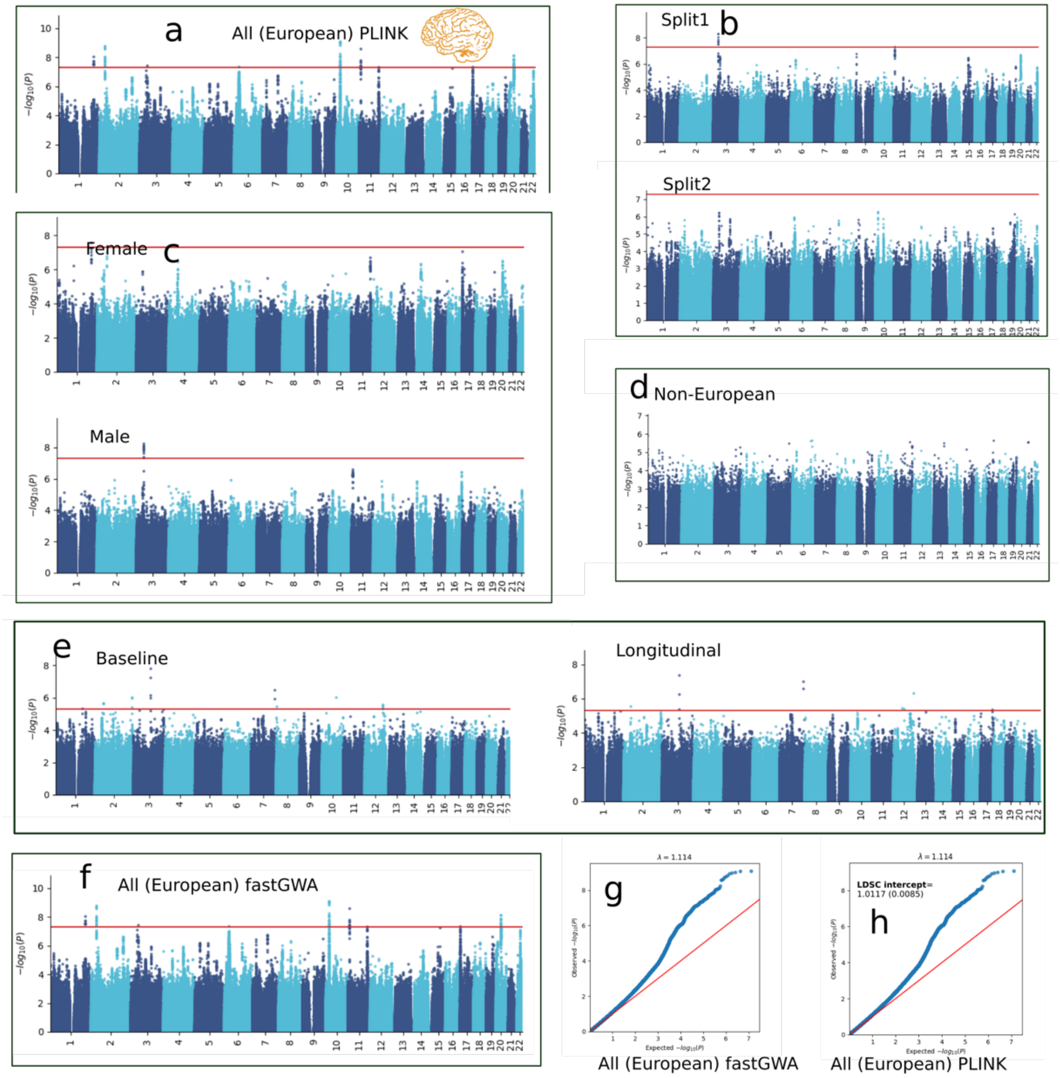


**a-f**) Manhattan plots for ASD3 in all European ancestry (PLINK linear models), split1, split2, females, males, non-European ancestries, baseline and longitudinal data, and European ancestry (fastGWA). **g-h**) the QQ plot, lambda, LDSC intercept estimate for fastGWA and PLINK linear models.

**Figure 6: The sensitivity analyses of LLD1 GWAS**

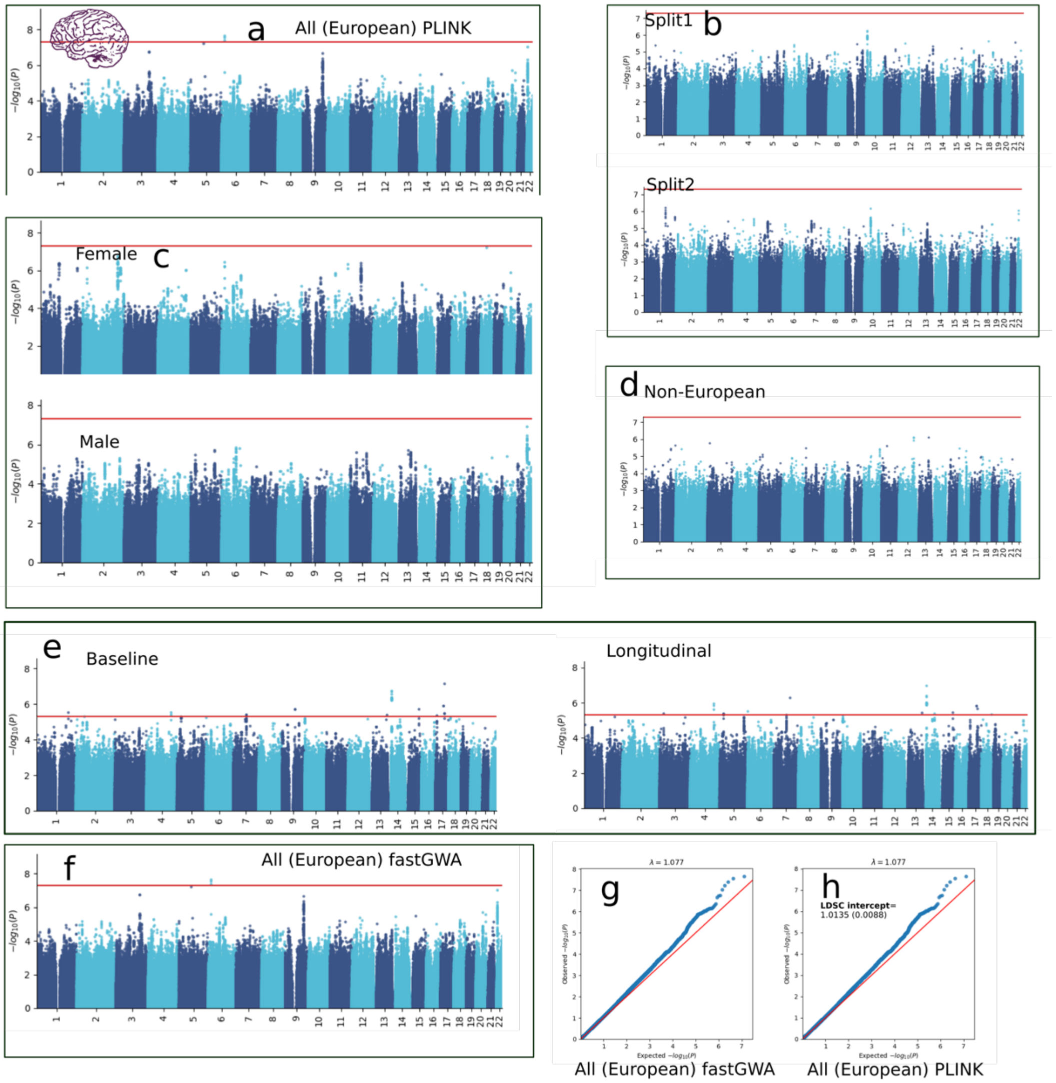


**a-f**) Manhattan plots for LLD1 in all European ancestry (PLINK linear models), split1, split2, females, males, non-European ancestries, baseline and longitudinal data, and European ancestry (fastGWA). **g-h**) the QQ plot, lambda, LDSC intercept estimate for fastGWA and PLINK linear models.

**Figure 7: The sensitivity analyses of LLD2 GWAS**

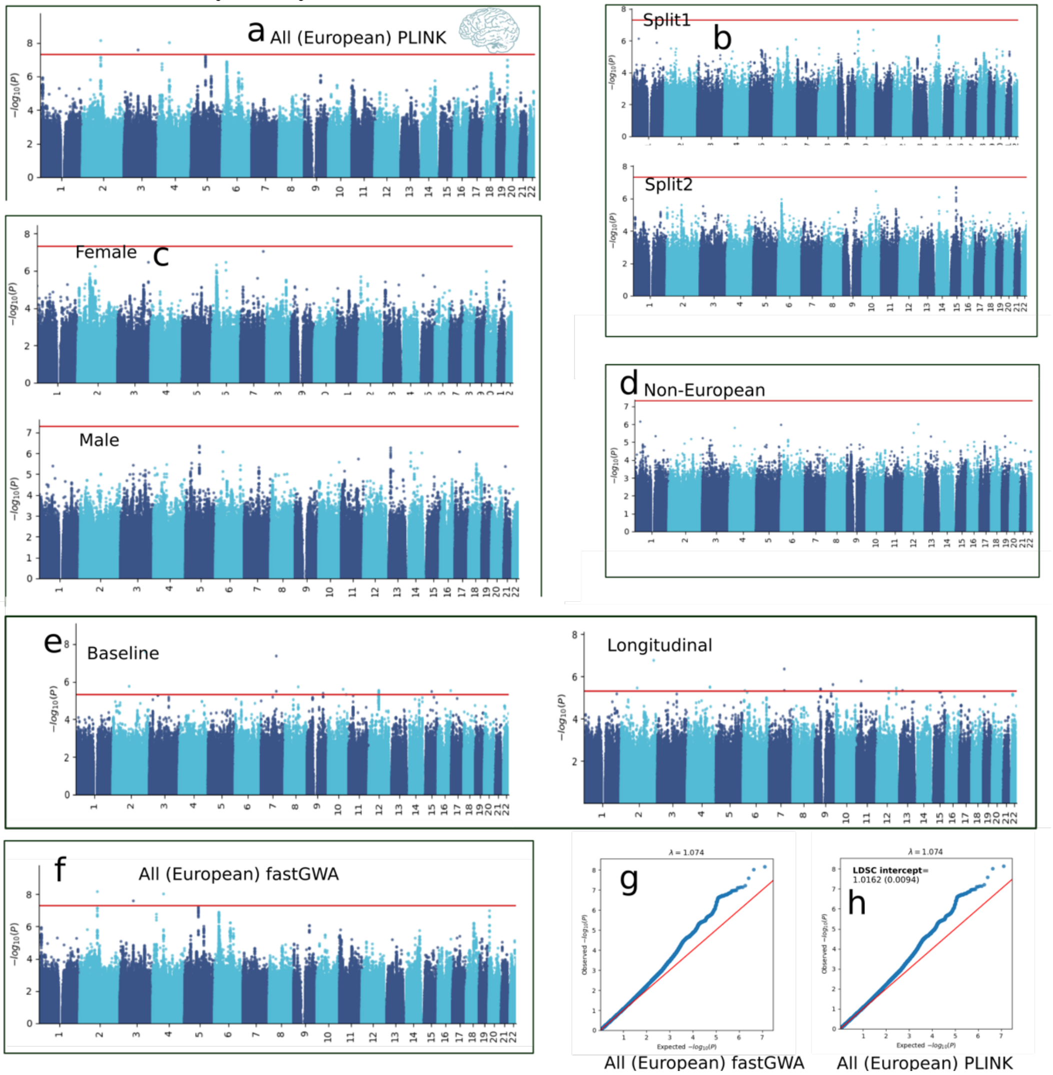


**a-f**) Manhattan plots for LLD2 in all European ancestry (PLINK linear models), split1, split2, females, males, non-European ancestries, baseline and longitudinal data, and European ancestry (fastGWA). **g-h**) the QQ plot, lambda, LDSC intercept estimate for fastGWA and PLINK linear models.

**Figure 8: The sensitivity analyses of SCZ1 GWAS**

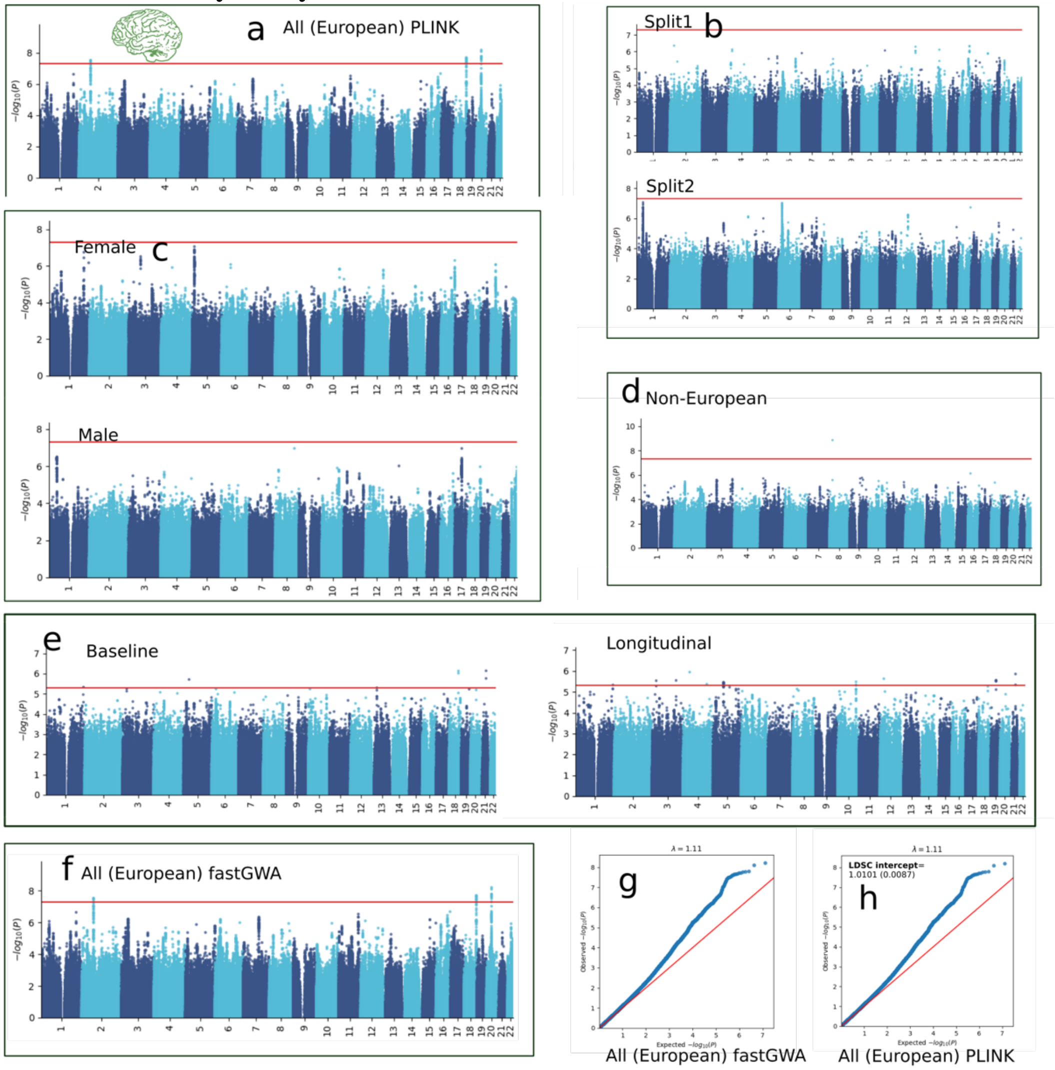


**a-f**) Manhattan plots for SCZ1 in all European ancestry (PLINK linear models), split1, split2, females, males, non-European ancestries, baseline and longitudinal data, and European ancestry (fastGWA). **g-h**) the QQ plot, lambda, LDSC intercept estimate for fastGWA and PLINK linear models.

**Figure 9: The sensitivity analyses of SCZ2 GWAS**

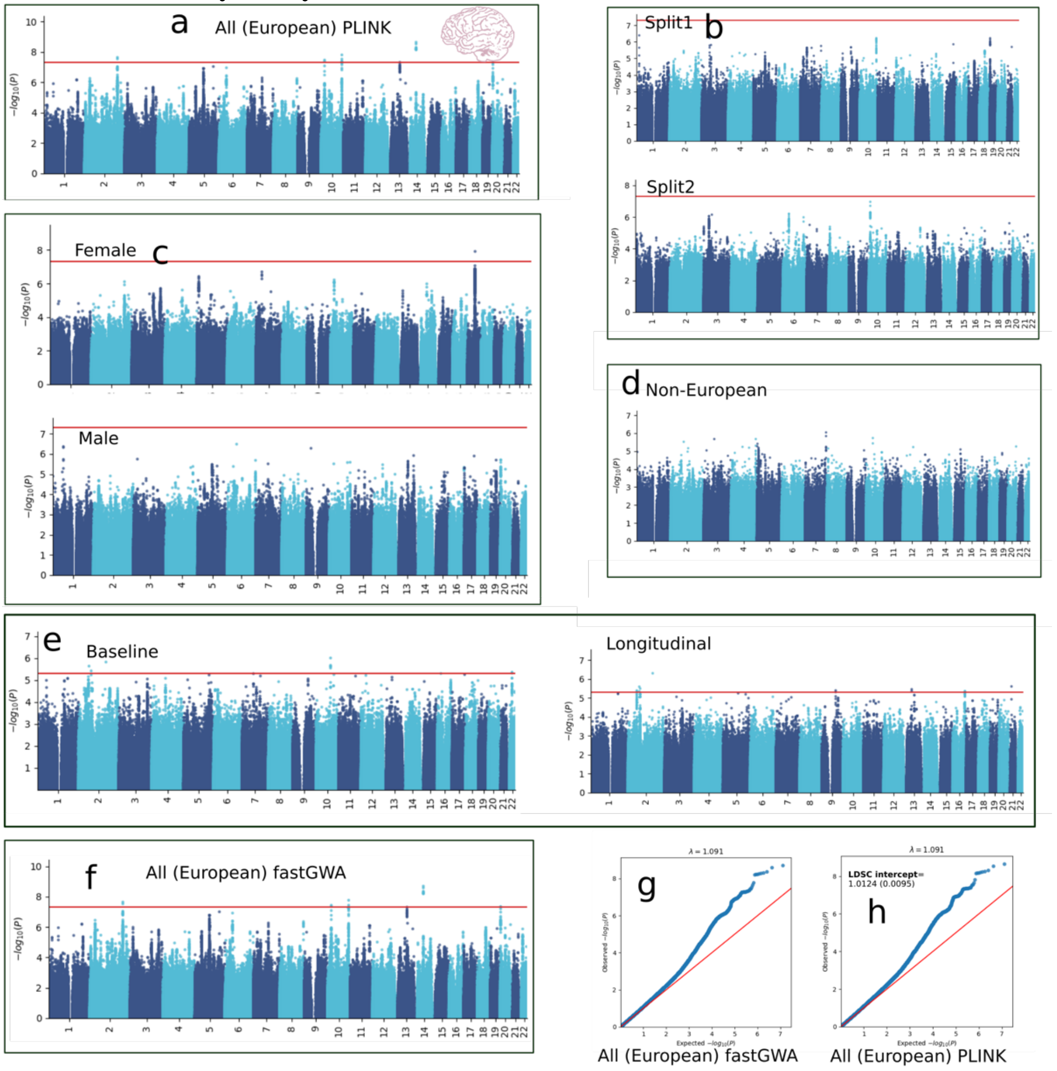


**a-f**) Manhattan plots for SCZ2 in all European ancestry (PLINK linear models), split1, split2, females, males, non-European ancestries, baseline and longitudinal data, and European ancestry (fastGWA). **g-h**) the QQ plot, lambda, LDSC intercept estimate for fastGWA and PLINK linear models.

**Figure 10: Exemplary genomic locus for each BAG in the nine human organ systems**

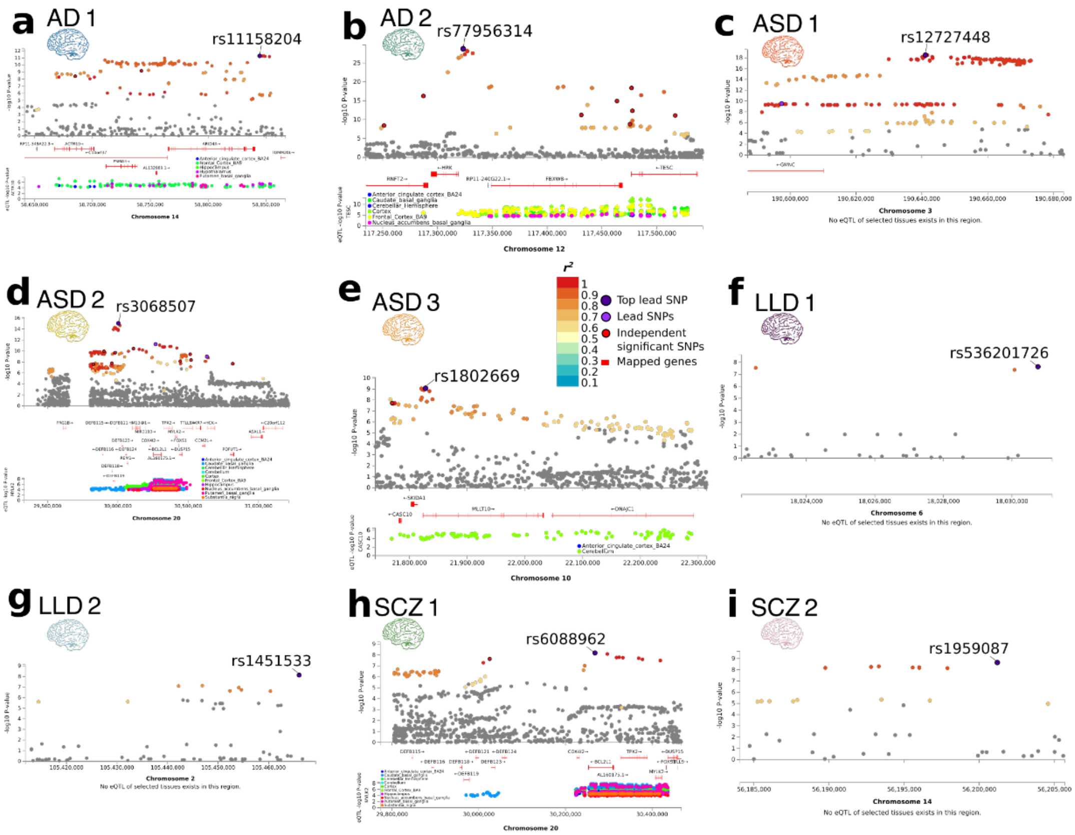


**a-i**) The exemplary genomic locus with the most significant signals for the nine DNEs. The top lead SNP, lead SNPs, and independent significant SNPs are annotated within each locus. We mapped the SNPs to the genes and predicted eQTL in different brain tissues using GETx v8 data. If significant results exist, an exemplary gene was shown for the eQTL analysis for each DNE.

**Figure 11: Phenome-wide associations of the causal SNP (rs9257566) in the GWAS Catalog and Ensembl**

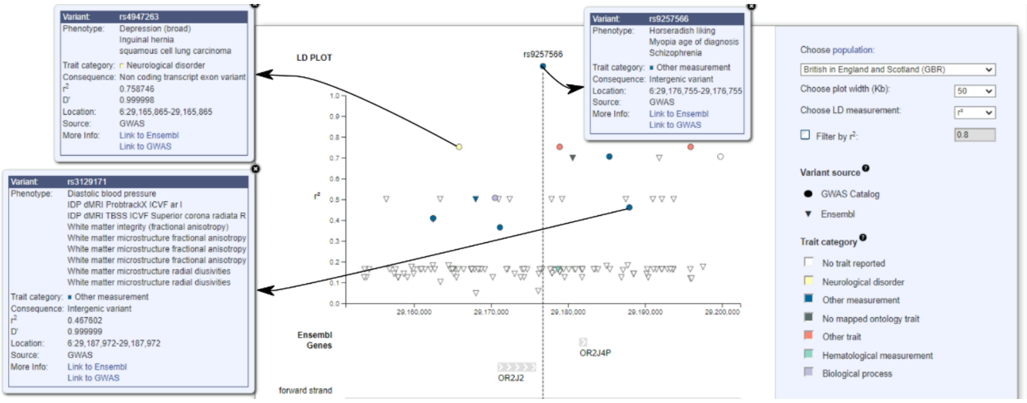


We manually queried the causal SNP in the GWAS Catalog on July 12th 2023: https://www.ebi.ac.uk/gwas/variants/rs9257566.

**Figure 12: Sensitivity check on the prior probability of *p12* for Genetic colocalization analyses**

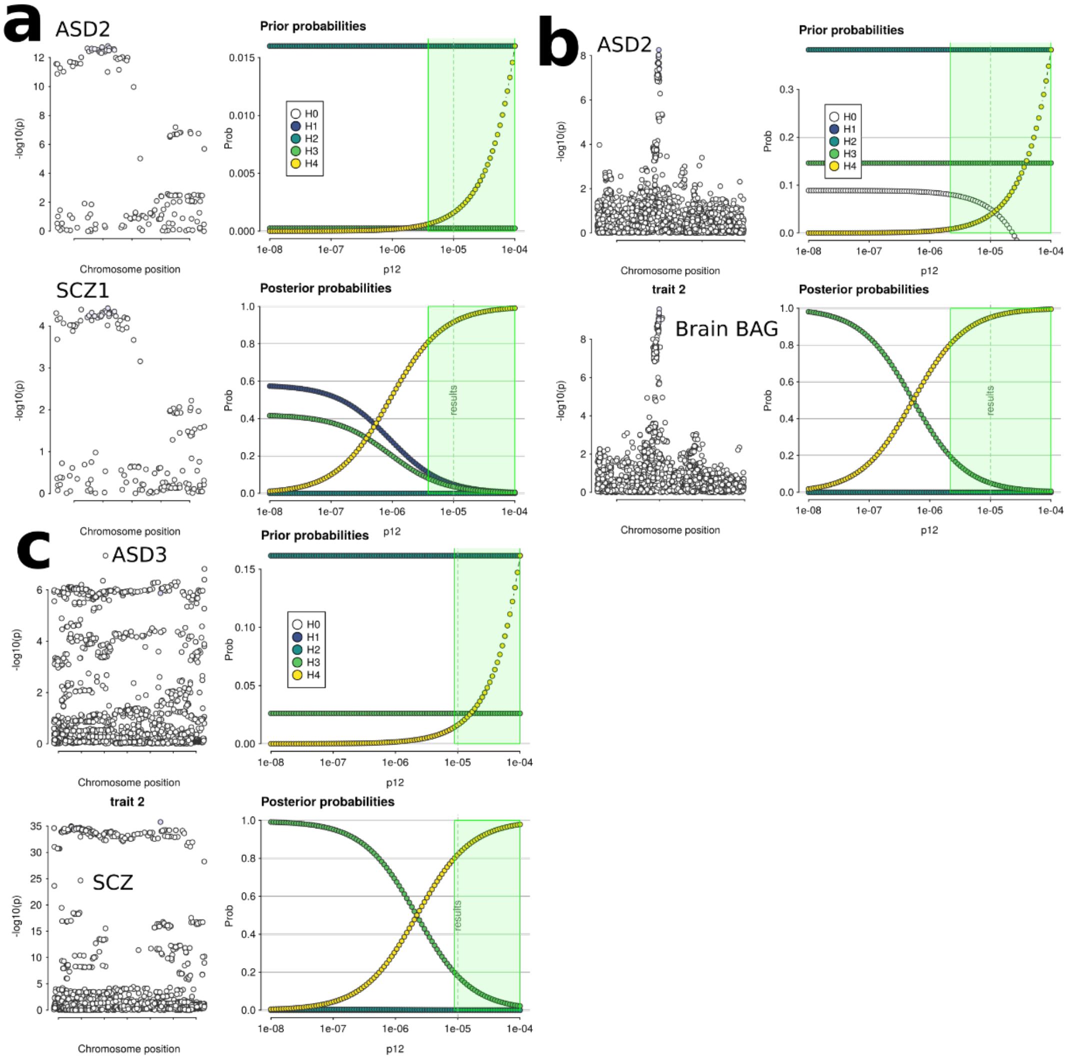


**a**) The sensitivity check for the prior probability of ***p12*** for the shared causal SNP between ASD2 and SCZ1. **b**) The sensitivity check for the prior probability of ***p12*** for the shared causal SNP between ASD2 and brain BAG. **c**) The sensitivity check for the prior probability of ***p12*** for the shared causal SNP between ASD3 and SCZ.

**Figure 13: Genetic colocalization results between the nine DNEs, nine BAGs, and six neurodegenerative and neuropsychiatric disorders from PGC**

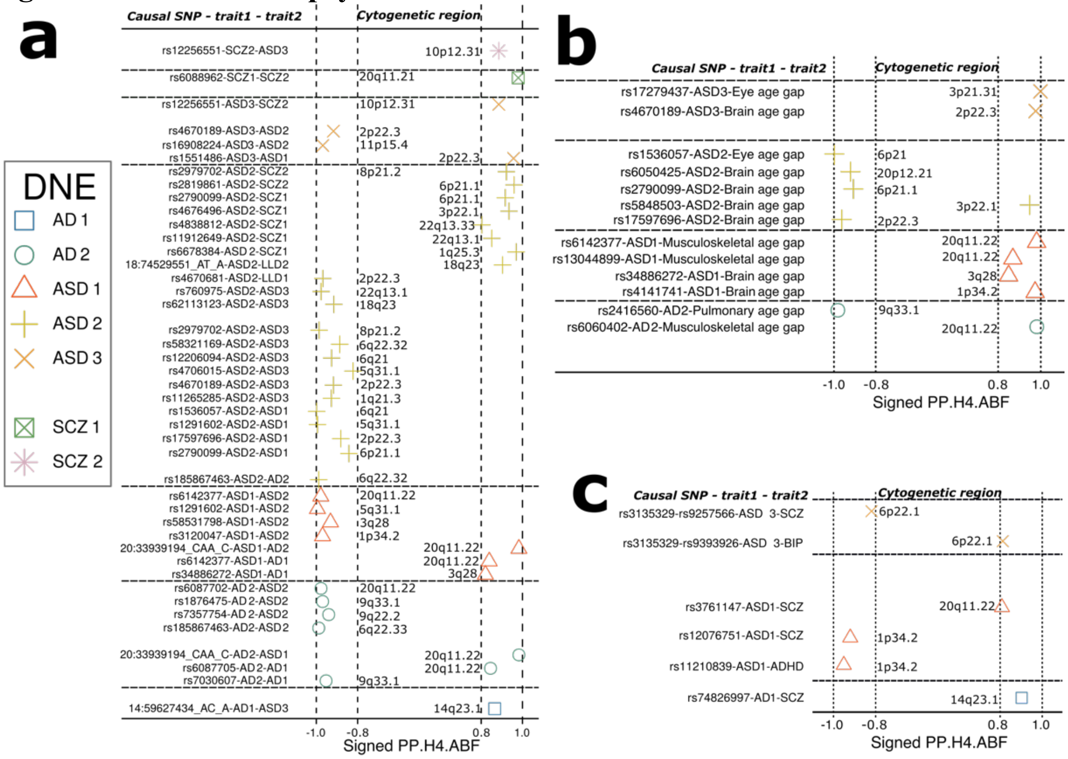


**a**) The identified colocalization signals between pairwise DNEs. The causal SNP for both traits and their cytogenetic region are annotated. The sign of the PP.H4.ABF is determined by the $\beta$ values of the causal SNP for both traits. **b**) The identified colocalization signals between the nine DNEs and the nine BAGs. **c**) The identified colocalization signals between the nine DNEs and the six neurodegenerative and neuropsychiatric disorders from PGC.

**Figure 14: Mendelian randomization sensitivity check for the brain BAG on LLD1**

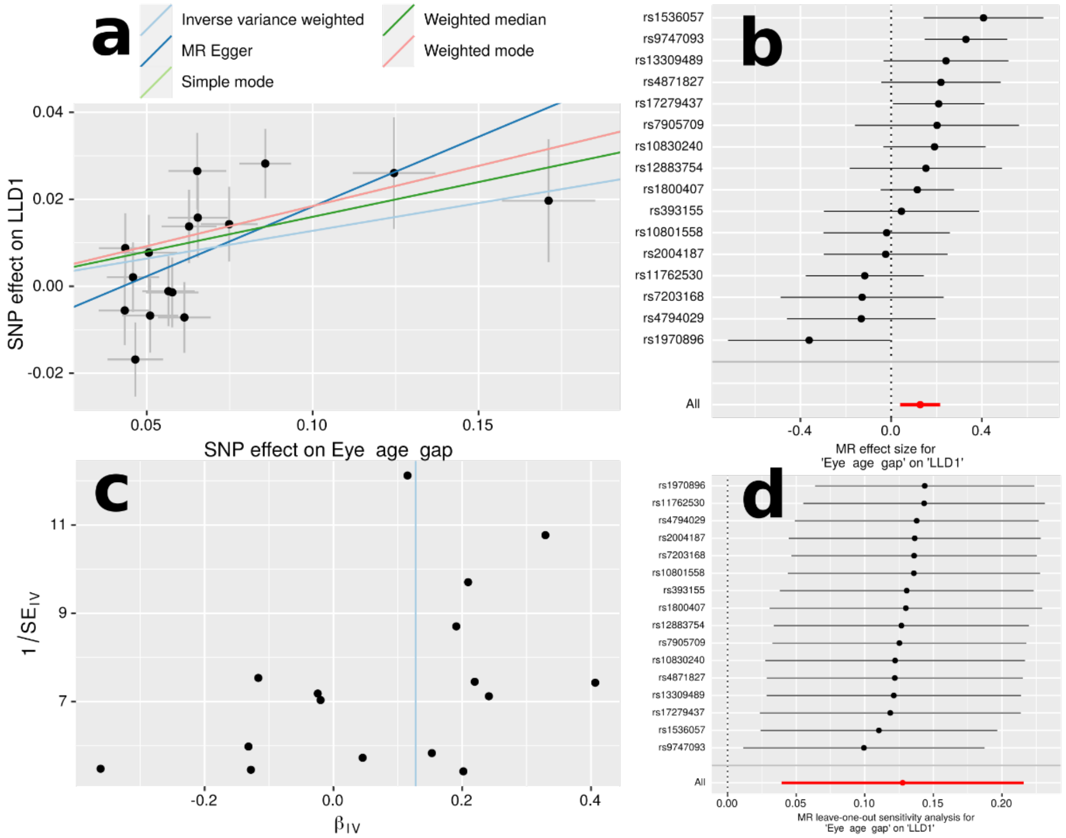


**a**) Scatter plot for the MR effect sizes of the exposure variable (*x*-axis, SD units) and the outcome variable (*y*-axis, log OR) with standard error bars. The slopes of the regression line correspond to the causal effect sizes estimated by the five different estimators. **b**) Forest plot for the single-SNP MR results. Each line represents the MR effect (log OR) for the exposure variable on the outcome variable using only one SNP; the red line shows the MR effect using all SNPs together. **c**) Funnel plot for the relationship between the causal effect of the exposure variable on the outcome variable. Each dot represents MR effect sizes estimated using each SNP as a separate instrument against the inverse of the standard error of the causal estimate. The vertical red line shows the MR estimates using all SNPs. **d**) Leave-one-out analysis of the exposure variable on the outcome variable. Each row represents the MR effect (log OR) and the 95% CI by excluding that SNP from the analysis. The red line depicts the IVW estimator using all SNPs.

**Figure 15: Mendelian randomization sensitivity check for the cardiovascular BAG on ASD3**

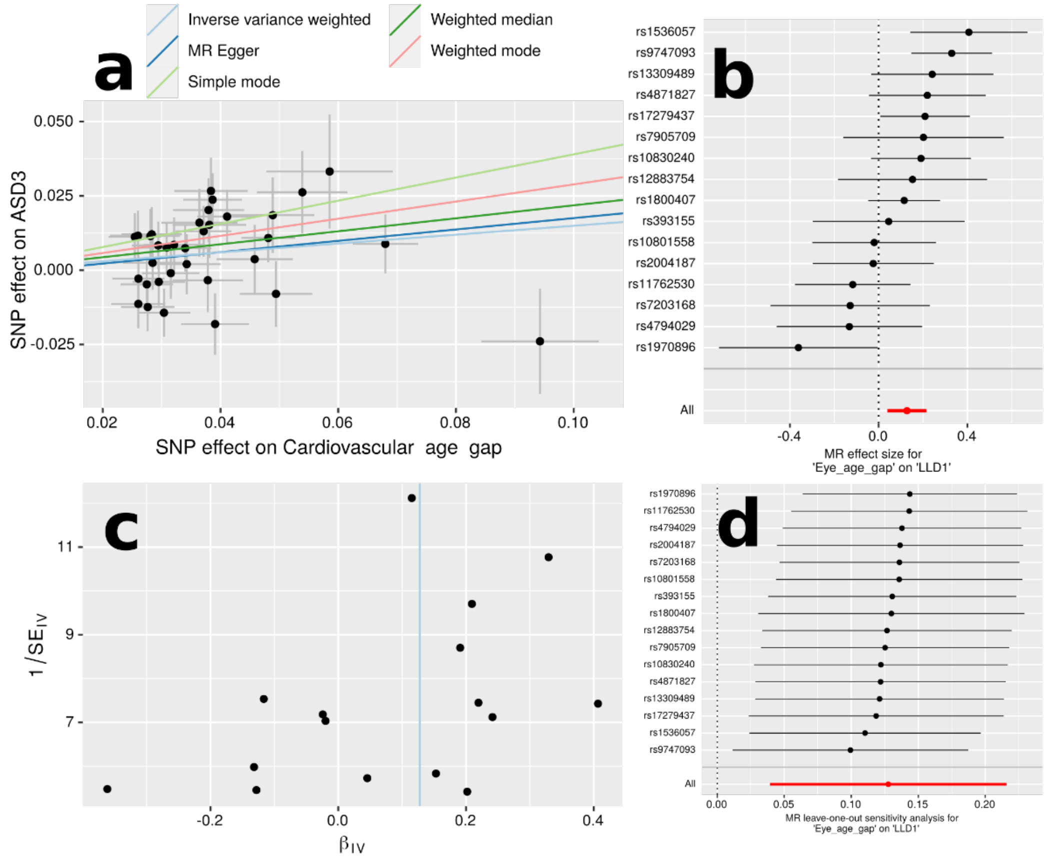


**a**) Scatter plot for the MR effect sizes of the exposure variable (*x*-axis, SD units) and the outcome variable (*y*-axis, log OR) with standard error bars. The slopes of the regression line correspond to the causal effect sizes estimated by the five different estimators. **b**) Forest plot for the single-SNP MR results. Each line represents the MR effect (log OR) for the exposure variable on the outcome variable using only one SNP; the red line shows the MR effect using all SNPs together. **c**) Funnel plot for the relationship between the causal effect of the exposure variable on the outcome variable. Each dot represents MR effect sizes estimated using each SNP as a separate instrument against the inverse of the standard error of the causal estimate. The vertical red line shows the MR estimates using all SNPs. **d**) Leave-one-out analysis of the exposure variable on the outcome variable. Each row represents the MR effect (log OR) and the 95% CI by excluding that SNP from the analysis. The red line depicts the IVW estimator using all SNPs.

**Figure 16: Mendelian randomization sensitivity check for the pulmonary BAG on LLD2**

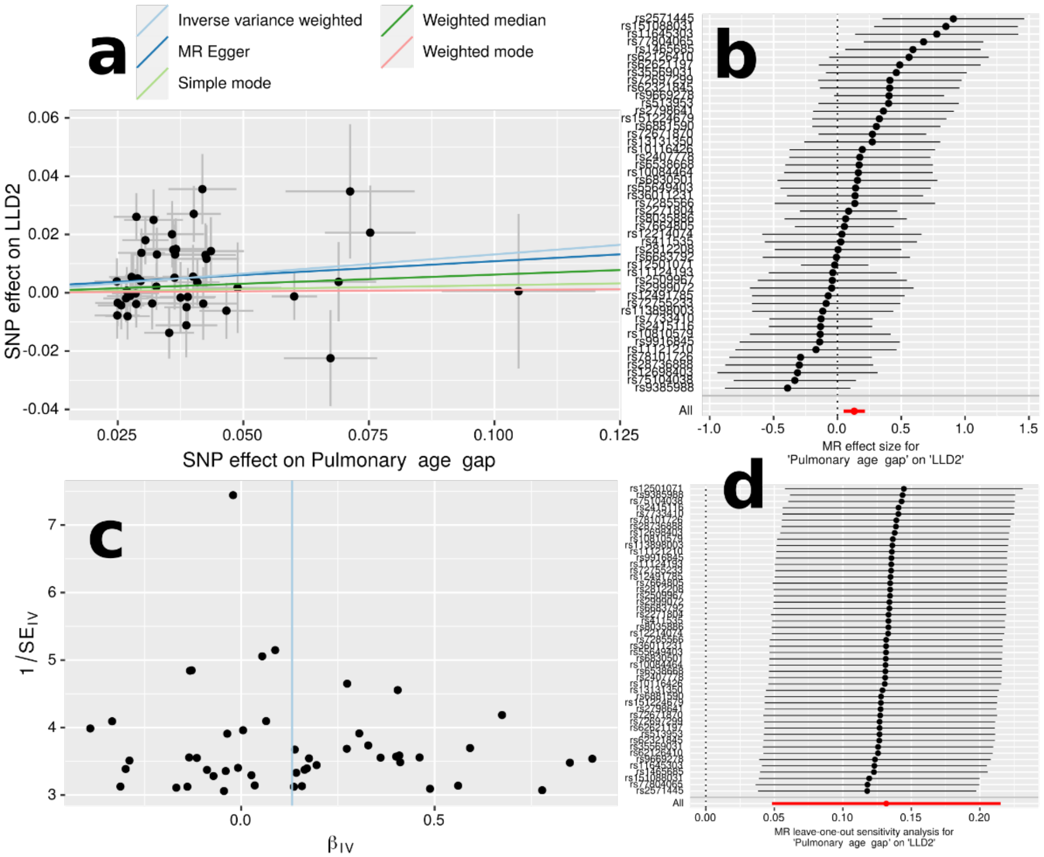


**a**) Scatter plot for the MR effect sizes of the exposure variable (*x*-axis, SD units) and the outcome variable (*y*-axis, log OR) with standard error bars. The slopes of the regression line correspond to the causal effect sizes estimated by the five different estimators. **b**) Forest plot for the single-SNP MR results. Each line represents the MR effect (log OR) for the exposure variable on the outcome variable using only one SNP; the red line shows the MR effect using all SNPs together. **c**) Funnel plot for the relationship between the causal effect of the exposure variable on the outcome variable. Each dot represents MR effect sizes estimated using each SNP as a separate instrument against the inverse of the standard error of the causal estimate. The vertical red line shows the MR estimates using all SNPs. **d**) Leave-one-out analysis of the exposure variable on the outcome variable. Each row represents the MR effect (log OR) and the 95% CI by excluding that SNP from the analysis. The red line depicts the IVW estimator using all SNPs.

**Figure 17: Mendelian randomization sensitivity check for AD2 on AD**

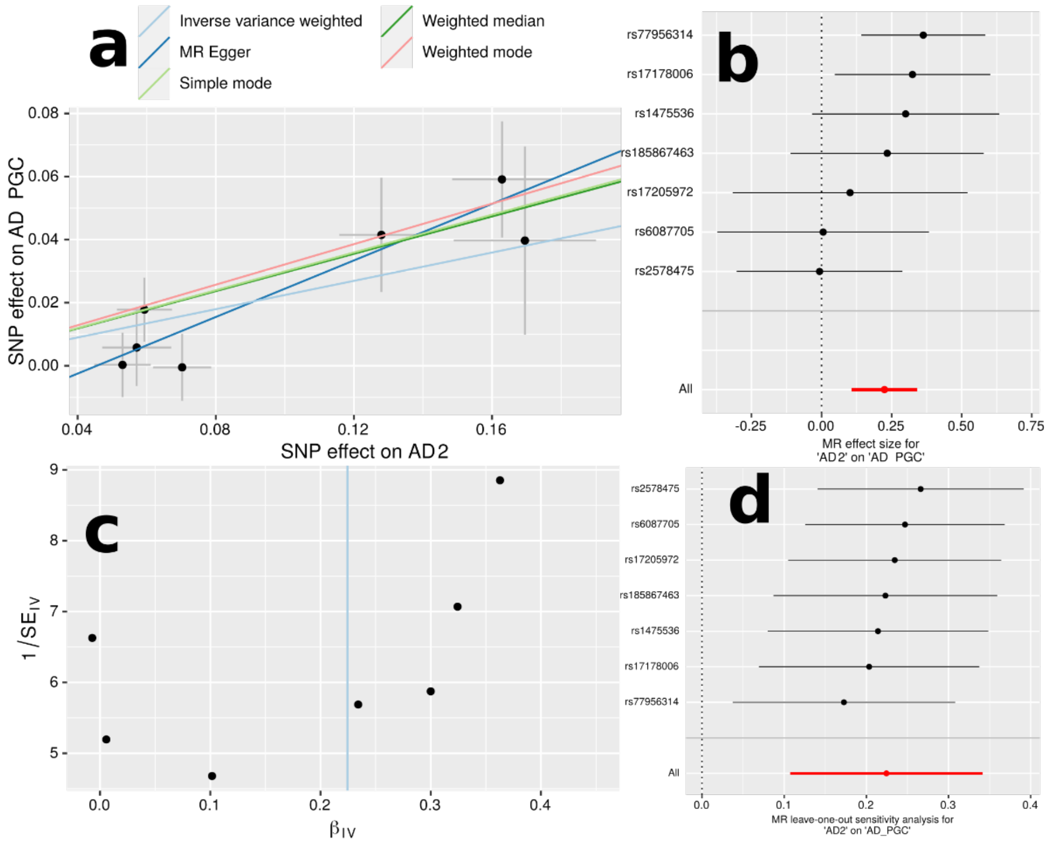


**a**) Scatter plot for the MR effect sizes of the exposure variable (*x*-axis, SD units) and the outcome variable (*y*-axis, log OR) with standard error bars. The slopes of the regression line correspond to the causal effect sizes estimated by the five different estimators. **b**) Forest plot for the single-SNP MR results. Each line represents the MR effect (log OR) for the exposure variable on the outcome variable using only one SNP; the red line shows the MR effect using all SNPs together. **c**) Funnel plot for the relationship between the causal effect of the exposure variable on the outcome variable. Each dot represents MR effect sizes estimated using each SNP as a separate instrument against the inverse of the standard error of the causal estimate. The vertical red line shows the MR estimates using all SNPs. **d**) Leave-one-out analysis of the exposure variable on the outcome variable. Each row represents the MR effect (log OR) and the 95% CI by excluding that SNP from the analysis. The red line depicts the IVW estimator using all SNPs.

**Figure 18: Incremental $R^2$ of the nine DNEs to predict the 14 disease categories using only the PRS target population**

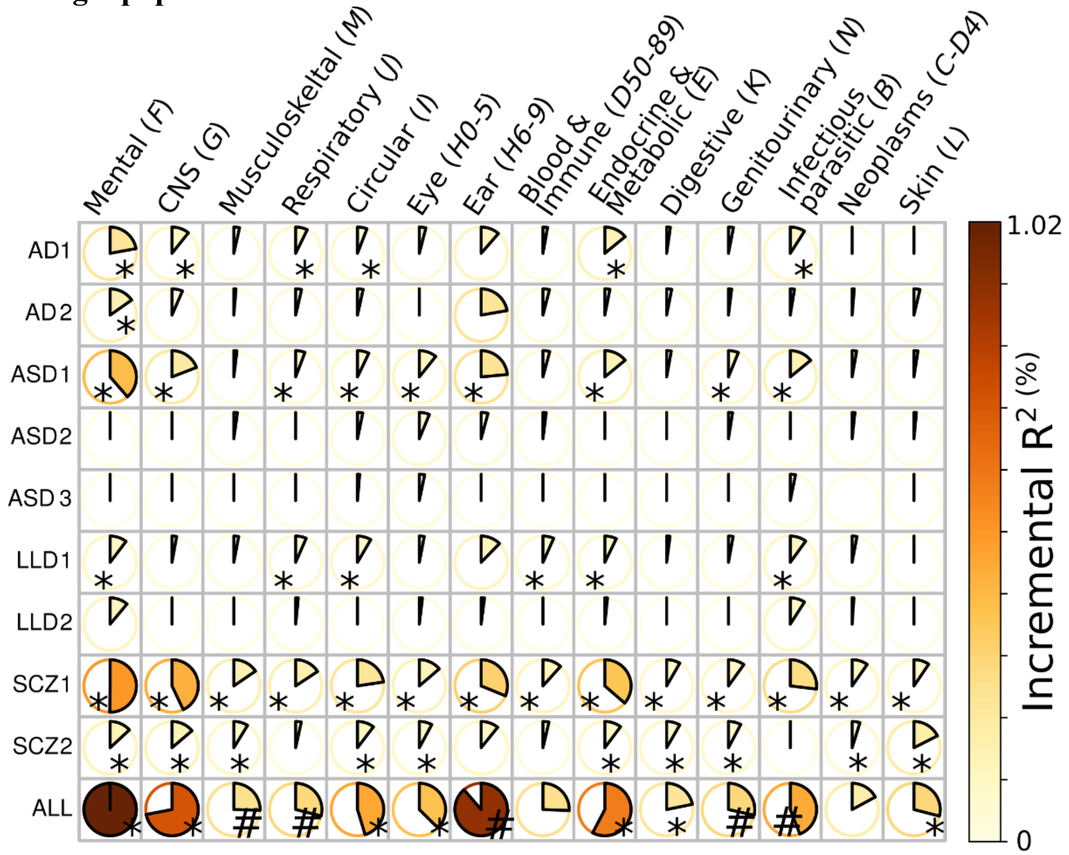


We redid the linear regression using the nine DNEs from only the RPS target population ($N$=15,891) to fairly compare the results with the nine PRSs. # indicates a nominal significance level (0.05); * represents results passing the Bonferroni correction. The DNEs can significantly enhance prediction performance when combined with other commonly available features (e.g., age and sex).

**Figure 19: Incremental $R^2$ of the nine PRSs derived from PLINK to explain the nine DNEs in the PRS target population**

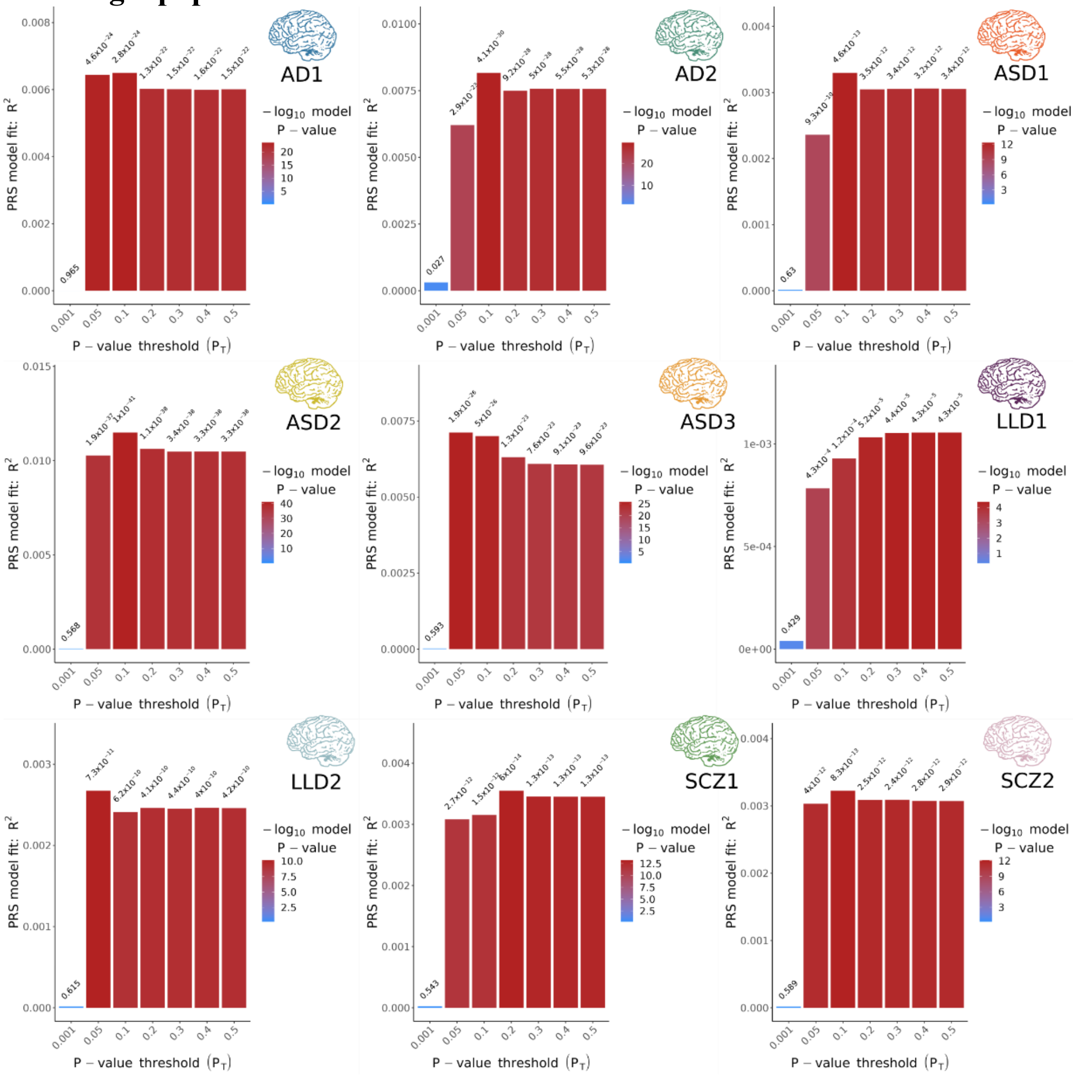


We used a linear regression model to fit the nine PRSs and other covariates to predict the phenotypes (i.e., the nine DENs). The incremental $R^2$ indicates the variance that each PRS additionally contributes to the phenotype of interest.

**Figure 20: Incremental $R^2$ of the nine PRSs derived from PRS-CS to explain the nine DNEs in the PRS target population**

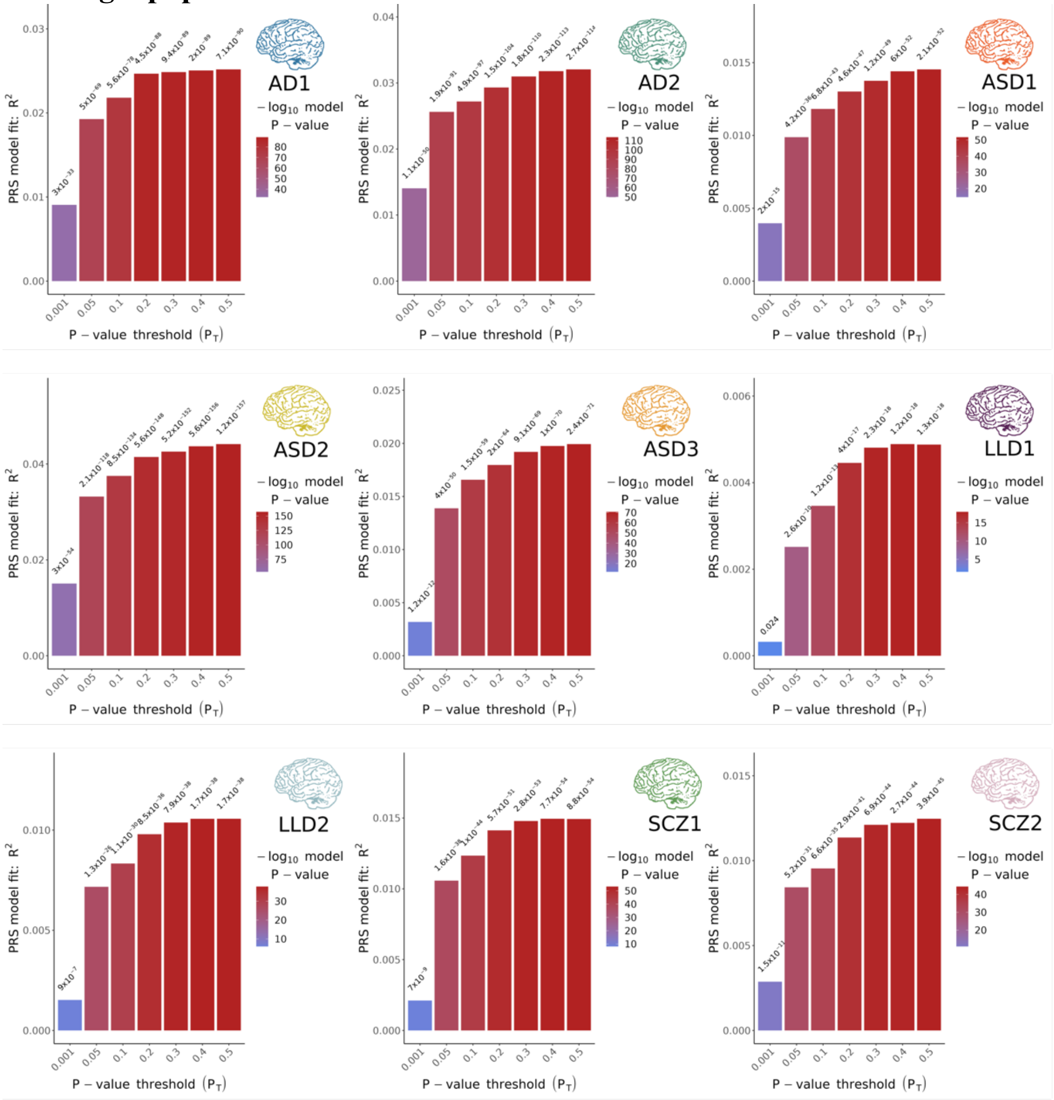


We used a linear regression model to fit the nine PRSs and other covariates to predict the phenotypes (i.e., the nine DENs). The incremental $R^2$ indicates the variance that each PRS additionally contributes to the phenotype of interest. We use the threshold method here to fairly compare the performance between PLINK and PRS-CS (with the same set of SNPs), although PRS-CS does not require a clumping procedure and considers LD through modeling.

**Figure 21: Scatter plot for the nine PRSs derived from PLINK and PRS-CS**

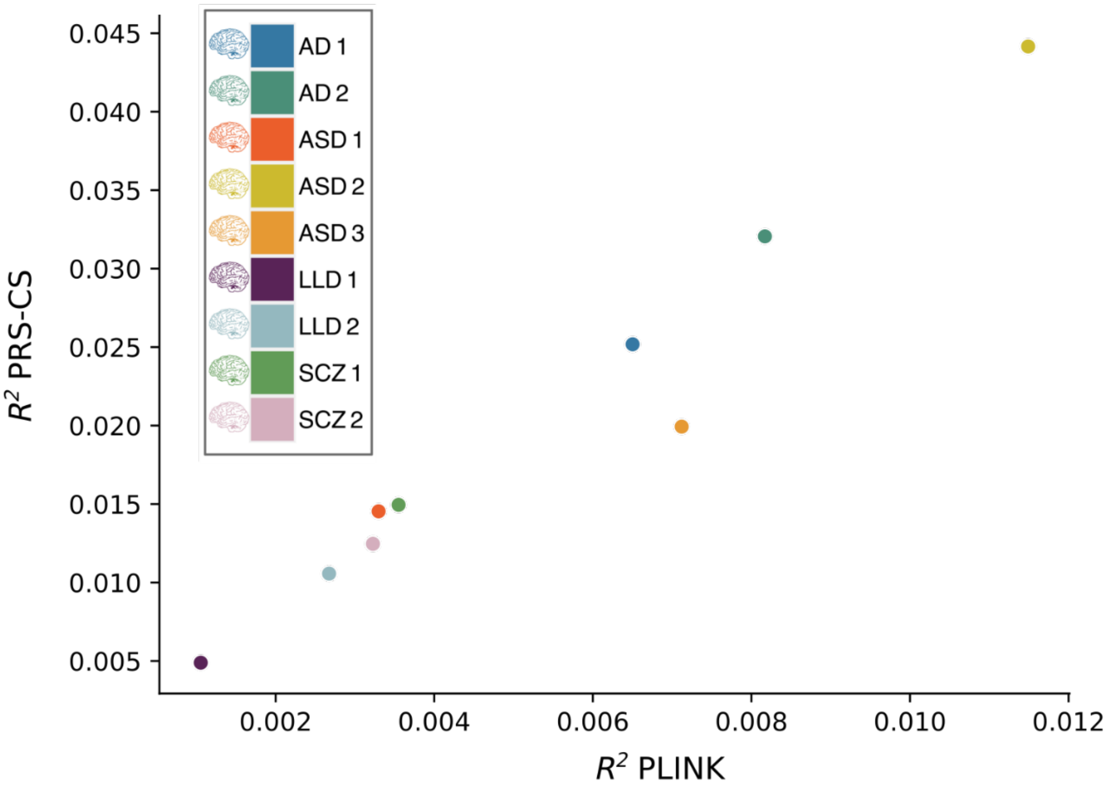

**Figure 22: Incremental $R^2$ of the nine DNEs to predict the 14 disease categories using the nine PRSs derived from PRS-CS for only the PRS target population**

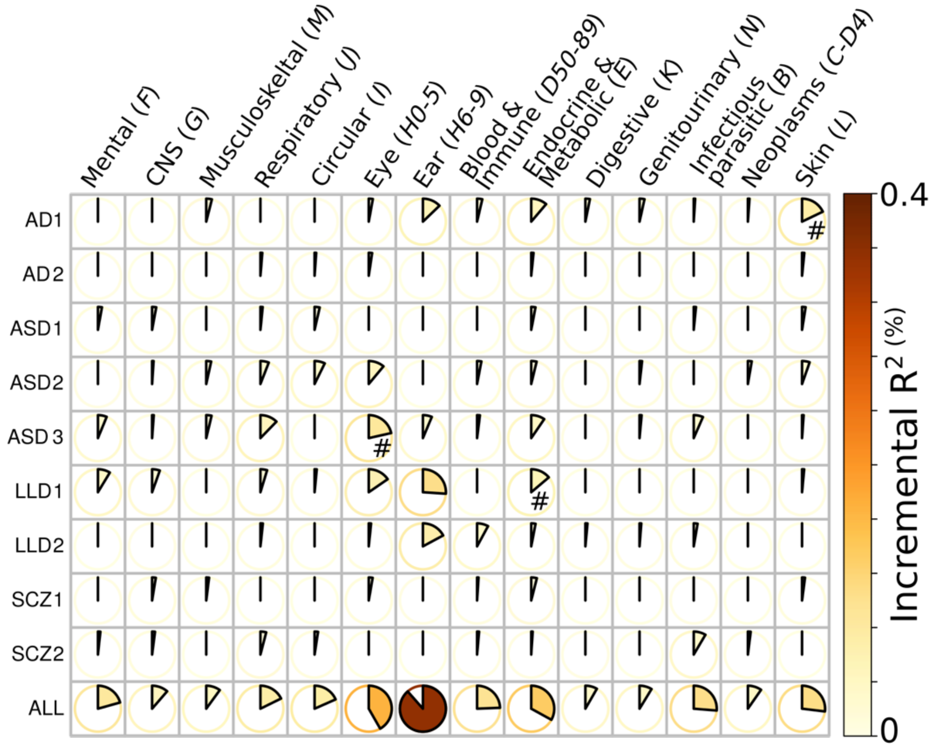


We redid the linear regression using the nine DNEs derived by the PRS-CS method from only the RPS target population (*N*=15,891). # indicates a nominal significance level (0.05).

**Figure 23: Model considerations and illustrations of the semi-supervised clustering approaches**

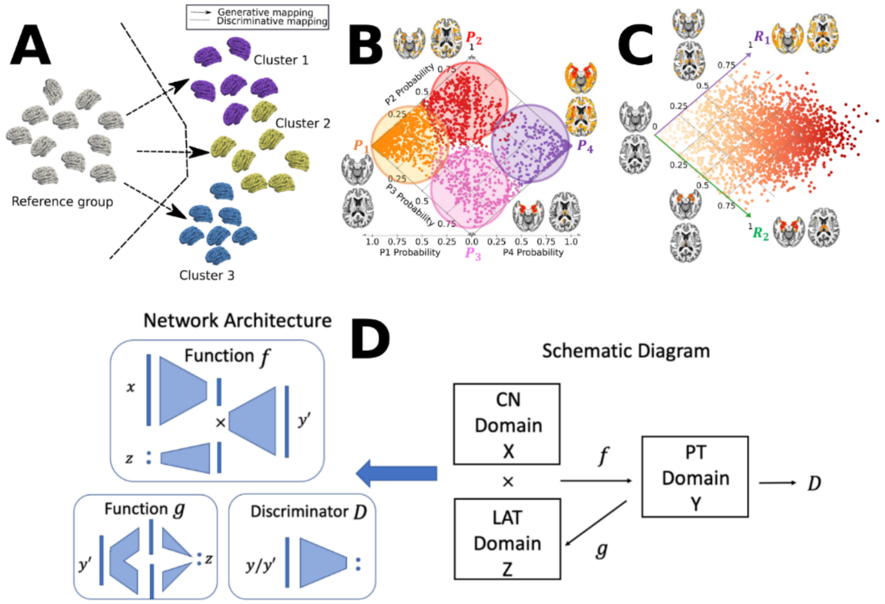


**A**) Schematic figures of semi-supervised clustering methods, including generative[1,2,17] and discriminative approaches.[18,19] Semi-supervised clustering methods dissect the neuroanatomical heterogeneity of brain diseases by seeking the so-called "*1-to-k*" mapping between the healthy control (CN) group as a reference and the patient (PT) group as a target. They sought to tease out clusters that are likely driven by distinct pathological trajectories instead of by global similarity/dissimilarity in data. The figure is adapted from our previous work.[20] **B**) Conceptual illustration of applying Smile-GAN ADNI to cluster MCI/AD participants into four hard-coded subtypes (P1, P2, P3, and P4) and two longitudinal pathways after applying the model to longitudinal data: P1→P2→P4; *ii*) P1→P3→P4. The figure is adapted from our previous work.[2] **C**) Conceptual illustration of applying Surreal-GAN to ADNI to recapitulate the neuroanatomical heterogeneity of MCI/AD participants into two continuous dimensional scores. Methodological advances are to capture disease severity and heterogeneity simultaneously using only cross-sectional data and allow the same individuals to express themselves along multiple dimensions. The figure is adapted from our previous work.[1] **D**) General network architectures (left) and schematic diagram of the semi-supervised learning (right, corresponding to Fig. A). The figure is adapted from our previous work.[1]

**Figure 24: Comparisons of *beta* coefficient and P-value between PLINK and fastGWA**

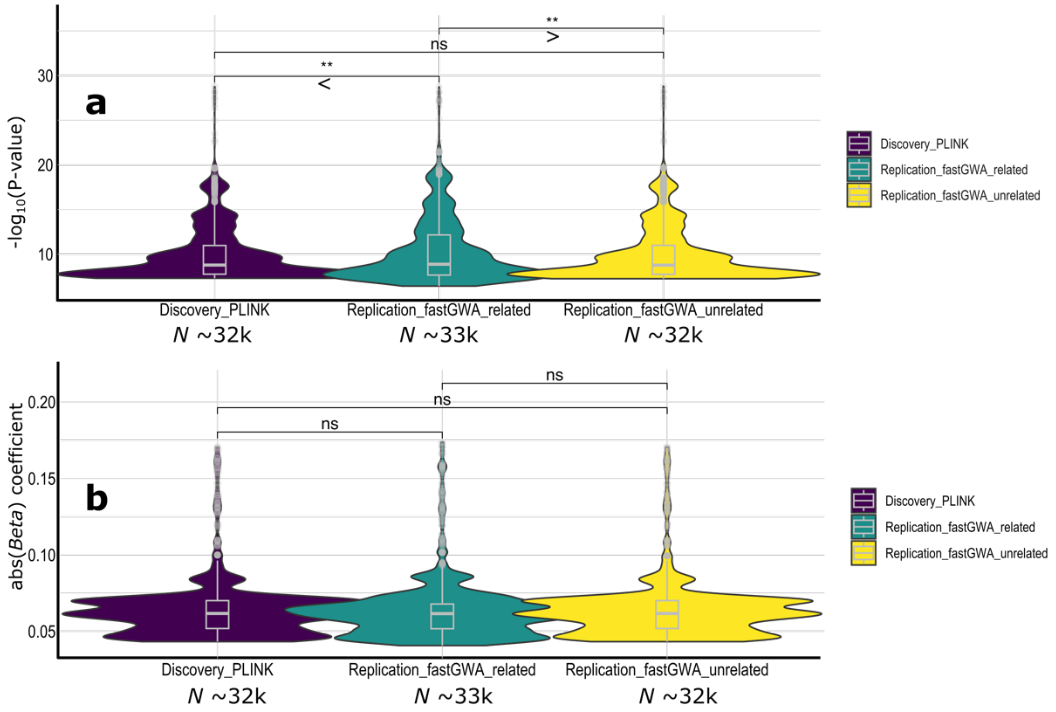


Comparisons on -log10(P-value) and beta coefficient values for the three sets of GWASs: *i*) PLINK (*N*~32k), *ii*) fastGWA with related (*N*~33k), and *iii*) fastGWA without unrelated (a.k.a. to a linear model) (*N*~32k). The fastGWA case with unrelated individuals is similar to the simple linear model implemented in PLINK. The *ns* denotes not significant with a two-sample t-test. The symbol * indicates statistical significance, and the symbol < or > denotes the direction of the mean measurements in the different groups.

**Table 1: Descriptive statistics of the difference between the healthy control and disease groups for the 9 DNEs in the UKBB population.** The LLD group encompasses a broad definition of depression without imposing age restrictions (participants aged 60 years and above). Despite the small sample sizes and imbalanced data in AD, ASD, and SCZ patient groups within UKBB, we presented the P-values derived from both the standard two-sample t-test and a linear regression that included covariates when comparing DNEs between the CN and PT groups.

| DNE | CN (6390) vs PT (*N*) | P-value (t-test) | P-value (regression) |
|---|---|---|---|
| AD1 | 0.37±0.20 vs 0.62±0.28 (*N*=23; AD) | $<1.0x10^{-9}$ | $5.23x10^{-9}$ |
| AD2 | 0.29±0.19 vs 0.45±0.27 (*N*=23; AD) | $1x10^{-4}$ | $1.07x10^{-7}$ |
| ASD1 | -0.82±1.30 vs -1.26±0.88 (*N*=6; ASD) | 0.40 | 0.25 |
| ASD2 | -1.15±1.09 vs -1.05±0.94 (*N*=6; ASD) | 0.80 | 0.73 |
| ASD3 | -1.13±1.03 vs -0.67±0.82 (*N*=6; ASD) | 0.28 | 0.28 |
| LLD1 | -1.22±2.17 vs -1.41±2.09 (*N*=1329; LLD) | $3.08x10^{-3}$ | $4.45x10^{-3}$ |
| LLD2 | -0.45±1.63 vs -0.34±1.65 (*N*=1329; LLD) | $1.58x10^{-2}$ | $2.00x10^{-2}$ |
| SCZ1 | -1.12±2.09 vs 0.17±2.55 (*N*=23; SCZ) | $3.11x10^{-3}$ | $2.41x10^{-3}$ |
| SCZ2 | -1.89±2.29 vs -0.40±2.34 (*N*=23; SCZ) | $1.78x10^{-3}$ | $1.92x10^{-3}$ |

**Table 2: The SNP-based heritability estimates.** We used the GCTA software to estimate the SNP-based heritability of the nine DNEs.

| DNE | $h^2$ | $h^2$ (SE) | P-value | *N* |
|---|---|---|---|---|
| AD1 | 0.49 | 0.02 | $<1x10^{-10}$ | 33,592 |
| AD2 | 0.55 | 0.02 | $<1x10^{-10}$ | 33,592 |
| ASD1 | 0.39 | 0.02 | $<1x10^{-10}$ | 33,592 |
| ASD2 | 0.66 | 0.02 | $<1x10^{-10}$ | 33,592 |
| ASD3 | 0.50 | 0.02 | $<1x10^{-10}$ | 33,592 |
| LLD1 | 0.24 | 0.02 | $<1x10^{-10}$ | 33,592 |
| LLD2 | 0.27 | 0.02 | $<1x10^{-10}$ | 33,592 |
| SCZ1 | 0.42 | 0.02 | $<1x10^{-10}$ | 33,592 |
| SCZ2 | 0.36 | 0.02 | $<1x10^{-10}$ | 33,592 |

**Table 3: The six case-control GWAS on neurodegenerative and neuropsychiatric disorders from PGC (a) and four lifestyle factors and cognitive scores (b).**

**a:**

We downloaded and manually harmonized (e.g., GRCh 37, flip the effect allele, etc.) the 6 GWAS summary statistics from the PGC website (https://pgc.unc.edu/for-researchers/download-results/). Two studies (ASD and OCD) were not used in the Mendelian randomization analyses, in which the allele frequency information was unavailable. In the case of AD GWAS summary statistics, the original dataset lacked a column for SNP numbers. To overcome this limitation, we employed a mapping approach using the chromosome number and position of the AD GWAS summary statistics to dpSNP version 150, which allowed us to obtain the corresponding SNP numbers. Furthermore, for the GWAS summary statistics of ASD and OCD, the authors did not provide the allele frequency information for the effect allele. As a result, we could not include these two diseases in our Mendelian randomization analyses. The *TwoSampleMR* package requires this information to harmonize the GWAS summary statistics precisely.

| Trait | Dataset | URL | PubMed ID | Sample size | Ancestry |
|---|---|---|---|---|---|
| AD | PGC | https://ctg.cncr.nl/software/summary_statistics | 34493870 | 1,126,536 | European |
| ADHD | PGC | https://figshare.com/articles/dataset/adhd2022/22564390 | 36702997 | 225,534 | European |
| ASD | PGC | https://figshare.com/articles/dataset/asd2019/14671989 | 30804558 | 46,351 | European |
| BIP | PGC | https://figshare.com/articles/dataset/PGC3_bipolar_disorder_GWAS_summary_statistics/14102594 | 34002096 | 413,466 | European |
| OCD | PGC | https://figshare.com/articles/dataset/ocd2018/14672103 | 28761083 | 9725 | European |
| SCZ | PGC | https://figshare.com/articles/dataset/scz2022/19426775 | 35396580 | 320,404 | European |

**b:**

We downloaded and manually harmonized (e.g., GRCh 37, flip the effect allele, etc.) the GWAS summary statistics from the GWAS Catalog (https://www.ebi.ac.uk/gwas/). We finally included computer use time, education (years), intelligence, and reaction time in our genetic correlation analysis. We did not include these data in colocalization and Mendelian randomization analysis.

| Trait | URL | PubMed ID | Sample size | Ancestry |
|---|---|---|---|---|
| Computer use | https://ftp.ebi.ac.uk/pub/databases/gwas/summary_statistics/GCST010001-GCST011000/GCST010085 | 32317632 | 408,815 | European |
| Education (years) | https://ftp.ebi.ac.uk/pub/databases/gwas/summary_statistics/GCST008001-GCST009000/GCST008396/ | 23722424 | 126,559 | European |
| Intelligence | https://ftp.ebi.ac.uk/pub/databases/gwas/summary_statistics/GCST004001-GCST005000/GCST004364/ | 28530673 | 78,308 | European |
| Reaction time | https://ftp.ebi.ac.uk/pub/databases/gwas/summary_statistics/GCST006001-GCST007000/GCST006268/ | 29844566 | 330,069 | European |

**Table 4: Genetic correlation estimates between the nine DNEs**. We reported the genetic correlation ($g_c$) estimates, their standard errors, and the Z and P-values.

| DNE 1 | DNE 2 | $g_c$ mean | $g_c$ se | Z | P-value |
|---|---|---|---|---|---|
| AD1 | AD1 | 1 | ~0 | 216111.743 | 0 |
| AD1 | AD2 | -0.0475 | 0.0669 | -0.7101 | 0.4777 |
| AD1 | ASD1 | 0.0902 | 0.0626 | 1.4409 | 0.1496 |
| AD1 | ASD2 | 0.1809 | 0.0444 | 4.0748 | 4.61E-05 |
| AD1 | ASD3 | -0.1198 | 0.0554 | -2.1629 | 0.0305 |
| AD1 | LLD1 | -0.2622 | 0.0747 | -3.5084 | 0.0005 |
| AD1 | LLD2 | 0.1471 | 0.0685 | 2.1482 | 0.0317 |
| AD1 | SCZ1 | 0.3217 | 0.0492 | 6.5329 | 6.45E-11 |
| AD1 | SCZ2 | 0.0929 | 0.0597 | 1.5561 | 0.1197 |
| AD2 | AD1 | -0.0475 | 0.0669 | -0.7101 | 0.4777 |
| AD2 | AD2 | 1 | ~0 | 82056060.9 | 0 |
| AD2 | ASD1 | 0.1808 | 0.0583 | 3.1016 | 0.0019 |
| AD2 | ASD2 | -0.3257 | 0.0447 | -7.2933 | 3.02E-13 |
| AD2 | ASD3 | 0.244 | 0.0575 | 4.2471 | 2.17E-05 |
| AD2 | LLD1 | 0.1326 | 0.0831 | 1.5951 | 0.1107 |
| AD2 | LLD2 | -0.0751 | 0.0822 | -0.914 | 0.3607 |
| AD2 | SCZ1 | -0.1173 | 0.0644 | -1.8229 | 0.0683 |
| AD2 | SCZ2 | 0.009 | 0.0656 | 0.1371 | 0.8909 |
| ASD1 | AD1 | 0.0902 | 0.0626 | 1.4409 | 0.1496 |
| ASD1 | AD2 | 0.1808 | 0.0583 | 3.1016 | 0.0019 |
| ASD1 | ASD1 | 1 | ~0 | 71678.6548 | 0 |
| ASD1 | ASD2 | -0.5499 | 0.0408 | -13.4864 | 1.88E-41 |
| ASD1 | ASD3 | 0.2088 | 0.0543 | 3.8425 | 0.0001 |
| ASD1 | LLD1 | 0.1568 | 0.074 | 2.1191 | 0.0341 |
| ASD1 | LLD2 | 0.1464 | 0.074 | 1.9788 | 0.0478 |
| ASD1 | SCZ1 | 0.0024 | 0.06 | 0.0397 | 0.9683 |
| ASD1 | SCZ2 | 0.0147 | 0.0694 | 0.2116 | 0.8324 |
| ASD2 | AD1 | 0.1809 | 0.0444 | 4.0748 | 4.61E-05 |
| ASD2 | AD2 | -0.3257 | 0.0447 | -7.2933 | 3.02E-13 |
| ASD2 | ASD1 | -0.5499 | 0.0408 | -13.4864 | 1.88E-41 |
| ASD2 | ASD2 | 1 | ~0 | 398121.682 | 0 |
| ASD2 | ASD3 | -0.5374 | 0.0393 | -13.6754 | 1.42E-42 |
| ASD2 | LLD1 | -0.4037 | 0.064 | -6.3038 | 2.90E-10 |
| ASD2 | LLD2 | 0.2864 | 0.0671 | 4.2705 | 1.95E-05 |
| ASD2 | SCZ1 | 0.5672 | 0.0429 | 13.2214 | 6.60E-40 |
| ASD2 | SCZ2 | 0.0486 | 0.0527 | 0.9223 | 0.3564 |
| ASD3 | AD1 | -0.1198 | 0.0554 | -2.1629 | 0.0305 |
| ASD3 | AD2 | 0.244 | 0.0575 | 4.2471 | 2.17E-05 |
| ASD3 | ASD1 | 0.2088 | 0.0543 | 3.8425 | 0.0001 |
| ASD3 | ASD2 | -0.5374 | 0.0393 | -13.6754 | 1.42E-42 |
| ASD3 | ASD3 | 1 | ~0 | 677630000 | 0 |
| ASD3 | LLD1 | 0.5328 | 0.0606 | 8.7932 | 1.45E-18 |
| ASD3 | LLD2 | -0.3544 | 0.0716 | -4.9494 | 7.44E-07 |
| ASD3 | SCZ1 | -0.5117 | 0.0515 | -9.9268 | 3.18E-23 |
| ASD3 | SCZ2 | 0.3387 | 0.0536 | 6.3199 | 2.62E-10 |
| LLD1 | AD1 | -0.2622 | 0.0747 | -3.5084 | 0.0005 |
| LLD1 | AD2 | 0.1326 | 0.0831 | 1.5951 | 0.1107 |

| | | | | | |
|---|---|---|---|---|---|
| LLD1 | ASD1 | 0.1568 | 0.074 | 2.1191 | 0.0341 |
| LLD1 | ASD2 | -0.4037 | 0.064 | -6.3038 | 2.90E-10 |
| LLD1 | ASD3 | 0.5328 | 0.0606 | 8.7932 | 1.45E-18 |
| LLD1 | LLD1 | 1 | ~0 | 203600.738 | 0 |
| LLD1 | LLD2 | -0.5191 | 0.087 | -5.9652 | 2.44E-09 |
| LLD1 | SCZ1 | -0.4472 | 0.0778 | -5.7515 | 8.85E-09 |
| LLD1 | SCZ2 | 0.1616 | 0.081 | 1.9966 | 0.0459 |
| LLD2 | AD1 | 0.1471 | 0.0685 | 2.1482 | 0.0317 |
| LLD2 | AD2 | -0.0751 | 0.0822 | -0.914 | 0.3607 |
| LLD2 | ASD1 | 0.1464 | 0.074 | 1.9788 | 0.0478 |
| LLD2 | ASD2 | 0.2864 | 0.0671 | 4.2705 | 1.95E-05 |
| LLD2 | ASD3 | -0.3544 | 0.0716 | -4.9494 | 7.44E-07 |
| LLD2 | LLD1 | -0.5191 | 0.087 | -5.9652 | 2.44E-09 |
| LLD2 | LLD2 | 1 | ~0 | 1719937.56 | 0 |
| LLD2 | SCZ1 | 0.5205 | 0.0742 | 7.0159 | 2.29E-12 |
| LLD2 | SCZ2 | -0.1326 | 0.0744 | -1.7807 | 0.075 |
| SCZ1 | AD1 | 0.3217 | 0.0492 | 6.5329 | 6.45E-11 |
| SCZ1 | AD2 | -0.1173 | 0.0644 | -1.8229 | 0.0683 |
| SCZ1 | ASD1 | 0.0024 | 0.06 | 0.0397 | 0.9683 |
| SCZ1 | ASD2 | 0.5672 | 0.0429 | 13.2214 | 6.60E-40 |
| SCZ1 | ASD3 | -0.5117 | 0.0515 | -9.9268 | 3.18E-23 |
| SCZ1 | LLD1 | -0.4472 | 0.0778 | -5.7515 | 8.85E-09 |
| SCZ1 | LLD2 | 0.5205 | 0.0742 | 7.0159 | 2.29E-12 |
| SCZ1 | SCZ1 | 1 | ~0 | 245033.804 | 0 |
| SCZ1 | SCZ2 | 0.172 | 0.0608 | 2.8277 | 0.0047 |
| SCZ2 | AD1 | 0.0929 | 0.0597 | 1.5561 | 0.1197 |
| SCZ2 | AD2 | 0.009 | 0.0656 | 0.1371 | 0.8909 |
| SCZ2 | ASD1 | 0.0147 | 0.0694 | 0.2116 | 0.8324 |
| SCZ2 | ASD2 | 0.0486 | 0.0527 | 0.9223 | 0.3564 |
| SCZ2 | ASD3 | 0.3387 | 0.0536 | 6.3199 | 2.62E-10 |
| SCZ2 | LLD1 | 0.1616 | 0.081 | 1.9966 | 0.0459 |
| SCZ2 | LLD2 | -0.1326 | 0.0744 | -1.7807 | 0.075 |
| SCZ2 | SCZ1 | 0.172 | 0.0608 | 2.8277 | 0.0047 |
| SCZ2 | SCZ2 | 1 | ~0 | 332378.501 | 0 |

**Table 5: Genetic correlation estimates between the nine DNEs and the nine BAGs**. We reported the genetic correlation ($g_c$) estimates, their standard errors, and the Z and P-values.

| DNE | BAG | $g_c$ mean | $g_c$ se | Z | P-value |
|---|---|---|---|---|---|
| AD1 | Brain age gap | 0.2329 | 0.0491 | 4.7385 | 2.15E-06 |
| AD1 | Cardiovascular age gap | -0.0119 | 0.0457 | -0.2604 | 0.7946 |
| AD1 | Eye age gap | 0.0492 | 0.0803 | 0.6136 | 0.5395 |
| AD1 | Hepatic age gap | 0.0356 | 0.0604 | 0.5895 | 0.5555 |
| AD1 | Immune age gap | 0.0268 | 0.0724 | 0.3698 | 0.7115 |
| AD1 | Metabolic age gap | 0.0794 | 0.0556 | 1.4279 | 0.1533 |
| AD1 | Musculoskeletal age gap | 0.0677 | 0.0479 | 1.4152 | 0.157 |
| AD1 | Pulmonary age gap | 0.0573 | 0.0418 | 1.372 | 0.1701 |
| AD1 | Renal age gap | -0.0612 | 0.0402 | -1.522 | 0.128 |
| AD2 | Brain age gap | -0.021 | 0.0609 | -0.3442 | 0.7307 |
| AD2 | Cardiovascular age gap | -0.0551 | 0.0489 | -1.1284 | 0.2591 |
| AD2 | Eye age gap | 0.1729 | 0.0667 | 2.5908 | 0.0096 |
| AD2 | Hepatic age gap | -0.013 | 0.0541 | -0.2409 | 0.8096 |
| AD2 | Immune age gap | -0.036 | 0.0642 | -0.5606 | 0.5751 |
| AD2 | Metabolic age gap | -0.009 | 0.0517 | -0.1743 | 0.8616 |
| AD2 | Musculoskeletal age gap | 0.0657 | 0.0486 | 1.3537 | 0.1758 |
| AD2 | Pulmonary age gap | 0.061 | 0.0437 | 1.3976 | 0.1622 |
| AD2 | Renal age gap | 0.0166 | 0.0433 | 0.3825 | 0.7021 |
| ASD1 | Brain age gap | 0.4414 | 0.0504 | 8.7536 | 2.07E-18 |
| ASD1 | Cardiovascular age gap | -0.0596 | 0.0475 | -1.2538 | 0.2099 |
| ASD1 | Eye age gap | 0.194 | 0.0728 | 2.6635 | 0.0077 |
| ASD1 | Hepatic age gap | 0.0822 | 0.0595 | 1.3821 | 0.1669 |
| ASD1 | Immune age gap | 0.0053 | 0.0654 | 0.0817 | 0.9349 |
| ASD1 | Metabolic age gap | 0.0399 | 0.0521 | 0.7655 | 0.444 |
| ASD1 | Musculoskeletal age gap | 0.0839 | 0.0545 | 1.5405 | 0.1234 |
| ASD1 | Pulmonary age gap | -0.0255 | 0.0432 | -0.5903 | 0.555 |
| ASD1 | Renal age gap | 0.0689 | 0.0444 | 1.5518 | 0.1207 |
| ASD2 | Brain age gap | -0.1225 | 0.0674 | -1.8189 | 0.0689 |
| ASD2 | Cardiovascular age gap | 0.045 | 0.0414 | 1.0848 | 0.278 |
| ASD2 | Eye age gap | -0.1338 | 0.0479 | -2.7948 | 0.0052 |
| ASD2 | Hepatic age gap | -0.0466 | 0.0465 | -1.002 | 0.3163 |
| ASD2 | Immune age gap | -0.0202 | 0.0532 | -0.3798 | 0.7041 |
| ASD2 | Metabolic age gap | -0.0081 | 0.0433 | -0.1869 | 0.8517 |
| ASD2 | Musculoskeletal age gap | -0.0199 | 0.038 | -0.5252 | 0.5994 |
| ASD2 | Pulmonary age gap | 0.0238 | 0.0413 | 0.5772 | 0.5638 |
| ASD2 | Renal age gap | -0.0313 | 0.0373 | -0.839 | 0.4015 |
| ASD3 | Brain age gap | -0.0376 | 0.0647 | -0.5811 | 0.5612 |
| ASD3 | Cardiovascular age gap | 0.0114 | 0.0463 | 0.246 | 0.8057 |
| ASD3 | Eye age gap | -0.0904 | 0.052 | -1.737 | 0.0824 |
| ASD3 | Hepatic age gap | -0.0139 | 0.0558 | -0.2499 | 0.8027 |
| ASD3 | Immune age gap | 0.0121 | 0.0501 | 0.2416 | 0.8091 |
| ASD3 | Metabolic age gap | 0.0128 | 0.0493 | 0.2592 | 0.7954 |
| ASD3 | Musculoskeletal age gap | 0.0631 | 0.0498 | 1.2654 | 0.2057 |
| ASD3 | Pulmonary age gap | -0.0131 | 0.0477 | -0.2741 | 0.784 |
| ASD3 | Renal age gap | 0.0006 | 0.0461 | 0.0122 | 0.9903 |
| LLD1 | Brain age gap | -0.0928 | 0.0822 | -1.1283 | 0.2592 |
| LLD1 | Cardiovascular age gap | -0.0522 | 0.0714 | -0.7311 | 0.4647 |

| | | | | | |
|---|---|---|---|---|---|
| LLD1 | Eye age gap | -0.0931 | 0.0782 | -1.1908 | 0.2337 |
| LLD1 | Hepatic age gap | -0.0803 | 0.0743 | -1.0805 | 0.2799 |
| LLD1 | Immune age gap | 0.1297 | 0.0728 | 1.7826 | 0.0747 |
| LLD1 | Metabolic age gap | -0.1394 | 0.0696 | -2.0032 | 0.0452 |
| LLD1 | Musculoskeletal age gap | -0.1013 | 0.0686 | -1.4771 | 0.1396 |
| LLD1 | Pulmonary age gap | -0.1378 | 0.0668 | -2.0621 | 0.0392 |
| LLD1 | Renal age gap | 0.0119 | 0.0628 | 0.1902 | 0.8491 |
| LLD2 | Brain age gap | 0.2433 | 0.0779 | 3.1228 | 0.0018 |
| LLD2 | Cardiovascular age gap | 0.0065 | 0.0641 | 0.1012 | 0.9194 |
| LLD2 | Eye age gap | 0.0818 | 0.0713 | 1.1466 | 0.2515 |
| LLD2 | Hepatic age gap | 0.0889 | 0.0721 | 1.2328 | 0.2176 |
| LLD2 | Immune age gap | 0.0181 | 0.0684 | 0.2644 | 0.7915 |
| LLD2 | Metabolic age gap | 0.0679 | 0.0704 | 0.9637 | 0.3352 |
| LLD2 | Musculoskeletal age gap | 0.0475 | 0.0663 | 0.7176 | 0.473 |
| LLD2 | Pulmonary age gap | 0.016 | 0.0654 | 0.245 | 0.8064 |
| LLD2 | Renal age gap | -0.0297 | 0.0639 | -0.4652 | 0.6418 |
| SCZ1 | Brain age gap | 0.2603 | 0.0587 | 4.4319 | 9.34E-06 |
| SCZ1 | Cardiovascular age gap | -0.0058 | 0.0512 | -0.1128 | 0.9102 |
| SCZ1 | Eye age gap | 0.0094 | 0.0507 | 0.1855 | 0.8528 |
| SCZ1 | Hepatic age gap | 0.0746 | 0.0612 | 1.218 | 0.2232 |
| SCZ1 | Immune age gap | 0.02 | 0.0568 | 0.3522 | 0.7247 |
| SCZ1 | Metabolic age gap | 0.096 | 0.0528 | 1.8195 | 0.0688 |
| SCZ1 | Musculoskeletal age gap | 0.0802 | 0.0532 | 1.5065 | 0.1319 |
| SCZ1 | Pulmonary age gap | 0.0389 | 0.052 | 0.7488 | 0.454 |
| SCZ1 | Renal age gap | 0.0049 | 0.0493 | 0.0999 | 0.9204 |
| SCZ2 | Brain age gap | 0.0884 | 0.0623 | 1.4197 | 0.1557 |
| SCZ2 | Cardiovascular age gap | 0.0652 | 0.0502 | 1.3004 | 0.1935 |
| SCZ2 | Eye age gap | -0.0667 | 0.0713 | -0.9354 | 0.3496 |
| SCZ2 | Hepatic age gap | -0.0115 | 0.0641 | -0.1796 | 0.8574 |
| SCZ2 | Immune age gap | 0.0472 | 0.0644 | 0.7329 | 0.4636 |
| SCZ2 | Metabolic age gap | 0.1295 | 0.0551 | 2.3518 | 0.0187 |
| SCZ2 | Musculoskeletal age gap | 0.0518 | 0.0527 | 0.9825 | 0.3258 |
| SCZ2 | Pulmonary age gap | 0.0207 | 0.0454 | 0.4566 | 0.648 |
| SCZ2 | Renal age gap | -0.0837 | 0.0457 | -1.8312 | 0.0671 |

**Table 6: Genetic correlation estimates between the nine DNEs and the six neurodegenerative and neuropsychiatric disorders from PGC (a) and four lifestyle factors and cognitive scores (b)**. We reported the genetic correlation ($g_c$) estimates, their standard errors, and the Z and P-values.

**a**:

| DNE | PGC | $g_c$ mean | $g_c$ se | Z | P-value |
|---|---|---|---|---|---|
| AD1 | AD | -0.0672 | 0.1027 | -0.6541 | 0.513 |
| AD1 | ADHD | 0.0236 | 0.0396 | 0.5956 | 0.5515 |
| AD1 | ASD | 0.0433 | 0.0655 | 0.6603 | 0.5091 |
| AD1 | BIP | -0.0847 | 0.0404 | -2.0979 | 0.0359 |
| AD1 | OCD | -0.0076 | 0.0768 | -0.0987 | 0.9213 |
| AD1 | SCZ | 0.0071 | 0.0326 | 0.2185 | 0.8271 |
| AD2 | AD | 0.2244 | 0.1192 | 1.8821 | 0.0498 |
| AD2 | ADHD | -0.0365 | 0.0412 | -0.8863 | 0.3754 |
| AD2 | ASD | -0.0392 | 0.0602 | -0.6507 | 0.5152 |
| AD2 | BIP | -0.0219 | 0.0417 | -0.5264 | 0.5986 |
| AD2 | OCD | -0.0822 | 0.0817 | -1.0055 | 0.3147 |
| AD2 | SCZ | -0.0161 | 0.036 | -0.4461 | 0.6555 |
| ASD1 | AD | 0.1227 | 0.1248 | 0.9833 | 0.3255 |
| ASD1 | ADHD | -0.0385 | 0.0449 | -0.8577 | 0.3911 |
| ASD1 | ASD | 0.0417 | 0.0676 | 0.6164 | 0.5376 |
| ASD1 | BIP | -0.0066 | 0.0433 | -0.1532 | 0.8783 |
| ASD1 | OCD | -0.0146 | 0.0857 | -0.1707 | 0.8644 |
| ASD1 | SCZ | 0.0066 | 0.0364 | 0.1808 | 0.8565 |
| ASD2 | AD | -0.0315 | 0.0849 | -0.3707 | 0.7109 |
| ASD2 | ADHD | -0.0112 | 0.034 | -0.3286 | 0.7424 |
| ASD2 | ASD | -0.067 | 0.0459 | -1.4578 | 0.1449 |
| ASD2 | BIP | -0.0553 | 0.0368 | -1.5013 | 0.1333 |
| ASD2 | OCD | 0.0245 | 0.0804 | 0.305 | 0.7603 |
| ASD2 | SCZ | -0.0477 | 0.0372 | -1.2801 | 0.2005 |
| ASD3 | AD | 0.0272 | 0.0886 | 0.3074 | 0.7585 |
| ASD3 | ADHD | -0.0232 | 0.037 | -0.6257 | 0.5315 |
| ASD3 | ASD | 0.0678 | 0.0534 | 1.2703 | 0.204 |
| ASD3 | BIP | 0.0933 | 0.0463 | 2.0137 | 0.044 |
| ASD3 | OCD | 0.0824 | 0.0801 | 1.029 | 0.3035 |
| ASD3 | SCZ | 0.0375 | 0.0448 | 0.8368 | 0.4027 |
| LLD1 | AD | 0.0545 | 0.13 | 0.4195 | 0.6749 |
| LLD1 | ADHD | 0.0097 | 0.05 | 0.1949 | 0.8455 |
| LLD1 | ASD | 0.0672 | 0.0707 | 0.9498 | 0.3422 |
| LLD1 | BIP | 0.027 | 0.0526 | 0.5131 | 0.6079 |
| LLD1 | OCD | 0.0133 | 0.1253 | 0.1064 | 0.9152 |
| LLD1 | SCZ | -0.0037 | 0.0579 | -0.0643 | 0.9487 |
| LLD2 | AD | 0.1553 | 0.1516 | 1.0243 | 0.3057 |
| LLD2 | ADHD | -0.0424 | 0.0467 | -0.9077 | 0.364 |
| LLD2 | ASD | 0.0905 | 0.0625 | 1.4462 | 0.1481 |
| LLD2 | BIP | -0.0203 | 0.0594 | -0.3419 | 0.7324 |
| LLD2 | OCD | 0.0193 | 0.112 | 0.1724 | 0.8631 |
| LLD2 | SCZ | -0.0004 | 0.0623 | -0.007 | 0.9944 |
| SCZ1 | AD | 0.229 | 0.1398 | 1.6381 | 0.1014 |
| SCZ1 | ADHD | 0.0621 | 0.0421 | 1.4735 | 0.1406 |
| SCZ1 | ASD | -0.0101 | 0.0536 | -0.1882 | 0.8507 |

| | | | | | |
|---|---|---|---|---|---|
| SCZ1 | BIP | -0.0498 | 0.0467 | -1.066 | 0.2864 |
| SCZ1 | OCD | -0.0222 | 0.093 | -0.2386 | 0.8114 |
| SCZ1 | SCZ | 0.047 | 0.042 | 1.1179 | 0.2636 |
| SCZ2 | AD | 0.0101 | 0.1161 | 0.0867 | 0.9309 |
| SCZ2 | ADHD | 0.0227 | 0.04 | 0.5686 | 0.5696 |
| SCZ2 | ASD | 0.0402 | 0.0661 | 0.609 | 0.5425 |
| SCZ2 | BIP | 0.0727 | 0.0409 | 1.7764 | 0.0757 |
| SCZ2 | OCD | 0.0336 | 0.1002 | 0.3349 | 0.7377 |
| SCZ2 | SCZ | -0.001 | 0.0369 | -0.0284 | 0.9773 |

**b**:

| DNE | Lifestyle/cognition | $g_c$ mean | $g_c$ std | Z | P-value |
|---|---|---|---|---|---|
| AD1 | Computer | -0.0979 | 0.1372 | -0.7131 | 0.4758 |
| AD1 | Education | 0.0006 | 0.0498 | 0.0121 | 0.9903 |
| AD1 | Intellengence | -0.0051 | 0.0496 | -0.1031 | 0.9179 |
| AD1 | Reaction_time | 0.1416 | 0.1176 | 1.2046 | 0.2284 |
| AD2 | Computer | -0.0853 | 0.1909 | -0.4467 | 0.6551 |
| AD2 | Education | 0.0065 | 0.0491 | 0.132 | 0.895 |
| AD2 | Intellengence | -0.0212 | 0.0524 | -0.4051 | 0.6854 |
| AD2 | Reaction_time | -0.0962 | 0.163 | -0.5899 | 0.5553 |
| ASD1 | Computer | -0.0372 | 0.1197 | -0.311 | 0.7558 |
| ASD1 | Education | 0.0894 | 0.0545 | 1.6405 | 0.1009 |
| ASD1 | Intellengence | 0.0247 | 0.0481 | 0.5132 | 0.6078 |
| ASD1 | Reaction_time | 0.3551 | 0.1509 | 2.353 | 0.0186 |
| ASD2 | Computer | -0.0415 | 0.1115 | -0.3718 | 0.71 |
| ASD2 | Education | -0.0166 | 0.0456 | -0.3646 | 0.7154 |
| ASD2 | Intellengence | 0.0137 | 0.0454 | 0.3019 | 0.7627 |
| ASD2 | Reaction_time | -0.1248 | 0.1089 | -1.146 | 0.2518 |
| ASD3 | Computer | 0.0564 | 0.1248 | 0.4521 | 0.6512 |
| ASD3 | Education | 0.0437 | 0.0485 | 0.9012 | 0.3675 |
| ASD3 | Intellengence | 0.0661 | 0.0442 | 1.4967 | 0.1345 |
| ASD3 | Reaction_time | 0.1764 | 0.1265 | 1.3948 | 0.1631 |
| LLD1 | Computer | 0.1077 | 0.1652 | 0.6521 | 0.5143 |
| LLD1 | Education | 0.0534 | 0.0633 | 0.8435 | 0.399 |
| LLD1 | Intellengence | 0.0368 | 0.0652 | 0.5637 | 0.5729 |
| LLD1 | Reaction_time | -0.0211 | 0.1468 | -0.1435 | 0.8859 |
| LLD2 | Computer | -0.1827 | 0.1434 | -1.2745 | 0.2025 |
| LLD2 | Education | 0.0733 | 0.0668 | 1.0977 | 0.2723 |
| LLD2 | Intellengence | 0.0464 | 0.0628 | 0.7385 | 0.4602 |
| LLD2 | Reaction_time | 0.0266 | 0.1522 | 0.175 | 0.8611 |
| SCZ1 | Computer | -0.1565 | 0.1289 | -1.2143 | 0.2246 |
| SCZ1 | Education | -0.0836 | 0.0554 | -1.5095 | 0.1312 |
| SCZ1 | Intellengence | -0.0797 | 0.0491 | -1.6224 | 0.1047 |
| SCZ1 | Reaction_time | -0.1326 | 0.1499 | -0.8849 | 0.3762 |
| SCZ2 | Computer | -0.0562 | 0.2184 | -0.2572 | 0.7971 |
| SCZ2 | Education | -0.0656 | 0.0578 | -1.1354 | 0.2562 |
| SCZ2 | Intellengence | -0.0569 | 0.0526 | -1.0822 | 0.2792 |
| SCZ2 | Reaction_time | 0.0806 | 0.1318 | 0.6114 | 0.5409 |

**Table 7: Incremental $R^2$ of the nine DNEs to predict the 14 disease categories**. We reported the incremental $R^2$, P-value, $\beta$, and $\beta$ SE between each null and alternative model.

| DNE | Disease | Incremental $R^2$ | P | BETA | SE |
|---|---|---|---|---|---|
| AD1 | mental, behavioral disorder diagnosis | 0.00237542 | 2.94E-08 | 0.13163489 | 0.0237199 |
| AD2 | mental, behavioral disorder diagnosis | 0.00085888 | 0.0008553 | 0.08425787 | 0.02526249 |
| ASD1 | mental, behavioral disorder diagnosis | 0.00393483 | 9.56E-13 | 0.03738796 | 0.00523183 |
| ASD2 | mental, behavioral disorder diagnosis | 5.73E-07 | 0.93134456 | -0.0003819 | 0.00443215 |
| ASD3 | mental, behavioral disorder diagnosis | 0.0002156 | 0.09481276 | -0.0107486 | 0.00643358 |
| LLD1 | mental, behavioral disorder diagnosis | 0.00097948 | 0.00036995 | -0.00886 | 0.00248743 |
| LLD2 | mental, behavioral disorder diagnosis | 0.00079657 | 0.00132269 | 0.01111768 | 0.00346135 |
| SCZ1 | mental, behavioral disorder diagnosis | 0.00516644 | 2.90E-16 | 0.02019967 | 0.00246578 |
| SCZ2 | mental, behavioral disorder diagnosis | 0.00146472 | 1.34E-05 | 0.00952533 | 0.00218649 |
| ALL | mental, behavioral disorder diagnosis | 0.010108 | 1.74E-05 | NA | NA |
| AD1 | nerve system diagnosis | 0.00174659 | 4.72E-07 | 0.11738861 | 0.02328798 |
| AD2 | nerve system diagnosis | 0.00028308 | 0.04255513 | 0.05012132 | 0.024711 |
| ASD1 | nerve system diagnosis | 0.00209247 | 3.51E-08 | 0.02820525 | 0.00511151 |
| ASD2 | nerve system diagnosis | 2.64E-05 | 0.53560537 | -0.0026973 | 0.00435412 |
| ASD3 | nerve system diagnosis | 6.52E-05 | 0.33052015 | -0.0061449 | 0.00631466 |
| LLD1 | nerve system diagnosis | 0.00068081 | 0.00166028 | -0.007672 | 0.00243871 |
| LLD2 | nerve system diagnosis | 1.09E-05 | 0.69044837 | 0.00134605 | 0.00337984 |
| SCZ1 | nerve system diagnosis | 0.00301593 | 3.60E-11 | 0.01621908 | 0.00244751 |
| SCZ2 | nerve system diagnosis | 0.00064412 | 0.0022194 | 0.00657296 | 0.00214806 |
| ALL | nerve system diagnosis | 0.00628288 | 1.33E-05 | NA | NA |
| AD1 | musculoskeletal system diagnosis | 0.00043802 | 0.00108777 | 0.05706111 | 0.0174642 |
| AD2 | musculoskeletal system diagnosis | 0.00010102 | 0.11667892 | 0.02930294 | 0.01867687 |
| ASD1 | musculoskeletal system diagnosis | 0.00034275 | 0.00385563 | 0.01118673 | 0.00387068 |
| ASD2 | musculoskeletal system diagnosis | 3.93E-07 | 0.92206973 | 0.00032089 | 0.00328009 |
| ASD3 | musculoskeletal system diagnosis | 6.18E-05 | 0.21985805 | -0.0057587 | 0.00469349 |
| LLD1 | musculoskeletal system diagnosis | 0.00029832 | 0.00701853 | -0.0049747 | 0.00184504 |
| LLD2 | musculoskeletal system diagnosis | 2.06E-05 | 0.4785934 | 0.00182184 | 0.00257111 |
| SCZ1 | musculoskeletal system diagnosis | 0.00090987 | 2.50E-06 | 0.00871233 | 0.00184982 |
| SCZ2 | musculoskeletal system diagnosis | 0.0003082 | 0.00613966 | 0.00442347 | 0.00161408 |
| ALL | musculoskeletal system diagnosis | 0.0017996 | 0.003754 | NA | NA |

| | | | | | |
|---|---|---|---|---|---|
| AD1 | respiratory system diagnosis | 0.00070687 | 0.00029667 | 0.07865535 | 0.02173289 |
| AD2 | respiratory system diagnosis | 0.0001999 | 0.05433952 | 0.04453486 | 0.02314344 |
| ASD1 | respiratory system diagnosis | 0.00081394 | 0.00010337 | 0.0186489 | 0.00480176 |
| ASD2 | respiratory system diagnosis | 1.13E-05 | 0.64762659 | -0.0018569 | 0.00406266 |
| ASD3 | respiratory system diagnosis | 6.59E-05 | 0.26917672 | -0.0064775 | 0.00586191 |
| LLD1 | respiratory system diagnosis | 0.00037747 | 0.00819313 | -0.0060533 | 0.00228908 |
| LLD2 | respiratory system diagnosis | 9.69E-05 | 0.18033324 | 0.00427643 | 0.00319183 |
| SCZ1 | respiratory system diagnosis | 0.00117879 | 2.98E-06 | 0.01069371 | 0.00228768 |
| SCZ2 | respiratory system diagnosis | 0.00016666 | 0.07894093 | 0.00350661 | 0.00199579 |
| ALL | respiratory system diagnosis | 0.00243291 | 0.00603 | NA | NA |
| AD1 | circular system diagnosis | 0.00094607 | 8.17E-07 | 0.07931326 | 0.01607884 |
| AD2 | circular system diagnosis | 0.00022776 | 0.01554087 | 0.04174621 | 0.01725237 |
| ASD1 | circular system diagnosis | 0.00064175 | 4.88E-05 | 0.0144134 | 0.00354811 |
| ASD2 | circular system diagnosis | 0.00010097 | 0.10718718 | 0.00487077 | 0.00302337 |
| ASD3 | circular system diagnosis | 0.00011245 | 0.08911677 | -0.0074193 | 0.0043639 |
| LLD1 | circular system diagnosis | 0.00079564 | 6.12E-06 | -0.007702 | 0.00170269 |
| LLD2 | circular system diagnosis | 2.64E-05 | 0.40989677 | 0.00194535 | 0.00236059 |
| SCZ1 | circular system diagnosis | 0.00192407 | 2.03E-12 | 0.01191283 | 0.00169291 |
| SCZ2 | circular system diagnosis | 0.00044959 | 0.00067551 | 0.00505463 | 0.00148671 |
| ALL | circular system diagnosis | 0.00356403 | 0.000203 | NA | NA |
| AD1 | eye diagnosis | 0.00064458 | 0.00098626 | 0.07076414 | 0.02147457 |
| AD2 | eye diagnosis | 1.10E-05 | 0.66635253 | 0.00983323 | 0.02280592 |
| ASD1 | eye diagnosis | 0.00143173 | 9.11E-07 | 0.02297203 | 0.00467624 |
| ASD2 | eye diagnosis | 0.00014656 | 0.11620726 | 0.00628472 | 0.00400043 |
| ASD3 | eye diagnosis | 0.00043907 | 0.00654861 | -0.0155847 | 0.00573083 |
| LLD1 | eye diagnosis | 0.00047845 | 0.00453561 | -0.0064098 | 0.00225788 |
| LLD2 | eye diagnosis | 0.00013452 | 0.13232354 | 0.00466842 | 0.00310172 |
| SCZ1 | eye diagnosis | 0.001841 | 2.58E-08 | 0.01254589 | 0.00225184 |
| SCZ2 | eye diagnosis | 0.00046267 | 0.00525298 | 0.00547183 | 0.00196009 |
| ALL | eye diagnosis | 0.00383532 | 2.31E-5 | NA | NA |
| AD1 | ear diagnosis | 0.00048026 | 0.08319279 | 0.03553414 | 0.02050817 |
| AD2 | ear diagnosis | 0.001865 | 0.00064021 | 0.07408068 | 0.02169016 |
| ASD1 | ear diagnosis | 0.00072129 | 0.03374312 | 0.0096174 | 0.004529 |
| ASD2 | ear diagnosis | 0.00014624 | 0.33908348 | 0.00364804 | 0.00381579 |
| ASD3 | ear diagnosis | 0.00033791 | 0.14617111 | -0.008052 | 0.00554031 |
| LLD1 | ear diagnosis | 0.00078238 | 0.02702076 | -0.0047008 | 0.00212547 |
| LLD2 | ear diagnosis | 7.75E-06 | 0.82579589 | -0.0006504 | 0.00295487 |
| SCZ1 | ear diagnosis | 0.00186538 | 0.0006394 | 0.00733901 | 0.00214858 |
| SCZ2 | ear diagnosis | 0.00039387 | 0.11666754 | 0.002924 | 0.00186349 |
| ALL | ear diagnosis | 0.00576759 | 0.000457 | NA | NA |
| AD1 | blood and immune system diagnosis | 0.00046232 | 0.001881 | 0.06247271 | 0.02009431 |
| AD2 | blood and immune system diagnosis | 0.00037029 | 0.00540481 | 0.05992387 | 0.02153769 |
| ASD1 | blood and immune system diagnosis | 0.00051838 | 0.00099665 | 0.01474243 | 0.00447807 |

| | | | | | |
|---|---|---|---|---|---|
| ASD2 | blood and immune system diagnosis | 7.10E-06 | 0.70009125 | 0.00145876 | 0.00378696 |
| ASD3 | blood and immune system diagnosis | 4.20E-05 | 0.34866926 | -0.0050982 | 0.00543979 |
| LLD1 | blood and immune system diagnosis | 0.00077428 | 5.75E-05 | -0.0085956 | 0.00213617 |
| LLD2 | blood and immune system diagnosis | 0.0001557 | 0.07124808 | 0.00531391 | 0.00294558 |
| SCZ1 | blood and immune system diagnosis | 0.00088436 | 1.71E-05 | 0.00914335 | 0.00212607 |
| SCZ2 | blood and immune system diagnosis | 0.00013656 | 0.09114085 | 0.00315171 | 0.00186545 |
| ALL | blood and immune system diagnosis | 0.00216765 | 0.00515 | NA | NA |
| AD1 | endocrine nutritional metabolic disease diagnosis | 0.00170318 | 2.77E-09 | 0.11728821 | 0.01971713 |
| AD2 | endocrine nutritional metabolic disease diagnosis | 0.00023609 | 0.02687606 | 0.04646234 | 0.02099007 |
| ASD1 | endocrine nutritional metabolic disease diagnosis | 0.00202934 | 8.64E-11 | 0.02829016 | 0.00435639 |
| ASD2 | endocrine nutritional metabolic disease diagnosis | 5.95E-06 | 0.72523073 | -0.0013015 | 0.00370294 |
| ASD3 | endocrine nutritional metabolic disease diagnosis | 4.60E-05 | 0.32853428 | -0.005219 | 0.00534127 |
| LLD1 | endocrine nutritional metabolic disease diagnosis | 0.0006426 | 0.00026067 | -0.007638 | 0.0020912 |
| LLD2 | endocrine nutritional metabolic disease diagnosis | 0.00017299 | 0.05814821 | 0.00551014 | 0.00290814 |
| SCZ1 | endocrine nutritional metabolic disease diagnosis | 0.00348545 | 1.83E-17 | 0.0177244 | 0.00208154 |
| SCZ2 | endocrine nutritional metabolic disease diagnosis | 0.00067736 | 0.00017754 | 0.00681612 | 0.00181764 |
| ALL | endocrine nutritional metabolic disease diagnosis | 0.00613159 | 1.65E-8 | NA | NA |
| AD1 | digestive system diagnosis | 0.0003768 | 0.00098595 | 0.046342 | 0.0140647 |
| AD2 | digestive system diagnosis | 0.00017012 | 0.02685391 | 0.03327534 | 0.01503106 |
| ASD1 | digestive system diagnosis | 0.00028553 | 0.00413223 | 0.00898262 | 0.00313184 |
| ASD2 | digestive system diagnosis | 2.08E-06 | 0.80670262 | 0.00064384 | 0.00263132 |
| ASD3 | digestive system diagnosis | 5.02E-05 | 0.22919193 | -0.004561 | 0.00379302 |
| LLD1 | digestive system diagnosis | 0.00030248 | 0.00315939 | -0.004417 | 0.00149624 |
| LLD2 | digestive system diagnosis | 3.04E-06 | 0.7672152 | 0.00060941 | 0.00205864 |
| SCZ1 | digestive system diagnosis | 0.00061387 | 2.61E-05 | 0.00623438 | 0.00148229 |
| SCZ2 | digestive system diagnosis | 0.00022808 | 0.01037233 | 0.00333684 | 0.00130174 |
| ALL | digestive system diagnosis | 0.00151556 | 0.0178 | NA | NA |
| AD1 | genitourinary system diagnosis | 0.000224 | 0.02094975 | 0.04169411 | 0.01805642 |

| | | | | | |
|---|---|---|---|---|---|
| AD2 | genitourinary system diagnosis | 8.12E-05 | 0.16441193 | 0.02679599 | 0.01927153 |
| ASD1 | genitourinary system diagnosis | 0.00050929 | 0.00049865 | 0.01386977 | 0.00398311 |
| ASD2 | genitourinary system diagnosis | 1.00E-05 | 0.62541236 | 0.00164534 | 0.0033702 |
| ASD3 | genitourinary system diagnosis | 1.96E-05 | 0.49424717 | -0.0032869 | 0.00480832 |
| LLD1 | genitourinary system diagnosis | 0.00027778 | 0.01013494 | -0.0048879 | 0.00190083 |
| LLD2 | genitourinary system diagnosis | 2.96E-07 | 0.93311861 | -0.0002221 | 0.00264688 |
| SCZ1 | genitourinary system diagnosis | 0.00060353 | 0.00015069 | 0.00716381 | 0.0018898 |
| SCZ2 | genitourinary system diagnosis | 0.00018196 | 0.03743466 | 0.00343775 | 0.00165185 |
| ALL | genitourinary system diagnosis | 0.00149999 | 0.00529 | NA | NA |
| AD1 | infectious parasitic disease diagnosis | 0.00060453 | 0.00501325 | 0.0665503 | 0.02371018 |
| AD2 | infectious parasitic disease diagnosis | 0.00037672 | 0.02674529 | 0.05570015 | 0.02514052 |
| ASD1 | infectious parasitic disease diagnosis | 0.00118761 | 8.38E-05 | 0.02042624 | 0.00519109 |
| ASD2 | infectious parasitic disease diagnosis | 7.53E-05 | 0.32210343 | -0.0043873 | 0.00443079 |
| ASD3 | infectious parasitic disease diagnosis | 7.11E-05 | 0.33577691 | -0.0061356 | 0.006374 |
| LLD1 | infectious parasitic disease diagnosis | 0.00038981 | 0.02423576 | -0.0055779 | 0.00247499 |
| LLD2 | infectious parasitic disease diagnosis | 0.00021208 | 0.09648928 | 0.00571781 | 0.00343974 |
| SCZ1 | infectious parasitic disease diagnosis | 0.00102638 | 0.00025572 | 0.00910426 | 0.00248899 |
| SCZ2 | infectious parasitic disease diagnosis | 0.00024195 | 0.0758475 | 0.003846 | 0.00216615 |
| ALL | infectious parasitic disease diagnosis | 0.0027959 | 0.03817 | NA | NA |
| AD1 | neoplasms diagnosis | 0.00024321 | 0.01811767 | 0.04361325 | 0.01845328 |
| AD2 | neoplasms diagnosis | 8.44E-05 | 0.16378184 | 0.02746435 | 0.01972268 |
| ASD1 | neoplasms diagnosis | 0.00024474 | 0.01775832 | 0.00970248 | 0.00409239 |
| ASD2 | Neoplasms diagnosis | 3.15E-05 | 0.39479562 | 0.00293534 | 0.00344939 |
| ASD3 | neoplasms diagnosis | 6.78E-05 | 0.21201808 | -0.0061899 | 0.00495953 |
| LLD1 | neoplasms diagnosis | 0.00039354 | 0.00264638 | -0.0058908 | 0.00195933 |
| LLD2 | neoplasms diagnosis | 5.40E-05 | 0.26559564 | 0.00302063 | 0.00271322 |
| SCZ1 | neoplasms diagnosis | 0.00071621 | 5.01E-05 | 0.0079017 | 0.00194794 |
| SCZ2 | neoplasms diagnosis | 0.00010107 | 0.12765485 | 0.00259783 | 0.00170518 |
| ALL | neoplasms diagnosis | 0.00124085 | 0.0324 | NA | NA |
| AD1 | skin system diagnosis | 0.00020354 | 0.0693237 | 0.04152431 | 0.0228598 |
| AD2 | skin system diagnosis | 0.00027384 | 0.03513816 | 0.05116606 | 0.02428366 |
| ASD1 | skin system diagnosis | 0.00033789 | 0.01927391 | 0.01182505 | 0.00505231 |
| ASD2 | skin system diagnosis | 6.23E-05 | 0.31487399 | 0.00427281 | 0.00425116 |
| ASD3 | skin system diagnosis | 5.40E-05 | 0.3494439 | -0.0057385 | 0.00613284 |
| LLD1 | skin system diagnosis | 0.00015087 | 0.11787319 | -0.0037373 | 0.00238973 |
| LLD2 | skin system diagnosis | 2.24E-05 | 0.5464881 | 0.0019931 | 0.00330504 |
| SCZ1 | skin system diagnosis | 0.00093169 | 0.00010191 | 0.00924265 | 0.0023776 |

| | | | | | |
|---|---|---|---|---|---|
| SCZ2 | skin system diagnosis | 0.00060612 | 0.0017224 | 0.00654178 | 0.00208663 |
| ALL | skin system diagnosis | 0.00194049 | 0.0082 | NA | NA |

**Table 8: Incremental $R^2$ of the nine PRSs to predict the 14 disease categories**. We reported the incremental $R^2$, P-value, $\beta$, and $\beta$ SE between each null and alternative model.

| DNE | Disease | Incremental $R^2$ | P | BETA | SE |
|---|---|---|---|---|---|
| AD1 | mental, behavioral disorder diagnosis | 0.00064961 | 0.0641007 | 1002.22394 | 541.157191 |
| AD2 | mental, behavioral disorder diagnosis | 8.58E-05 | 0.5011149 | -369.51219 | 549.216429 |
| ASD1 | mental, behavioral disorder diagnosis | 0.00022449 | 0.27642074 | -600.41844 | 551.57805 |
| ASD2 | mental, behavioral disorder diagnosis | 0.0004132 | 0.13977404 | 805.164113 | 545.159749 |
| ASD3 | mental, behavioral disorder diagnosis | 0.00031247 | 0.19911132 | 704.598659 | 548.622786 |
| LLD1 | mental, behavioral disorder diagnosis | 0.00061716 | 0.07113046 | 1003.80025 | 556.080907 |
| LLD2 | mental, behavioral disorder diagnosis | 0.0002309 | 0.2696651 | 607.047715 | 549.866926 |
| SCZ1 | mental, behavioral disorder diagnosis | 1.54E-06 | 0.9281793 | -48.781168 | 541.16156 |
| SCZ2 | mental, behavioral disorder diagnosis | 1.44E-05 | 0.78312993 | -153.71494 | 558.432645 |
| ALL | mental, behavioral disorder diagnosis | 0.00320626 | 0.0471 | NA | NA |
| AD1 | nerve system diagnosis | 0.00012982 | 0.37818039 | -469.10199 | 532.256402 |
| AD2 | nerve system diagnosis | 8.84E-06 | 0.81807889 | -125.14086 | 544.017862 |
| ASD1 | nerve system diagnosis | 0.00047378 | 0.09228039 | -918.66608 | 545.568786 |
| ASD2 | nerve system diagnosis | 0.00011352 | 0.40990846 | 438.566669 | 532.153289 |
| ASD3 | nerve system diagnosis | 0.00011748 | 0.40185596 | 458.295503 | 546.634352 |
| LLD1 | nerve system diagnosis | 0.00011958 | 0.39767626 | 462.314145 | 546.55861 |
| LLD2 | nerve system diagnosis | 8.59E-05 | 0.47356415 | 382.668535 | 533.889754 |
| SCZ1 | nerve system diagnosis | 2.31E-05 | 0.71018114 | -198.99193 | 535.446082 |
| SCZ2 | nerve system diagnosis | 1.39E-05 | 0.77311665 | -158.51624 | 549.794777 |
| ALL | nerve system diagnosis | 0.00144565 | 0.99121924 | NA | NA |
| AD1 | musculoskeletal system diagnosis | 0.00032126 | 0.07430323 | 705.910408 | 395.468313 |
| AD2 | musculoskeletal system diagnosis | 5.85E-05 | 0.44642918 | 306.586049 | 402.648802 |
| ASD1 | musculoskeletal system diagnosis | 3.03E-05 | 0.58346892 | 222.728268 | 406.17954 |
| ASD2 | musculoskeletal system diagnosis | 0.0001391 | 0.24024494 | 468.139954 | 398.596461 |
| ASD3 | musculoskeletal system diagnosis | 0.0003683 | 0.05600965 | 767.541718 | 401.588038 |
| LLD1 | musculoskeletal system diagnosis | 0.0004981 | 0.0262617 | 897.714485 | 403.871256 |
| LLD2 | musculoskeletal system diagnosis | 0.00025262 | 0.11350047 | 641.954968 | 405.570506 |
| SCZ1 | musculoskeletal system diagnosis | 0.00014305 | 0.23367679 | 476.429481 | 400.011321 |
| SCZ2 | musculoskeletal system diagnosis | 8.24E-05 | 0.36597573 | 369.092309 | 408.245671 |
| ALL | musculoskeletal system diagnosis | 0.00106871 | 0.98463353 | NA | NA |

| | | | | | |
|---|---|---|---|---|---|
| AD1 | respiratory system diagnosis | 3.58E-07 | 0.95859964 | 25.9027194 | 498.959488 |
| AD2 | respiratory system diagnosis | 0.00012268 | 0.33686049 | -485.98395 | 505.982348 |
| ASD1 | respiratory system diagnosis | 7.58E-05 | 0.45027386 | -379.38279 | 502.483277 |
| ASD2 | respiratory system diagnosis | 0.00027868 | 0.14775519 | 716.870208 | 495.170999 |
| ASD3 | respiratory system diagnosis | 0.0001 | 0.38588548 | 439.051137 | 506.299632 |
| LLD1 | respiratory system diagnosis | 7.08E-05 | 0.46554477 | 372.209321 | 510.014747 |
| LLD2 | respiratory system diagnosis | 4.02E-06 | 0.86196099 | 86.984747 | 500.236919 |
| SCZ1 | respiratory system diagnosis | 3.92E-06 | 0.86361192 | 86.0486396 | 500.903905 |
| SCZ2 | respiratory system diagnosis | 2.41E-06 | 0.89291482 | 68.9572743 | 512.223279 |
| ALL | respiratory system diagnosis | 0.00084301 | 0.99668619 | NA | NA |
| AD1 | circular system diagnosis | 6.95E-05 | 0.39355519 | 312.831994 | 366.642539 |
| AD2 | circular system diagnosis | 5.93E-06 | 0.8031248 | 93.8640218 | 376.49001 |
| ASD1 | circular system diagnosis | 2.91E-05 | 0.58088726 | -206.12577 | 373.34055 |
| ASD2 | circular system diagnosis | 0.0001807 | 0.16888152 | 507.755637 | 369.026621 |
| ASD3 | circular system diagnosis | 0.00028278 | 0.08523606 | 646.001466 | 375.299309 |
| LLD1 | circular system diagnosis | 0.00023365 | 0.11771428 | 583.459131 | 372.910196 |
| LLD2 | circular system diagnosis | 9.24E-05 | 0.32532533 | 365.720224 | 371.805009 |
| SCZ1 | circular system diagnosis | 5.08E-05 | 0.46558909 | 269.433018 | 369.233036 |
| SCZ2 | circular system diagnosis | 3.05E-06 | 0.85820061 | 67.2659646 | 376.479471 |
| ALL | circular system diagnosis | 0.00089966 | 0.99199868 | NA | NA |
| AD1 | eye diagnosis | 0.00025879 | 0.18269775 | 644.284394 | 483.446885 |
| AD2 | eye diagnosis | 0.00022827 | 0.21076704 | -618.19234 | 493.910587 |
| ASD1 | eye diagnosis | 3.79E-06 | 0.87181272 | 79.7061497 | 493.94914 |
| ASD2 | eye diagnosis | 0.00041161 | 0.09286303 | 812.293126 | 483.271089 |
| ASD3 | eye diagnosis | 6.95E-05 | 0.48999467 | 342.084976 | 495.508955 |
| LLD1 | eye diagnosis | 0.00047016 | 0.0724894 | 885.260819 | 492.788502 |
| LLD2 | eye diagnosis | 3.68E-07 | 0.9599381 | 24.8173899 | 494.036623 |
| SCZ1 | eye diagnosis | 0.00026382 | 0.17850672 | 658.611168 | 489.467756 |
| SCZ2 | eye diagnosis | 0.00025383 | 0.18694659 | 654.193647 | 495.655225 |
| ALL | eye diagnosis | 0.00161593 | 0.0407 | NA | NA |
| AD1 | ear diagnosis | 1.18E-06 | 0.95630261 | 25.3016667 | 461.722371 |
| AD2 | ear diagnosis | 1.92E-06 | 0.94436694 | 33.3348344 | 477.658502 |
| ASD1 | ear diagnosis | 9.03E-07 | 0.96180107 | -22.466759 | 469.059343 |
| ASD2 | ear diagnosis | 5.62E-06 | 0.90488044 | 55.1158338 | 461.188118 |
| ASD3 | ear diagnosis | 2.15E-05 | 0.81515058 | 111.663209 | 477.587586 |
| LLD1 | ear diagnosis | 6.94E-05 | 0.6745449 | -200.90695 | 478.396758 |
| LLD2 | ear diagnosis | 0.00032124 | 0.36632231 | -420.43454 | 465.33096 |
| SCZ1 | ear diagnosis | 0.00010179 | 0.61109025 | -233.14872 | 458.439857 |
| SCZ2 | ear diagnosis | 0.00031034 | 0.37458445 | -423.18102 | 476.529417 |
| ALL | ear diagnosis | 0.0011697 | 0.99999083 | NA | NA |
| AD1 | blood and immune system diagnosis | 0.00018737 | 0.20890925 | 579.970953 | 461.499957 |
| AD2 | blood and immune system diagnosis | 2.02E-05 | 0.68016882 | 192.909058 | 467.938136 |
| ASD1 | blood and immune system diagnosis | 0.00013414 | 0.28769645 | 502.647025 | 472.730224 |

| | | | | | |
|---|---|---|---|---|---|
| ASD2 | blood and immune system diagnosis | 0.00031393 | 0.10384963 | 745.069138 | 458.018381 |
| ASD3 | blood and immune system diagnosis | 0.00053559 | 0.03363234 | 996.357173 | 468.884593 |
| LLD1 | blood and immune system diagnosis | 0.00071889 | 0.01384368 | 1168.28845 | 474.521431 |
| LLD2 | blood and immune system diagnosis | 8.42E-06 | 0.78993023 | 123.761697 | 464.5476 |
| SCZ1 | blood and immune system diagnosis | 0.0002175 | 0.17578857 | 619.537735 | 457.561116 |
| SCZ2 | blood and immune system diagnosis | 0.00016311 | 0.24103624 | 554.785196 | 473.156528 |
| ALL | blood and immune system diagnosis | 0.00134203 | 0.97420569 | NA | NA |
| AD1 | endocrine nutritional metabolic disease diagnosis | 0.00026604 | 0.13425774 | 671.627555 | 448.430917 |
| AD2 | endocrine nutritional metabolic disease diagnosis | 7.89E-05 | 0.41487418 | 376.608031 | 461.866769 |
| ASD1 | endocrine nutritional metabolic disease diagnosis | 0.00013493 | 0.2862056 | -490.00265 | 459.414974 |
| ASD2 | endocrine nutritional metabolic disease diagnosis | 0.00021754 | 0.1756906 | 613.775322 | 453.202475 |
| ASD3 | endocrine nutritional metabolic disease diagnosis | 0.00018954 | 0.20622723 | 583.426632 | 461.517746 |
| LLD1 | endocrine nutritional metabolic disease diagnosis | 0.00028175 | 0.12329441 | 715.498022 | 464.213113 |
| LLD2 | endocrine nutritional metabolic disease diagnosis | 0.00042325 | 0.05891343 | 870.202252 | 460.617821 |
| SCZ1 | endocrine nutritional metabolic disease diagnosis | 0.0001602 | 0.24521434 | 532.02898 | 457.789904 |
| SCZ2 | endocrine nutritional metabolic disease diagnosis | 6.98E-05 | 0.44310247 | 357.254948 | 465.772066 |
| ALL | endocrine nutritional metabolic disease diagnosis | 0.00156547 | 0.01501 | NA | NA |
| AD1 | digestive system diagnosis | 3.48E-05 | 0.52394109 | 205.015732 | 321.691571 |
| AD2 | digestive system diagnosis | 1.36E-05 | 0.6901336 | 128.953796 | 323.447263 |
| ASD1 | digestive system diagnosis | 6.88E-06 | 0.77674165 | -92.438397 | 325.974354 |
| ASD2 | digestive system diagnosis | 2.13E-05 | 0.61763086 | 160.979059 | 322.456678 |
| ASD3 | digestive system diagnosis | 0.00020861 | 0.11849144 | 510.09124 | 326.714434 |
| LLD1 | digestive system diagnosis | 0.0002474 | 0.08911556 | 555.075958 | 326.466782 |
| LLD2 | digestive system diagnosis | 3.48E-05 | 0.52358285 | 208.670317 | 327.143573 |
| SCZ1 | digestive system diagnosis | 9.40E-06 | 0.7402962 | 107.2338 | 323.509209 |
| SCZ2 | digestive system diagnosis | 1.67E-06 | 0.88896812 | -45.990939 | 329.415677 |
| ALL | digestive system diagnosis | 0.00059561 | 0.99816066 | NA | NA |
| AD1 | genitourinary system diagnosis | 2.42E-05 | 0.62904948 | 197.134341 | 408.072549 |

| | | | | | |
|---|---|---|---|---|---|
| AD2 | genitourinary system diagnosis | 1.55E-06 | 0.90281412 | -50.787484 | 415.910268 |
| ASD1 | genitourinary system diagnosis | 4.77E-06 | 0.8301911 | 89.5073273 | 417.352611 |
| ASD2 | genitourinary system diagnosis | 0.00021722 | 0.14788697 | 588.689678 | 406.779499 |
| ASD3 | genitourinary system diagnosis | 0.00020057 | 0.16438796 | 578.155381 | 415.756544 |
| LLD1 | genitourinary system diagnosis | 0.00033077 | 0.0741569 | 741.582395 | 415.240109 |
| LLD2 | genitourinary system diagnosis | 0.00011428 | 0.29392077 | 433.729099 | 413.217642 |
| SCZ1 | genitourinary system diagnosis | 1.74E-05 | 0.68172858 | 168.562738 | 411.005681 |
| SCZ2 | genitourinary system diagnosis | 6.46E-05 | 0.43017865 | 334.49626 | 423.986254 |
| ALL | genitourinary system diagnosis | 0.00075426 | 0.99605949 | NA | NA |
| AD1 | infectious parasitic disease diagnosis | 0.00012504 | 0.41672454 | 438.365556 | 539.724217 |
| AD2 | infectious parasitic disease diagnosis | 1.45E-06 | 0.93037831 | -47.704374 | 545.975996 |
| ASD1 | infectious parasitic disease diagnosis | 1.93E-06 | 0.91972335 | -55.032836 | 546.021888 |
| ASD2 | infectious parasitic disease diagnosis | 0.00029874 | 0.20937165 | 676.383654 | 538.735929 |
| ASD3 | infectious parasitic disease diagnosis | 2.47E-05 | 0.71802971 | 196.768042 | 544.883732 |
| LLD1 | infectious parasitic disease diagnosis | 0.00058197 | 0.07976417 | 965.58153 | 550.97037 |
| LLD2 | infectious parasitic disease diagnosis | 0.00021798 | 0.28359279 | 578.273584 | 539.217861 |
| SCZ1 | infectious parasitic disease diagnosis | 3.16E-08 | 0.98969747 | -6.9676008 | 539.559839 |
| SCZ2 | infectious parasitic disease diagnosis | 5.03E-07 | 0.95891489 | 28.5143269 | 553.476197 |
| ALL | infectious parasitic disease diagnosis | 0.0013111 | 0.99696853 | NA | NA |
| AD1 | neoplasms diagnosis | 0.00020104 | 0.17126888 | 572.333976 | 418.285397 |
| AD2 | neoplasms diagnosis | 4.04E-05 | 0.53973881 | 262.71846 | 428.410684 |
| ASD1 | neoplasms diagnosis | 8.88E-05 | 0.36310323 | 393.482423 | 432.621551 |
| ASD2 | Neoplasms diagnosis | 0.00023631 | 0.13799223 | 625.996102 | 421.975866 |
| ASD3 | neoplasms diagnosis | 0.0004514 | 0.04035772 | 884.244689 | 431.24 |
| LLD1 | neoplasms diagnosis | 0.00060002 | 0.01809947 | 1022.49603 | 432.498186 |
| LLD2 | neoplasms diagnosis | 5.67E-05 | 0.46736839 | 311.884688 | 429.114895 |
| SCZ1 | neoplasms diagnosis | 0.00020427 | 0.16787178 | 577.592361 | 418.780355 |
| SCZ2 | neoplasms diagnosis | 0.00018107 | 0.19414879 | 561.13234 | 432.12741 |
| ALL | neoplasms diagnosis | 0.00105648 | 0.98685197 | NA | NA |
| AD1 | skin system diagnosis | 0.00018111 | 0.27120728 | 569.795946 | 517.799796 |
| AD2 | skin system diagnosis | 2.05E-05 | 0.71105439 | 194.826219 | 525.899195 |
| ASD1 | skin system diagnosis | 4.24E-06 | 0.86633766 | 89.0313404 | 528.936136 |
| ASD2 | skin system diagnosis | 0.00029347 | 0.16132298 | 725.034794 | 517.564377 |
| ASD3 | skin system diagnosis | 0.00018202 | 0.26999857 | 587.00734 | 532.096353 |
| LLD1 | skin system diagnosis | 3.60E-05 | 0.62379205 | 258.216555 | 526.41833 |
| LLD2 | skin system diagnosis | 0.0003434 | 0.12974866 | 792.93787 | 523.266134 |
| SCZ1 | skin system diagnosis | 0.00046878 | 0.07669251 | 906.684289 | 512.076915 |

| | | | | | |
|---|---|---|---|---|---|
| SCZ2 | skin system diagnosis | 0.00016546 | 0.29294279 | 556.74905 | 529.326224 |
| ALL | skin system diagnosis | 0.00088119 | 0.99775902 | NA | NA |

**Table 9: Prediction accuracy of the nine DNEs and PRSs to predict the 14 disease categories (a) and 8 cognitive scores (b)**. We reported the accuracy of each task using different sets of features for disease classification and Pearson's r for cognitive score prediction.

**a) Disease classification** (accuracy is presented below). Other evaluation metrics in both cross-validated and independent results are presented in **Supplementary eFile 22**.

| ICD-10 disease category | Age+sex+9PRS | Age+sex+9DNE | Age+sex+9DNE+9PRS | Age+sex+9DNE+9PRS+119IDP |
|---|---|---|---|---|
| blood and immune system diagnosis | 0.56632899 | 0.56777617 | 0.5726001 | 0.56253165 |
| circular system diagnosis | 0.61794501 | 0.62988423 | 0.62879884 | 0.59168242 |
| digestive system diagnosis | 0.58791049 | 0.58645743 | 0.59226969 | 0.49121212 |
| ear diagnosis | 0.58962693 | 0.60145587 | 0.58598726 | 0.6152381 |
| endocrine, nutritional, and metabolic disease diagnosis | 0.60173997 | 0.61430643 | 0.60898985 | 0.60020243 |
| eye diagnosis | 0.6373365 | 0.6510107 | 0.6450654 | 0.64383562 |
| genitourinary system diagnosis | 0.57487923 | 0.57487923 | 0.57487923 | 0.57473684 |
| infectious parasitic disease diagnosis | 0.53211009 | 0.57304164 | 0.56245589 | 0.56890199 |
| mental, behavioral disorder diagnosis | 0.51089248 | 0.55305692 | 0.57181307 | 0.56323529 |
| musculoskeletal system diagnosis | 0.59776321 | 0.59853452 | 0.60470497 | 0.54394492 |
| neoplasms diagnosis | 0.60051107 | 0.59582624 | 0.60647359 | 0.57117358 |
| nerve system diagnosis | 0.56425703 | 0.57563588 | 0.59772423 | 0.60098177 |
| respiratory system diagnosis | 0.55126792 | 0.5722161 | 0.56725469 | 0.56473988 |
| skin system diagnosis | 0.55808236 | 0.57467732 | 0.55500922 | 0.5374677 |

**b) Cognitive score prediction** (Pearson's *r* is presented below). Other evaluation metrics in both cross-validated and independent results are presented in **Supplementary eFile 23**.

| Cognitive score | Age+sex+9PRS | Age+sex+9DNE | Age+sex+9DNE+9PRS | Age+sex+9DNE+9PRS+119IDP |
|---|---|---|---|---|
| duration_to_complete_alphanumeric_path_trail_2_6350 | 0.25177398 | 0.2518425 | 0.24862606 | 0.25082623 |
| duration_to_complete_numeric_path_trail_1_6348 | 0.30790081 | 0.30813216 | 0.30811301 | 0.30697352 |
| fluid_intelligence_score_20016 | 0.21864076 | 0.22007454 | 0.22077979 | 0.24022209 |
| maximum_digits_remembered_correctly_4282 | 0.16682944 | 0.1753658 | 0.17018593 | 0.18650203 |
| mean_time_to_correctly_identify_matches_20023 | 0.28651708 | 0.28590273 | 0.28510592 | 0.28413488 |
| number_of_puzzles_correct_21004 | 0.24675305 | 0.24698281 | 0.24833632 | 0.24500443 |
| number_of_puzzles_correctly_solved_6373 | 0.28743091 | 0.28647204 | 0.28515696 | 0.30237714 |
| number_of_symbol_digit_matches_made_correctly_23324 | 0.45860853 | 0.45790451 | 0.45735217 | 0.46345688 |

**Table 10: Survival analysis results of the nine DNEs and PRSs to predict the risk of mortality**. We first performed survival analyses for baseline data with the nine DNEs and the nine PRSs (PRS target sample). In addition, we evaluated the model's performance by incorporating the second scan from a small proportion (*N*=1348) of the entire UKBB sample (*N*=39,178) vs. using only the baseline data from the whole UKBB sample (*N*=39,178). Baseline analyses had smaller sample sizes because deriving PRS needs to split the UKBB samples into the base and target data.

| Type | DNE/PRS | hazard ratio | CI lower bound | CI upper bound | P-value | *N* |
|---|---|---|---|---|---|---|
| **a)**: Baseline (PRS target sample) | SCZ1 | 1.34262 | 1.1484 | 1.5697 | 0.00022 | 15,891 |
| | PRS-AD1 | 1.2328 | 1.06077 | 1.43274 | 0.00635 | 15,891 |
| | ASD1 | 1.23129 | 1.05442 | 1.43783 | 0.00855 | 15,891 |
| | SCZ1 | 1.21404 | 1.04527 | 1.41005 | 0.01109 | 15,891 |
| | LLD2 | 1.18426 | 1.01797 | 1.37771 | 0.02848 | 15,891 |
| | AD1 | 1.13838 | 0.98248 | 1.31901 | 0.08458 | 15,891 |
| | PRS-ASD2 | 1.12939 | 0.97213 | 1.3121 | 0.11173 | 15,891 |
| | SCZ2 | 1.11113 | 0.96016 | 1.28583 | 0.15728 | 15,891 |
| | PRS-SCZ2 | 1.09773 | 0.94547 | 1.2745 | 0.22097 | 15,891 |
| | AD2 | 1.08453 | 0.93551 | 1.25729 | 0.28194 | 15,891 |
| | PRS-LLD2 | 1.08457 | 0.93304 | 1.2607 | 0.29037 | 15,891 |
| | LLD1 | 0.93948 | 0.80971 | 1.09004 | 0.41039 | 15,891 |
| | PRS-LLD1 | 1.06235 | 0.91384 | 1.23499 | 0.43114 | 15,891 |
| | ASD2 | 1.05697 | 0.91201 | 1.22496 | 0.46162 | 15,891 |
| | ASD3 | 1.04856 | 0.90775 | 1.2112 | 0.51927 | 15,891 |
| | PRS-AD2 | 1.01102 | 0.87006 | 1.17482 | 0.88627 | 15,891 |
| | PRS-ASD3 | 1.00733 | 0.86682 | 1.17063 | 0.92404 | 15,891 |
| | PRS-ASD1 | 1.00583 | 0.86327 | 1.17192 | 0.94062 | 15,891 |
| **b)**: Longitudinal (full sample) | SCZ1 | 1.234652638 | 1.11683 | 1.36491 | 0.000038 | 39,178 |
| | AD1 | 1.157282427 | 1.05117 | 1.2741 | 0.00291 | 39,178 |
| | ASD1 | 1.153757457 | 1.04326 | 1.27596 | 0.00536 | 39,178 |
| | SCZ2 | 1.098631922 | 0.99877 | 1.20848 | 0.05304 | 39,178 |
| | LLD2 | 1.069535863 | 0.96898 | 1.18052 | 0.18204 | 39,178 |
| | LLD1 | 0.939699309 | 0.85192 | 1.03652 | 0.21383 | 39,178 |
| | ASD3 | 1.056200006 | 0.96027 | 1.16172 | 0.26041 | 39,178 |
| | ASD2 | 1.046340959 | 0.95022 | 1.15219 | 0.35686 | 39,178 |
| | AD2 | 1.012537735 | 0.91803 | 1.11678 | 0.80319 | 39,178 |
| **c)**: Baseline (full sample) | SCZ1 | 1.232871435 | 1.11525 | 1.3629 | 0.0000427 | 39,178 |
| | AD1 | 1.155808222 | 1.04977 | 1.27255 | 0.00318 | 39,178 |
| | ASD1 | 1.153214621 | 1.0427 | 1.27544 | 0.00555 | 39,178 |
| | SCZ2 | 1.096161394 | 0.99646 | 1.20583 | 0.05914 | 39,178 |
| | LLD2 | 1.071480848 | 0.97079 | 1.18262 | 0.17032 | 39,178 |
| | LLD1 | 0.939015519 | 0.85121 | 1.03588 | 0.20904 | 39,178 |
| | ASD3 | 1.055976564 | 0.96006 | 1.16147 | 0.26225 | 39,178 |
| | ASD2 | 1.049954778 | 0.95347 | 1.15621 | 0.32163 | 39,178 |
| | AD2 | 1.012534478 | 0.91792 | 1.1169 | 0.80346 | 39,178 |

**Table 11: The 9 BAG GWAS and 11 chronic diseases GWAS used in our Mendelian randomization analyses.** For the nine BAGs (excluding brain BAG due to overlapped samples), we used the GWAS summary statistics summary from our previous work[21], so the data did not need to be harmonized. For the 11 diseases, 4 GWAS were downloaded and manually harmonized (e.g., GRCh 37, flip the effect allele, etc.) from the PGC website (https://pgc.unc.edu/for-researchers/download-results/). Two studies (ASD and OCD) were not used in the Mendelian randomization analyses, in which the allele frequency information was unavailable. The other seven diseases of GWAS were also curated in our previous work[21].

| Trait | Dataset | Number of IVs | PubMed ID | Ancestry |
|---|---|---|---|---|
| AD | PGC | 23 | 34493870 | European |
| ADHD | PGC | 13 | 36702997 | European |
| BIP | PGC | 29 | 34002096 | European |
| SCZ | PGC | 80 | 35396580 | European |
| Glaucoma | finn-b-H7_GLAUCOMA | 9 | NA | European |
| RA | ebi-a-GCST005569 | 11 | 23143596 | European |
| PBC | ebi-a-GCST003129 | 16 | 26394269 | European |
| CD | ieu-a-12 | 77 | 26192919 | European |
| IBD | ieu-a-292 | 81 | 23128233 | European |
| Breast cancer | ieu-a-1126 | 86 | 29059683 | European |
| Type 2 diabetes | ieu-a-26 | 10 | 22885922 | European |
| Cardiovascular BAG | UKBB (non-overlap) | 34 | 37398441 | European |
| Eye BAG | UKBB (non-overlap) | 16 | 37398441 | European |
| Hepatic BAG | UKBB (non-overlap) | 37 | 37398441 | European |
| Immune BAG | UKBB (non-overlap) | 47 | 37398441 | European |
| Metabolic BAG | UKBB (non-overlap) | 62 | 37398441 | European |
| Musculoskeletal BAG | UKBB (non-overlap) | 24 | 37398441 | European |
| Pulmonary BAG | UKBB (non-overlap) | 49 | 37398441 | European |
| Renal BAG | UKBB (non-overlap) | 42 | 37398441 | European |
